# Mind’s eye: widespread saccade-related evoked potentials support visual memory encoding in humans

**DOI:** 10.1101/2025.11.11.25339896

**Authors:** Gansheng Tan, Phillip Demarest, Yilin Li, Hohyun Cho, Haeorum Park, James R. Swift, Cory S. Inman, Joseph R. Manns, Stephan B. Hamann, Xiaoxuan Liu, Krista L. Wahlstrom, Ziwei Li, Martina K. Hollearn, Justin M. Campbell, Patrick E. Cettina, Siddharth S. Sivakumar, Eric C. Leuthardt, Jon T. Willie, Peter Brunner

**Author notes:** communicating authors: Jon T. Willie and Peter Brunner.

## Abstract

Saccadic eye movements modulate medial temporal lobe (MTL) activity in primates, a region critical for memory, yet whether and how this saccadic modulation supports memory encoding in humans remains unknown. By combining eye tracking with intracranial local field potentials in humans performing memory tasks, we identified widespread cortical evoked potentials time-locked to saccades (saccade-related evoked potentials, SREPs) that could not be explained by oculomuscular artifacts or retinal input. SREPs emerge earliest and with the greatest amplitude in the lateral temporal cortex. Machine learning analysis revealed that pre-saccadic oscillation and SREPs predicted recognition memory but not saccade direction, suggesting that SREPs are distinct from corollary discharge. Frontoparietal SREPs, rather than those in MTL, best predict memory outcomes, and successful encoding is associated with earlier, more pronounced SREPs. This study extends the active sensing framework to human visual memory: saccades are accompanied by a structured cortical event that influences memory encoding.

**Teaser:** Every eye movement is accompanied by a ripple of coordinated neural activity that shapes whether a visual experience will be remembered.

**Graphical abstract:** 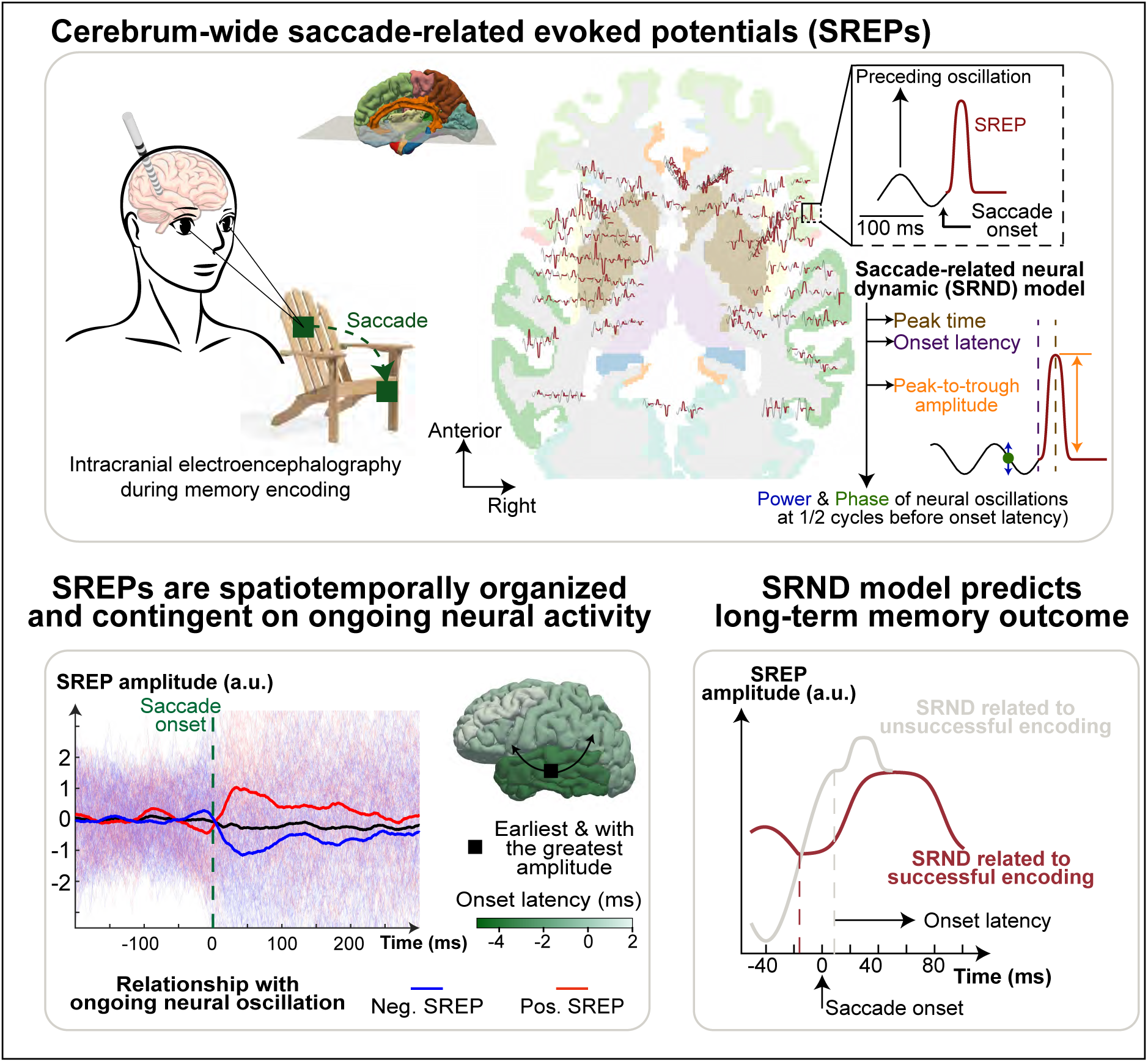

## Introduction

The mental faculty of generating, remembering, and recalling visual representations is the “mind’s eye”. During active vision, visual representations are built from sensory input gated by quasi-rhythmic eye movements that alternate between saccades and fixations(*1*). Saccades impose a predictable temporal structure on visual sampling. This temporal structure may help coordinate neural dynamics with saccades, supporting the mind’s eye during memory encoding(*2*, *3*). However, most neurophysiological accounts of visual memory encoding have not yet explicitly incorporated saccade-related neural dynamics.

Saccades in primates were accompanied by changes in electrical potentials in the medial temporal cortex(*3–6*). These changes in local field potentials (LFPs) have been shown to synchronize single-unit activity and reset the phase of neural oscillation, suggesting saccadic modulation of neural circuits(*3*, *7*, *8*). Here, we refer to such consistent LFP waveforms time-locked to a saccade as saccade-related evoked potentials (SREPs). While one interpretation is that SREPs are responses to a changing visual image on the retina, findings of SREPs in darkness and during rapid eye movement sleep challenge this hypothesis and suggest additional functions beyond sensory response(*9*, *10*). As saccades gate the timing of information entering the visual system, SREPs are well-positioned to coordinate when and how visual information is processed for effective memory encoding(*11*). Saccadic modulation of neural activity has almost exclusively been studied within single anatomical-functional regions. Most nonhuman primate studies and a small number of human intracranial studies have examined saccadic modulation in the medial temporal lobe (MTL), particularly the hippocampus(*3*, *5*, *6*, *8*, *9*, *12–14*). This MTL-centric view leaves unresolved a foundational question: do SREPs exist broadly across the cortical mantle, or are they confined within the MTL? Resolving this question is essential for determining the spatial specificity of SREP–memory relationships and for designing rational neuromodulation strategies. If SREPs are local MTL responses, then stimulation of the MTL at saccade onset may enhance the encoding mechanism. In contrast, if MTL SREPs are part of a broader cortical network, then stimulation of the MTL alone may be insufficient to support encoding. Data on the spatiotemporal organization of SREP remain scarce, largely due to the MTL-centric view of visual memory and the methodological constraints listed below.

Investigating the existence of SREPs and their spatiotemporal organization requires synchronized eye tracking and neural recordings. Because saccades occur over only tens of milliseconds, neural recordings must achieve sufficient temporal precision to capture the related neural dynamics. Neuroimaging methods, such as functional magnetic resonance imaging (fMRI), offer good spatial resolution but are limited in temporal resolution, whereas noninvasive methods, such as scalp electroencephalography, provide millisecond temporal resolution but lack spatial resolution to identify the underlying neural source(*15*). Source localization remains a challenging inverse problem that depends heavily on assumptions and parameter choices(*15*). Compounding these challenges, electrical signals generated by extraocular muscles can confound the analysis of SREP, as reported in human electrophysiology(*16*).

A more fundamental question concerns the function of SREPs. One hypothesis posits SREP as a form of *corollary discharge*, a copy of the motor signal that prepares the brain for the sensory consequences of an impending movement(*17*). In vision, the corollary discharge mechanism maintains visual continuity despite the disruption of visual input caused by saccades(*18*). Two pathways underlying the corollary discharge mechanism have been previously identified in nonhuman primates: the projections from the superior colliculus (SC) to the frontal eye field via the medial dorsal thalamic nucleus and to the middle temporal visual area through the inferior pulvinar. A recent study observed saccadic modulation of neuronal activity exhibiting some properties of corollary discharge in the human medial temporal lobe, a region involved in high-level visual processing and memory formation(*8*). This finding also comports with the active sensing framework, which posits that motor sampling, such as sniffing in rodents and saccades in primates, is not random but part of a coordinated strategy that optimizes sensory information processing(*19–22*) (**Figure 1A**). Although the literature suggests that saccadic modulation may play a role in memory encoding, direct evidence in humans has yet to be established.

**Figure 1.**
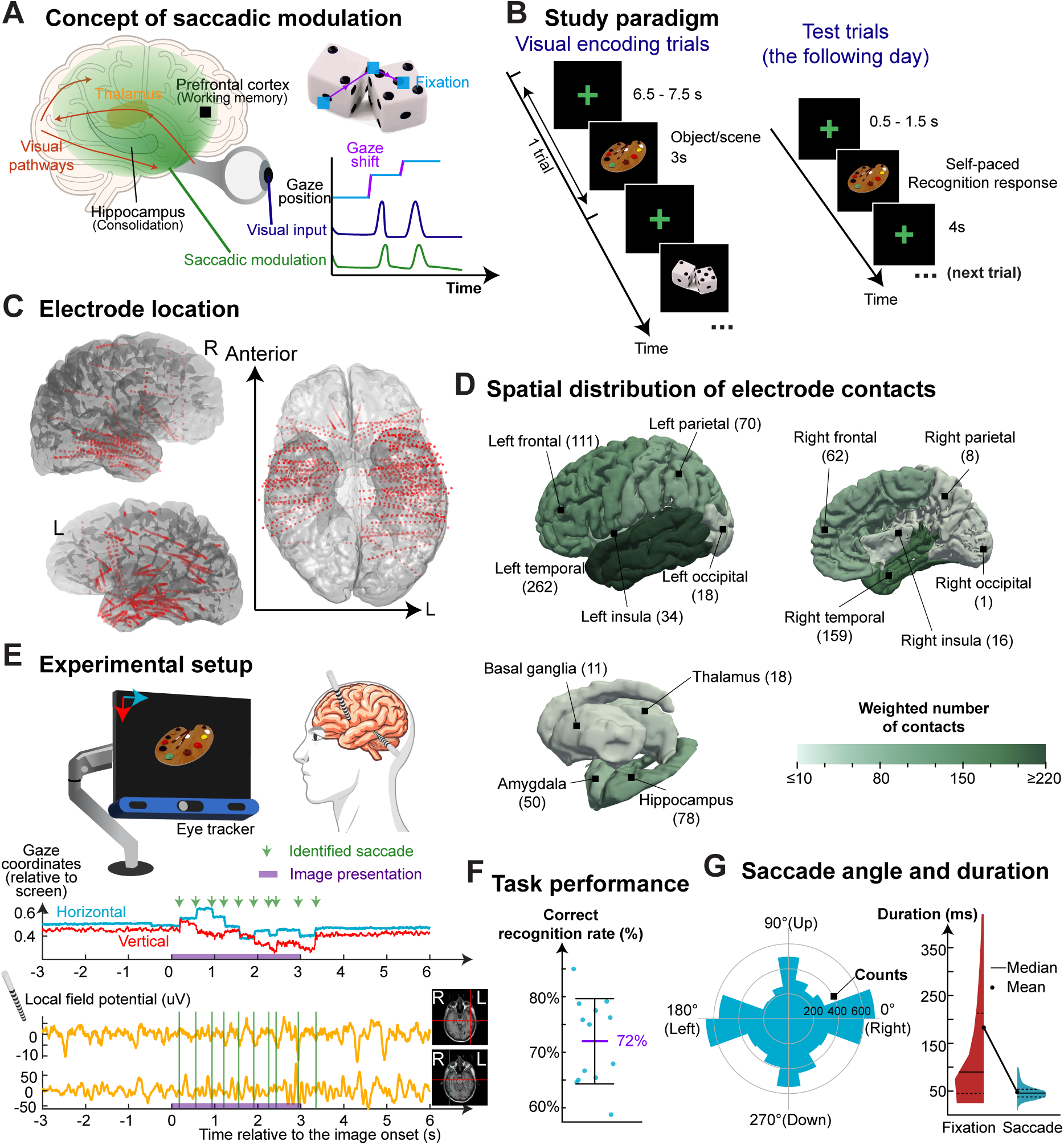
Experimental design for studying saccadic modulation in humans. **(A)** Concept of saccadic modulation of neural activity. Neural processing of visual information peaks shortly after fixation onset, when visual acuity is at its maximum. Retinal inputs are transmitted via optic nerves and tracts to the visual cortex via the lateral geniculate nucleus and optic radiations. **(B)** Study paradigm. Subjects were asked to view a series of objects and scenes, each presented for 3 seconds. Recognition memory was assessed the following day, where subjects used a keyboard to indicate whether they recognized the previously presented images among a series of old and new images. Examples of actual visual stimuli are shown in **Fig. S1G**. **(C)** Electrode location. We localized implanted stereoelectroencephalography (SEEG) electrodes in subject-specific anatomical space using post-implantation computed tomography (CT) imaging, co-registered to pre-implantation MRI. For visualization, we transformed electrode coordinates into a common anatomical reference space obtained from a reference subject (Subject ID 1). **(D)** Regional distribution of electrode contacts. The brain model is divided into lobes (i.e., frontal, temporal, parietal, and occipital cortices), the insula and cingulate cortices, and four subcortical regions of interest (i.e., amygdala, hippocampus, thalamus, and basal ganglia). Color represents the electrode density—weighted number of electrode contacts per region. When individual electrode-contact volume (∼2 mm³) overlapped voxels assigned to multiple brain regions, their contributions were proportionally allocated among these regions. White matter and cingulate regions are not shown. The left and right white matter contained 446 and 193 sampling electrode contacts, respectively. The left and right cingulate contained 19 and 9 contacts, respectively. The sampling density, defined as the ratio of the number of electrode contacts per unit anatomical volume, was greatest in the amygdala (13 contacts/cm^3^), followed by the hippocampus (7 contacts/cm^3^), and the left temporal cortex (4 contacts/cm^3^, **Fig. S2A**). (**E)** Experimental setup. BCI2000 controlled stimulus presentation, recorded local field potentials from SEEG electrodes, and tracked eye movements. We identified saccade onsets and offsets based on gaze coordinates using a clustering algorithm (see **MATERIALS AND METHODS**). **(F)** Task performance. Each data point represents the recognition accuracy for one subject performing the visual encoding task described in panel **B**. All eight subjects confirmed that they performed the task as instructed during a post-experiment debriefing. (**G**) Distribution of saccade angle and direction.

Besides SREP, another principal method for quantifying saccadic modulation in electrophysiological signals is spectral dynamics(*3*, *6*). Spectral dynamics analysis typically requires longer time series that span multiple cycles of the assumed oscillation to accurately estimate its periodic properties(*23*). Recent work has emphasized the importance of separating strictly defined rhythmic components from aperiodic activity when interpreting power spectra, and the methodology and interpretation of these components are under active investigation(*24*). Therefore, we focused on the form of saccadic modulation, SREP. Intracranial SREPs capture the rapid extracellular voltage deflections associated with synchronous neuronal activity following saccades(*7*, *8*, *25*).

This study characterizes the cortical dynamics of saccadic modulation and tests our hypothesis that saccadic modulation predicts recognition memory, challenging the MTL-centric view of visual memory encoding. The present study addresses technical challenges of studying saccadic modulation in humans by combining intracranial recordings in neurosurgical patients, synchronized gaze tracking, and rigorous control analyses to confirm the neural origin of SREPs. To identify the potential cortical pathway underlying saccadic modulation, we quantified SREP waveform consistency and characterized SREP onset latency and magnitude. We found that earlier and higher-amplitude SREPs were spatially clustered in the temporal lobe. Next, we tested our central hypothesis using explainable machine learning analysis to reveal the mapping between SREP characteristics and the memory outcome(*26*). In general, earlier SREPs with greater magnitude and later peak times predicted successful visual encoding. Finally, to assess whether these SREPs reflect a narrowly defined corollary discharge signal, we evaluated whether they encoded saccade direction(*17*). We confirmed SREPs encode either the current or subsequent saccade direction very rarely, if at all. Altogether, SREPs support visual encoding in the human brain, as a form of saccadic modulation, separate from classical corollary discharge.

## Results

### Visual memory encoding performance and eye movement characteristics

Eight human subjects participated in this study (see **Table S2** for patient characteristics). Across the eight subjects, 129 implanted electrode arrays contained a total of 1,629 electrode contacts, with 204 ± 30 (mean ± SD; SD, standard deviation) contacts across 16 ± 2 arrays per subject (**Figure 1C**). We identified and excluded electrode contacts exhibiting epileptiform activity or artifactual signals (e.g., due to poor contact), resulting in an average of 201 ± 29 valid contacts per subject (**Table S3**). For 6 subjects, two Behnke Fried Depth electrodes (AD-TECH), each with 8-contact micro-wires, were implanted. The resulting electrode coverage sampled both cortical and subcortical regions, although coverage was absent in the right occipital cortex, as reflected in the regional distribution of electrode contacts and sampling density maps (**Fig. 1D and Fig. S2A**).

Subjects participated in a series of visual memory encoding tasks, each comprising an encoding phase followed by a test phase the next day (**Fig. S1G** for exemplar stimuli). During each encoding trial, an image of an object or scene was presented on a screen. (**Figure 1B**). Subjects freely scanned the presented images (**Figure 1E**). Subjects completed one or more visual memory encoding tasks with distinct image sets on non-overlapping days, when feasible. The number of completed tasks depended on the duration of each patient’s stay in the epilepsy monitoring unit (**Table S1**).

We verified that subjects performed the task as instructed. All subjects exhibited memory performance above chance (i.e., 50%, see **MATERIALS AND METHODS:** Assessing memory outcome). The overall correct recognition rate, which incorporates both correct identification of previously presented images and correct rejection of novel images, was 72% ± 8% (mean ± standard deviation, SD) (**Figure 1F, Fig. S1F, Table S3**). Subjects exhibited saccadic kinematics consistent with human active visual sampling of the environment(*27*). The mean fixation duration was 183 ms (SD = 266 ms); and the mean saccade duration was 48 ms (SD = 15 ms, **Figure 1G**). The saccade directions were dispersed, with relatively higher probabilities of horizontal and vertical eye movements (35.5% and 26.5%, respectively, **Figure 1G**). These results indicate that subjects were attentive and visually scanned the presented images.

### SREPs are consistent, widespread, and exhibit two polarities

We first confirmed whether SREPs documented in nonhuman primate studies translate to humans(*3–5*) by quantifying the consistency of intracranial LFPs across saccades. In the presence of SREPs, we expected that the waveform consistency within the peri-saccadic window would be greater than that within the pre-saccadic baseline (*28*). The correlations between pairs of peri-saccadic LFPs exhibited a clear bimodal distribution, compared to the approximately normally distributed waveform correlations during baseline (**Figure 2A, Fig. S4A**). For 98% of electrode contacts, peri-saccadic waveform correlations showed significant deviation from this baseline distribution, suggesting widespread saccadic modulation of LFP across the brain (**Fig. S4B**, median p-value <1 × 10^−20^, significance threshold after correcting for multiple comparisons = 1.9 × 10^−5^, Kolmogorov-Smirnov tests). The magnitude of this deviation, quantified by the area under the absolute difference curve between peri-saccadic and baseline empirical cumulative distributions of waveform correlation, exceeded the effect size expected at the corrected significance threshold across most contacts (expected significance threshold effect size = 0.017; observed 2.5–97.5 percentile range = 0.011–0.228; **Fig. S4A, B**). To assess the temporal extent of waveform consistency, we systematically varied the peri-saccadic and baseline window sizes. This analysis showed that waveform consistency was strongest within a brief peri-saccadic interval (<100 ms; see Fig. S6 and Supplementary Materials).

**Figure 2.**
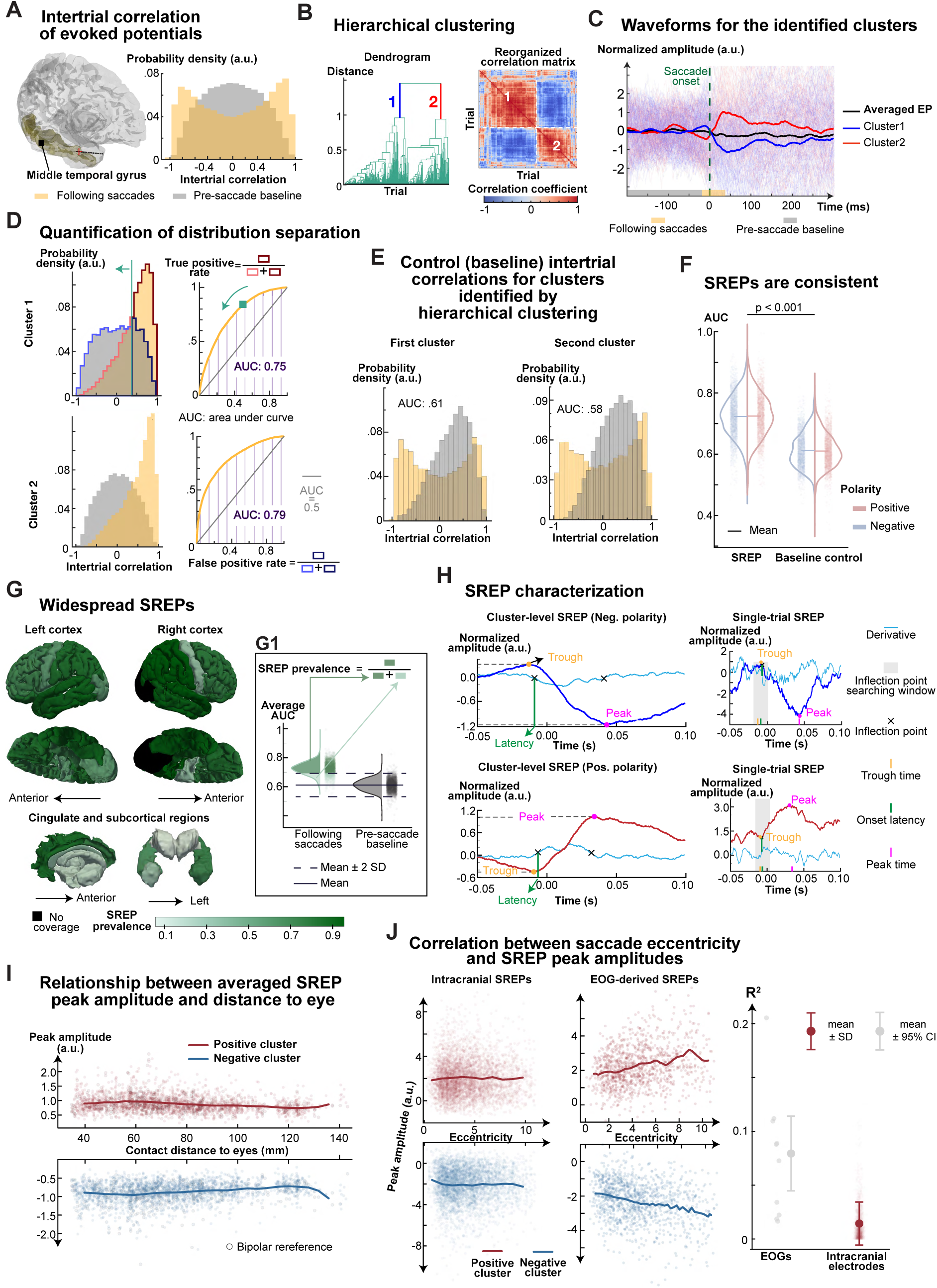
Evoked potentials following saccades are consistent and widespread across the cerebrum. **(A)** Intertrial correlation of local field potentials (LFPs). To determine the presence of saccade-related evoked potentials (SREPs), we calculated the intertrial correlation of LFPs during the peri-saccadic and the baseline window (i.e., -10 ms to 40 ms, and -200 ms to -10 ms, relative to saccade onsets, respectively). Here, each trial is defined as an LFP segment centered on a saccade event. The exemplar result for a representative electrode contact in the right middle temporal gyrus shows an intertrial correlation distribution during the peri-saccadic period that is skewed toward the tails. In contrast, the corresponding baseline intertrial correlation appears normally distributed. **(B)** Hierarchical clustering. The bimodal nature of the peri-saccadic correlation distribution suggests the presence of two EP clusters, with trials within a cluster positively correlated and those between clusters negatively correlated. The dendrogram and the reordered intertrial correlation matrix confirm the presence of two distinct SREP waveforms (shown in panel **C**). **(C)** Evoked potentials averaged across trials in clusters 1 and 2. For normalization, we z-scored LFPs before hierarchical clustering. Each line represents the z-scored LFP of one trial. **(D)** Quantification of the separation between peri-saccadic and baseline correlation distributions. We used a sliding threshold to compute the receiver operator characteristic (ROC) curve. For a set of thresholds from -1 to 1, we calculated the true positive rate (fraction of peri-saccadic probability density above a threshold relative to the combined density) and false positive rate (fraction of peri-saccadic probability density below a threshold relative to the combined density) for each threshold. We used the area under the ROC curve (AUC) to measure separability. **(E)** Hierarchical clustering applied to baseline intertrial correlation. This procedure accounts for potential AUC inflation due to hierarchical clustering. **(F)** Peri-saccadic and baseline AUCs for intertrial correlations. AUC following saccades was significantly higher than baseline AUCs (Cohen’s d = 1.857, two-tailed paired t-test, N = 2604, t statistic = 81.13, p < 0.001), indicating robust SREPs. Each dot represents an electrode contact in a single subject during a memory encoding task. Colors denote the SREP polarity based on the sign of the first peak (detailed in Panel **H**). **(G)** SREP prevalence across the cerebrum. Color represents SREP prevalence, which is defined as the proportion of electrode contacts with AUC in the peri-saccadic condition above the mean + 2 standard deviation (SD) of baseline AUC. The lighter color for the precentral gyrus and paracentral lobule does not indicate an absence of SREPs in these regions (58% and 37% for left and right hemispheres), and as shown in Figure 6H, SREPs from these regions contributed strongly to predicting memory outcomes. Electrode contacts are co-registered to a common reference brain model. The brain was segmented into Hippocampus (Left, Right), Amygdala (Left, Right), Basal Ganglia, Thalamus, Orbitofrontal (Left, Right), Inferior Frontal (Left, Right), Middle Frontal (Left, Right), Superior Frontal (Left, Right), Anterior Cingulate, Posterior Cingulate, Motor (i.e., precentral gyrus and paracentral lobule, Left, Right), Anterior Temporal (i.e., temporal pole, Left, Right), Middle Temporal (Left, Right), Superior Temporal (Left, Right), Inferior Temporal (Left, Right), Medial Temporal (Left, Right), Fusiform (Left, Right), Parietal (Left, Right), Occipital (Left, Right), and Insula (Left, Right). **Fig. S3** shows response ratios across the brain at various AUC thresholds (e.g., mean + 3 SD of baseline AUC) used to define SREP prevalence. **Fig. S4** shows the spatial distribution of intertrial consistency. **Fig. S5** shows the electrode contact-level intertrial consistency. **(H)** Characterization of averaged and single-trial SREPs. We characterized SREPs using latency, peak amplitude, trough amplitude, peak time, and trough time. After hierarchical clustering, we used MATLAB’s *findchangepts* function to identify two inflection points in the derivative of SREPs averaged across trials within a cluster. We defined latency as the first point and peak time as the time point within a 10-ms window around the second inflection point at which the z-scored SREP amplitude was maximal for positive clusters (or minimal for negative clusters). Similarly, we defined trough time as the time point within a 10-ms window around latency at which the z-scored SREP amplitude is minimal for positive clusters (or maximal for negative clusters). We identified the onset latency for single-trial SREP as the inflection point near the onset latency of the corresponding averaged SREP. We identified the peak and trough amplitudes for single-trial SREP by finding the maximum and minimum SREP amplitudes around the corresponding cluster-level peak and trough times. **(I)** Relationship between the averaged SREPs’ peak amplitude and eyeball-to-electrode distance. The x-axis represents the distance from the recording electrode contact to the nearest eye. The lines represent the average peak amplitude calculated over sliding windows of 10 mm, incremented by 5 mm. The distance explained 3.8% and 3.5% of the variance in the peak amplitude of averaged SREPs for positive and negative clusters, respectively, across all electrode contacts. **(J)** Relationship between saccade eccentricity and peak amplitudes of single-trial SREP derived separately from local field potentials (LFPs) and electrooculogram (EOG) signals. For SREP derived from LFPs, for both polarities, saccade eccentricity explained 0.0% of the variance in peak amplitude. Due to the large sample size, we show 2% of the data in the plot. We extracted electrooculogram (EOG) signals from frontopolar scalp EEG electrodes (Fp1 and Fp2) using bipolar re-referencing. Eccentricity explains 7.0% and 6.8% variance in oculomuscular signal peak amplitude for positive and negative clusters. The right plot compares correlation coefficients between saccade eccentricity and single-trial SREP amplitudes for intracranial electrodes and EOG. For each electrode contact, we calculated the correlation between eccentricity and single-trial SREP amplitudes for this contact. The mean correlation coefficient was 0.014 for intracranial electrodes and 0.079 for oculomuscular signals. Welch’s t-test for comparison between unbalanced sample sizes showed that correlation coefficients for EOGs were significantly higher than intracranial local field potential signals (t = 3.154, p = 0.012, Cohen’s d = 3.193, *N_EOG_* = 10, *N_EOG_* = 2367). For EOG correlation coefficients, the horizontal bars represent the bootstrapped 95% confidence interval (CI).

The bimodal peri-saccadic correlation distribution implied two internally consistent but polarity-opposed SREP waveforms. Consistent with this interpretation, agglomerative hierarchical clustering applied to peri-saccadic waveform correlations revealed two clusters of SREPs (**Figure 2B, C**). Therefore, we compared peri-saccadic and baseline waveform consistency for each cluster using the area under the receiver operating characteristic (ROC) curve(*28*) (see **MATERIALS AND METHODS:** Identification of SREP, **Figure 2D**). To control for hierarchical clustering-induced consistency inflation, we applied hierarchical clustering to baseline waveform correlations (**Figure 2E**). The SREP waveform consistency was significantly higher than the baseline-derived control waveform consistency (Cohen’s d = 2.651, two-tailed paired t-test, N = 2604, t statistic = 81.13, p < 0.001, **Figure 2F**). Our data show that SREPs in humans are consistent across saccades and exhibit two waveform polarities.

We next asked whether SREPs were localized to specific anatomical circuits or broadly distributed throughout the sampled cortex. The SREP prevalence (see **MATERIALS AND METHODS:** SREPs prevalence) across all cortical regions, except the right occipital lobe (not sampled) and right motor cortex (i.e., precentral gyrus and paracentral lobule, 37%), exceeded 50% (**Figure 2G**). The fusiform gyrus (left: 92.0%, right: 97.0%), lateral temporal cortex (left: 91.0%, right: 96.7%), and parietal lobes (left: 86.0%, right: 100.0%) were the three regions with the highest SREP prevalence. Control analysis using a threshold-independent estimate of SREP prevalence is included in the **Supplementary Materials**. Altogether, SREPs in humans are consistent and widespread.

### SREPs recorded in iEEG reflect saccadic modulation

Intracranial recordings have a high signal-to-noise ratio compared to noninvasive modalities and are therefore widely regarded as being free of electrooculogram (EOG) artifacts(*29*). Nevertheless, one study showed that signals recorded from subdural electrocorticography grids may be contaminated by EOG artifacts(*16*). In this study, Kovach et al. correlated the peak-to-trough amplitude of EPs at saccade times with saccade eccentricity and electrode-to-eyeball distances(*16*). Given this precedent, we tested two predictions that would hold if the observed SREPs reflect EOG contamination: (1) Cluster-level averaged SREP peak amplitude would decrease with increasing distance between the electrode contacts and the nearest eye globe center, which approximates the distance between electrode contacts and the myogenic dipole source generated by extraocular muscle activation; and (2) Single-trial SREP peak amplitude would increase with greater saccade eccentricity, reflecting stronger extraocular muscle activation. Furthermore, if SREPs reflect EOG activities, then positive and negative polarity SREPs may result from opposing myogenic field directions associated with distinct saccade directions. We used these predictions as falsifiable tests of the EOG-contamination account (see below and **Results**: Phase of preceding oscillatory activity associated with SREP polarity).

We tested the first prediction by quantifying peak amplitude for cluster-level SREPs and single-trial SREPs and regressing it against the distance from each contact to the eyeballs. (**Figure 2H**). The distance from the eyeballs accounted for only a minor fraction of variance in peak amplitude of averaged SREPs (**Figure 2I**, explained variance = 3.8%, *N_positive_* = 2573 for positive polarity; explained variance = 3.5%, *N_negative_* = 2575 for negative polarity, linear regression). To ensure that our findings were not dependent on a specific choice of SREP amplitude metric, we adopted the measure used in Kovach et al.(*16*), peak-to-trough amplitude. Distance from the eyeballs explained only 0.9% and 0.7% of the variance in peak-to-trough amplitudes of cluster-level SREPs for positive and negative clusters, respectively, across all electrode contacts (**Fig. S7C**, *N_positive_* = 2571; *N_negative_* = 2570, linear regression). These results do not support the first prediction.

To test the second prediction, we assessed whether single-trial SREP peak amplitude scaled with saccade eccentricity. Linear regression revealed that saccade eccentricity explained 0.0% of the variance in peak amplitude for both polarities (**Figure 2J**, |estimated coefficient| < 0.001, p < 0.001, *N* = 250531 for positive polarity; |estimated coefficient| < 0.001, p = 0.156, *N* = 250614 for negative polarity; linear regression with t-test). For both polarities, saccade eccentricity explained 0.0% variance in single-trial SREP peak-to-trough amplitude (**Fig. S7D,** |estimated slope| < 0.001, p = 0.020, *N* = 250531 for positive polarity; |estimated coefficient| < 0.001, p = 0.079, *N* = 250614 for negative polarity; linear regression with t-test). The results show no relationship between SREP peak amplitude and saccade eccentricity.

As a positive control, we confirmed that EP peak amplitude in electrooculogram (EOG) correlated with saccade eccentricity (**Figure 2J**). Compared to SREPs recorded with intracranial electrodes, single-trial EP peak amplitude from EOG signals showed a strong dependence on saccade eccentricity, with larger estimated slopes and greater explained variance (**Figure 2J**, explained variance = 7.0%, estimated slope = 0.133, p < 0.001, *N* = 1100 for positive polarity; explained variance = 6.7%, estimated slope = -0.130, p < 0.001, *N* = 1126 for negative polarity; linear regression with t-test). This dependence holds when we quantified single-trial EP magnitude from EOG signals using peak-to-trough amplitudes (**Fig. S7E**, explained variance = 2.7%, estimated slope = 0.108, p < 0.001, *N* = 1100 for positive polarity; explained variance = 8.2%, estimated slope = -0.171, p < 0.001, *N* = 1126 for negative polarity; linear regression with t-test). The variance explained by saccade eccentricity was significantly greater in the ocular-muscular signals captured in fronto-polar scalp EEG electrodes compared to intracranial recordings (**Figure 2J**, t = 3.154, p = 0.012, Cohen’s d = 3.193, *N_E0G_* = 10, *N_intracranial_* = 2367, Welch’s t-test, rank-biserial correlation = 0.833). These results convincingly demonstrated that the observed SREPs cannot be explained by EOG signals.

We next examined whether changes in visual input drove the SREPs. If this were the case, we would expect reduced or absent SREP consistency following saccades that occurred in the absence of meaningful visual changes. To test this, we analyzed SREP consistency during intertrial intervals, when participants made saccades on a uniform black background containing a cross at the center. Despite the absence of changes in visual inputs, SREP consistency during cross presentation was significantly higher than baseline (**Fig. S7H**, paired t-test, p<0.001, Cohen’s d = 2.631, N = 2604). Across electrode contacts, SREP consistency during image presentation was positively correlated with that during fixation-cross presentation (**Fig. S7I**, Pearson correlation coefficient = 0.752, p < 0.001, based on t-distribution). This correlation indicated regional specificity in how saccades modulate LFPs independently of concurrent visual input. The spatial pattern of SREP prevalence during fixation-cross presentation was similar to that during image presentation (**Fig. S7G**, **Figure 2G**). These results demonstrate that SREPs are not attributable to changes in visual input nor oculomuscular signals.

### SREP polarity is associated with the preceding oscillatory phase

We next sought to understand the physiological basis of our observation that SREPs exhibited two opposing polarities. We considered two parsimonious mechanisms to explain SREP bipolarity: (1) indirect volume-conducted oculomuscular activity, which we have already shown to be unlikely; and (2) direct neural activity encoding saccade planning or execution. Both mechanisms predict polarity-specific directional clustering, i.e., that saccades associated with SREPs of the same polarity exhibit greater directional clustering compared to all saccades, and that saccade directions differ between SREPs of opposing polarities.

To test these predictions, we quantified directional clustering of saccades using Rayleigh statistics. If there were polarity-specific directional clustering, Rayleigh statistics of saccade direction associated with each polarity would be higher than those computed from saccades pooled across both polarities. Our analysis showed no evidence of polarity-specific directional clustering. For both polarities, differences in Rayleigh statistics between monopolar and combined conditions were centered at zero (**Fig. S8C,** t = -0.953, p = 0.341, Cohen’s d = -0.022, *N* = 2068 for positive polarity; t = -1.028, p = 0.304, Cohen’s d = -0.023, *N* = 2058 for negative polarity; two-tailed t-test). Across electrode contacts, there was no significant difference in saccade direction between SREPs of opposing polarities (95% CI for W statistics = [1.926, 2.107], the corresponding 95% CI for p-value is [0.486, 0.512], Mardia-Watson-Wheeler test). In contrast to SREPs, EPs at saccade times derived from EOG signals, capturing volume-conducted oculomuscular activities, exhibited strong polarity-specific directional clustering (**Fig. S8A**). For EOG signals, polarity-specific Rayleigh statistics were significantly higher than those computed from the same number of saccades pooled across polarities (**Fig. S8B,** t = 5.950, p < 0.001, Cohen’s d = 1.881, *N* = 15 for positive polarity; t = 4.553, p = 0.001, Cohen’s d = 1.440, *N* = 18 for negative polarity; paired t-test). The circular distribution of saccade direction differed significantly between positive and negative EPs from EOGs at saccade times (p < 0.001 for all subjects, 95% CI for W statistics = [33.223, 46.050], Mardia-Watson-Wheeler test). These findings show that neither oculomotor nor neural activity coding saccade directions explains the observation of opposing SREP polarities.

Could the phase of preceding oscillatory activity influence SREP polarity? This hypothesis stems from the phase-locking relationship between saccade timing and alpha oscillations reported in a previous study(*2*). To test this hypothesis, we first assessed the presence of neural oscillations and their dominant frequencies based on the one-over-frequency (1/*f*)-normalized power spectral density, which is necessary for estimating oscillatory phase with the Hilbert transform (**Figure 3A, B**). Across contacts, dominant oscillatory frequencies varied by location and were centered in the alpha range (∼11 Hz, **Fig. S8D**), with a mean oscillation power of 5.9 *μV*^2^/*Hz* (**Fig. S8E)**. We then estimated the phase of preceding neural oscillations at their dominant frequency. Quantification of difference in oscillation phase for positive and negative SREPs at five time points: 0.5, 1, 1.5, 2, and 3 oscillatory cycles (OCs) before the onset latency of each SREP revealed that phase separation was significantly higher than the null distributions generated by permuting SREP polarity labels (**Figure 3C**, p < 0.001 for all time points, paired Wilcoxon signed-rank test, N = 1970 pairs). The degree of separation, quantified by the rank-biserial correlation, continuously increased as time approached SREP onset latency, from 0.204 (3 OCs before SREP onset latency) to 0.581, 0.790, 0.901, and 1.000 (0.5 OCs before SREP onset latency). These results show that for each electrode contact, SREP polarity is associated with the phase of pre-SREP neural oscillations.

**Figure 3.**
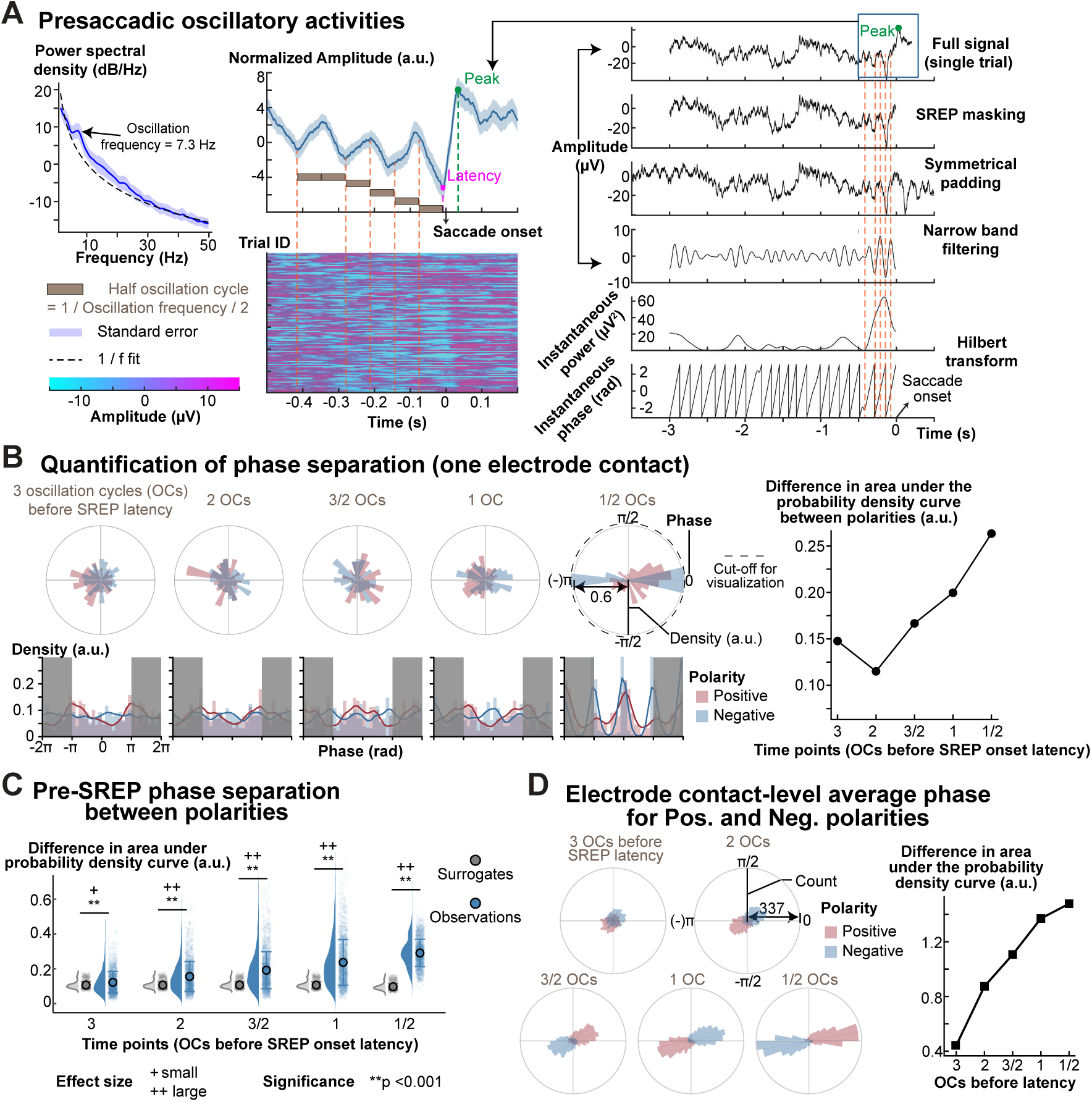
Pre-saccadic oscillatory activity determines SREP polarities following saccades. **(A)** Identification of ongoing oscillation. To account for the aperiodic activity, we fitted a one-over-frequency (1/*f*) curve. We defined the oscillation frequency as the frequency at which the residual of the power spectral density from the 1/*f* fit was largest. The middle panel shows the averaged SREP and single-trial SREPs. The heatmap shows the voltage signals from single trials. We estimated the instantaneous phase and power at 5 time points: 0.5, 1, 1.5, 2, and 3 oscillatory cycles (OCs) before the SREP onset latency. Specifically, we applied the Hilbert transform to padded, narrowband-passed signal segments preceding the SREP onset latency to avoid potential SREP influence on phase estimation. We only retained instantaneous power and phase corresponding to the masked, unpadded signal. **(B)** Pre-saccadic oscillation phase for the electrode contact shown in panel A and quantification of phase separation between SREPs of opposing polarities. The circular histograms show the phase distribution for an electrode contact, colored by SREP polarity. To account for the circular nature of phase (i.e., -π is equal to π), we unwrapped phase values and extended the phase range from [-*π*, *π*] to [-2*π*, 2*π*]. We then estimated the probability density function and calculated the absolute difference in the area under the probability density curve to quantify the phase separation between polarities. **(C)** Phase separation between polarities across electrode contacts. Each data point represents the difference in the area under the probability density curve for one electrode contact. To assess whether the phases preceding SREP were different for positive and negative SREPs, we generated the surrogate (null) distribution by randomly permuting the polarity labels. The grey dots indicate the mean separation metric from the 50 permutations for each electrode contact. We used a paired Wilcoxon signed-rank test (two-sided, N = 1970 pairs per condition) to compare observed and null phase-separation metrics across electrode contacts for each time point. All comparisons indicated statistically significant differences (p < 0.001). Effect sizes (rank biserial correlation, a non-parametric analog to Cohen’s d) increased progressively across conditions: from 0.204 in the earliest time point and 1.000 in the latest, suggesting strengthening of phase separation between polarities as time approached saccades. The rank-biserial correlations were 0.204, 0.581, 0.790, 0.901, and 1.000 for the 5 time points. The data shown are from electrode contacts that exhibit both oscillatory activities and consistent SREP following saccades. We defined contacts exhibiting oscillatory activities as those in which the largest deviation between the power spectral density and the fitted 1/*f* aperiodic curve occurred at a frequency below 30 Hz. This criterion excludes broadband activity (the elbow in the PSD) or high-frequency artifacts, such as 60 Hz line noise, resulting in 1970 electrode contacts (**Fig. S8D**). **(D)** Average phase across trials for positive and negative SREPs. The circular histogram shows the distribution of phase for the two polarities averaged across trials for each electrode contact. The right plot shows increased separation in the average phase between polarities as the time approaches SREP latency.

Previous analyses demonstrate a relationship between the pre-SREP oscillatory phase and SREP polarity. We asked whether this relationship was consistent across the sampled cerebrum. If the relationship reflects a spatially consistent organizing principle, then the mean phase preceding positive and negative SREPs should cluster by polarity across electrode contacts. To test this, we computed the circular mean phase separately for positive and negative SREPs at each contact by averaging across saccades. Across contacts, mean phases formed distinct polarity-specific clusters (**Figure 3D**, all Bonferroni-corrected p < 0.001, number of comparisons = 10, *N_positive_* = 1975, *N_positive_* = 1976, Rayleigh test). Phase concentration increased progressively as time approached SREP onset latency, resulting in greater separation of the mean phases between polarities (**Figure 3D**, Rayleigh statistic/mean resultant length R = 0.186, 0.386, 0.478, 0.597, and 0.677 at 3, 2, 1.5, 1, and 0.5 OCs before SREP onset for negative polarity, R = 0.174, 0.352, 0.458, 0.583, and 0.662 for positive polarity). At 1 OC before SREP onset latency, the mean phase values associated with positive SREPs were near ±*π*, corresponding to the trough of the oscillation, whereas the mean phase values associated with negative SREPs were near 0, corresponding to the oscillation peak. These results support that the relationship between SREP polarity and the phase of pre-saccadic oscillation is spatially consistent.

The phase–polarity relationship suggests a broader saccadic modulation of neural oscillations. Active sensing theory proposes that neuronal oscillations create cyclic windows of efficient sensory processing(*30*, *31*). In this framework, saccades may align oscillatory phase to place visual sampling within an optimal phase window or increase oscillatory power to enhance local population synchrony. If saccades align the oscillatory phase, we expect higher inter-trial phase coherence (ITPC) at saccade onset than at baseline. For both polarities, ITPC increased monotonically from 2 OCs before SREP onset latency, then decreased after SREP peak time (**Figure 3E**). The effect sizes (i.e., Cohen’s d) between ITPC at 2 OCs before SREP onset latency and subsequent 4 time points (from 1 OC before SREP onset latency to 1 OC after SREP peak time) were 1.214, 1.625, 1.739, 1.704, and 1.427. In addition, oscillatory power increased from 2 OCs before the SREP onset latency and remained elevated at that level throughout the SREP period (**Fig. S8F**). We were next interested in whether phase alignment and oscillatory power during SREPs reflect dissociable neural processes. If they were dissociable, we would expect their temporal dynamics to differ. Thus, we assessed their correlation at 5 timepoints relative to SREP onset latency and found a strong positive correlation between changes in oscillatory power and ITPC at SREP onset latency (**Figure S8H**, *R*^2^ = 0.274). The correlation was weaker at time points preceding and following SREP onset, with *R*^2^ = 0.022 at 1 OC before SREP latency, *R*^2^ = 0.118 at 0.5 OCs before SREP latency, *R*^2^= 0.198 at the SREP peak, followed by a decline to *R*^2^ = 0.046 at 0.5 OCs after the SREP peak. These results show partially divergent temporal profiles of phase alignment and oscillatory power.

Together, our results demonstrate a systematic relationship between pre-SREP oscillation phases and SREP polarity, along with an interplay between ongoing oscillations and SREP.

### Temporal lobe SREPs exhibit the earliest onsets and the highest peak amplitudes

To understand the large-scale organization of SREPs, we sought to characterize the spatial structure in SREP onset latency throughout the brain. One precondition for the presence of such a structure is greater inter-contact (i.e., across-location) variability in these two SREP parameters than trial-by-trial (intra-contact) variability. Because we hypothesized that SREPs contribute to visual memory encoding, we expected non-trivial intra-contact variability that would explain variance in subsequent recognition of the presented images. Therefore, we quantified intra-contact and inter-contact variability in SREP onset latency (see **Supplementary Materials:** Quantifying intra- vs inter-contact variability of SREP characteristics for additional details). The average intra-contact standard deviation of SREP latencies was 5.3 ms for both polarities (**Figure 4A**), whereas the mean time-interval between peak time and latency was 38.6 ms (SD = 12.3 ms). Inter-contact standard deviation of SREP onset latencies was 7.1 ms for negative SREPs and 6.6 ms for positive SREPs, both significantly greater than intra-contact variability (**Figure 4A**, N = 786, W statistics = 24141, p < 0.001, rank-biserial correlation = -0.766 for negative polarity; N = 797, W statistics = 34836, p < 0.001, rank-biserial correlation = -0.669 for positive polarity; Wilcoxon signed-rank test). This finding shows that, despite variability in SREP onset latency across saccades, greater variability arises across regions, suggesting a systematic spatial structure in SREP onset latency.

**Figure 4.**
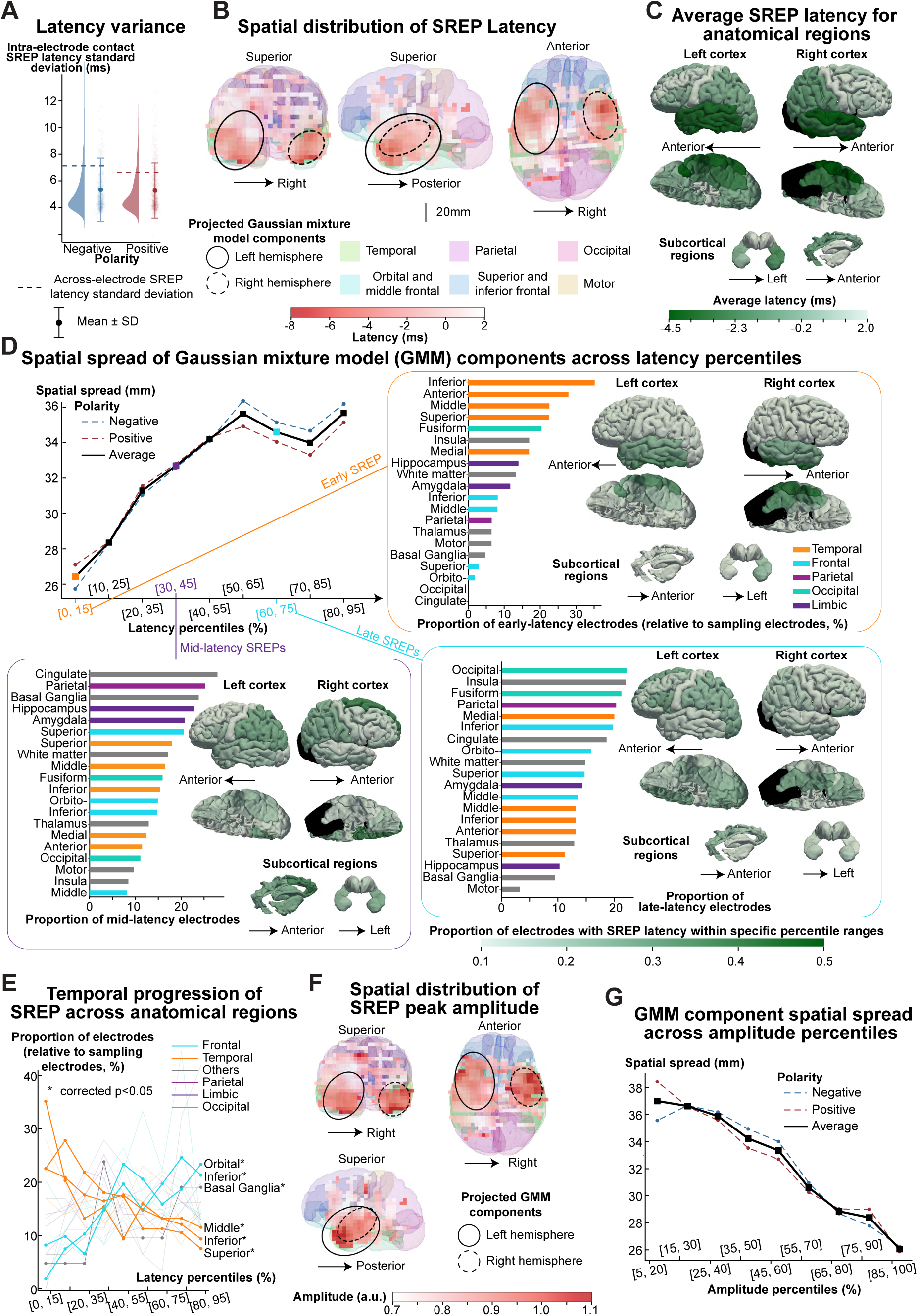
SREPs show a structured spatiotemporal organization. **(A)** Comparison between intra-contact and inter-contact SREP onset latency variance. The averaged intra-electrode-contact standard deviation of SREP onset latencies was 5.3 ms for both negative and positive SREPs. The intra-contact standard deviation of SREP onset latencies was significantly lower compared to the intra-contact standard deviation (7.1 ms and 6.6 ms for negative and positive SREPs, N = 786, W statistics = 24141, p < 0.001, Rank-biserial correlation = -0.766 for negative polarity; N = 797, W statistics = 34836, p < 0.001, Rank-biserial correlation = -0.669 for positive polarity; Wilcoxon signed-rank test). To estimate intra-contact variance in SREP onset latency, we included single trials whose SREP waveforms were similar to the averaged SREP waveform, defined as having a correlation coefficient above the median. We resampled the single-trial SREP onset latency for each contact to the 50th percentile of the trial count for all electrode contacts. To estimate the inter-contact standard deviation in SREP latencies, we resampled electrode contacts to equalize sample sizes, matching the median per-contact trial count. We repeated the process 100 times and plotted the mean inter-contact standard deviation in SREP latencies using dashed lines. **(B)** Spatial distribution of SREP latencies across electrode contacts. Similar to **Fig. S5**, we projected the 3D brain surface and electrode coordinates onto a 2D plane. We overlay a heatmap to represent the spatial pattern of SREP latencies. To generate the heatmap, we discretized this 2D space into a regular grid with a 5 mm resolution. We computed the local average SREP latency within a 10-mm radius for each grid element. Heatmap elements that have no nearby sampling electrodes are shown as fully transparent. To analyze the spatial dynamics of SREP over time, we stratified electrodes by SREP latency percentiles. For each percentile-defined subset, we applied a 3D Gaussian Mixture Model (GMM) to identify two spatial clusters of electrode contacts. We extracted the cluster centroids in the left and right hemispheres. We quantified the spatial spread based on standard deviations along the x-, y-, and z-axes. The solid and dashed ellipses represent the two GMM components identified from electrode contacts exhibiting SREP latency in the 0–15^th^ percentile range. **(C)** Average SREP latency across anatomical regions. We parcellated the brain into subregions, detailed in Figure 2. Colors denote the mean SREP latency, averaged across polarities and electrode contacts localized to each anatomical region. **(D)** Relationship between spatial spread and SREP onset latency percentile ranges. We quantified spatial spread as the square root of the sum of variances along the x-, y-, and z-axes (i.e., the Euclidean norm of the standard deviations). For illustration, we selected early, mid, and late percentile ranges ([0–15], [30–45], and [60–75]) and plotted the proportion of electrodes showing SREP with onset latency within each range. We calculated the proportion as the ratio of the number of electrodes in a region with latency within a given percentile range to the total number of electrodes localized in that region. The accompanying 3D brain plots show the anatomical distribution of these proportions across regions. Colors do not imply strict lobar classification. The fusiform gyrus is shown using the occipital color scheme, although anatomically it spans both the occipital and temporal cortices. **(E)** Sequential progression of SREP across functional-anatomical regions. Each data point represents the proportion of electrodes in a functional-anatomical region that showed SREP within a specific percentile range of onset latency. Thick lines represent regions where the proportion changed monotonically across latency percentiles based on Spearman’s correlation. Middle (*ρ* = −0.97, corrected *p* < 0.001), inferior (*ρ* = −0.74, corrected *p* = 0.455), and superior (*ρ* = −0.98, corrected *p* < 0.001) temporal gyri showed high and low proportions of early and late SREPs, respectively. By contrast, orbitofrontal (*ρ* = 0.82, Bonferroni corrected asymptotic p = 0.136, number of comparisons = 20) and inferior frontal (*ρ* = 0.89, *corrcted p* = 0.025) gyri had a low and high proportions of early and late SREPs, respectively, indicating temporal to frontal lobe progression. **(F)** Spatial distribution of SREP peak amplitude across electrodes. **(G)** Relationship between spatial spread and SREP peak amplitude percentile ranges.

Based on cluster-level SREP onset latency, electrode contacts with the earliest SREPs were clustered in the temporal lobe (**Figure 4B**). Gaussian Mixture Model (GMM) analysis revealed two predominant clusters in the left and right temporal lobes, with the earliest SREP localized to the temporal neocortex, including the inferior, basal, and lateral temporal gyri. SREP onset latency averaged within anatomical regions was earlier in the bilateral temporal lobes than in extra-temporal brain regions (**Figure 4C**). In addition, this spatiotemporal pattern was similar for SREPs of both polarities (**Fig. S9E–F**). These results quantitatively confirm that within the cerebral locations we sampled, SREPs occur earliest in the temporal lobes.

Given that SREPs were widespread and occurred earliest in the temporal neocortex, we next asked whether later SREPs were organized as a progressive expansion from the temporal neocortex into a broader distributed network. If temporal SREPs mark the beginning of a broader saccadic modulation, then later-latency SREPs should become more spatially dispersed. To test this, we stratified electrode contacts by SREP onset latency percentiles and quantified spatial spread as the Euclidean norm of the standard deviations of contact coordinates along three orthogonal axes (Left-right, posterior-anterior, and inferior-superior). Spatial spread increased progressively from early to late SREPs (**Figure 4D, Fig. S9D**). The earliest SREPs, with onset latencies in the 0–15^th^ percentile bin, were overwhelmingly observed in the bilateral temporal lobes and insula (**Figure 4D**). We observed mid-latency SREPs (30–45^th^ percentile bin) in the parietal lobe, subcortical regions (basal ganglia and thalamus), and the medial temporal lobe (amygdala and hippocampus). Late SREPs (60–75^th^ percentile bin) were more broadly distributed. These findings suggest that widespread SREPs follow a spatial progression from temporal lobes toward broader cortical and subcortical regions.

We then characterized the involvement of functional-anatomical regions in this progressive saccadic modulation. For each latency percentile bin, we quantified the proportion of SREP contacts located in each anatomical region. Consistent with our prior findings that temporal saccadic modulation was earliest, temporal regions accounted for a larger proportion of contacts in early-latency bins, and this proportion declined as SREP onset latency increased (**Figure 4E**). This decline was strongest in the middle temporal gyrus (Spearman’s ρ = -0.97, Bonferroni-corrected p < 0.001, number of comparisons = 20) and superior temporal gyrus (ρ = -0.98, corrected p < 0.001). The inferior temporal gyrus followed the same trend (ρ = -0.74, corrected p = 0.455). In contrast, frontal regions were more involved at a later stage of saccadic modulation. We found a significant positive correlation between the latency percentile bin (ordered from earliest to latest) and the proportion of electrodes whose SREP onset latency fell into that bin in the inferior frontal (ρ = 0.89, corrected p = 0.025) and orbital frontal gyri (ρ = 0.82, corrected p = 0.136).

Previous analyses suggest that the temporal neocortex may be the source of saccadic modulation. If this is true, then the temporal cortex should not only show the earliest SREPs, but also the strongest SREPs. Cluster-level average SREPs with the highest peak amplitudes averaged across polarities were also clustered in the bilateral temporal lobes (**Figure 4F**, **Fig. S10G,** see **Supplementary Materials** for intra- and inter-contact variability in SREP peak amplitude). The spatial spread of stratified electrodes by percentile bins of absolute peak amplitude decreased monotonically with increasing saccadic modulation strength, measured by absolute SREP peak amplitude (**Figure 4G**). Stronger SREPs formed localized spatial clusters in the temporal lobes, whereas SREPs with lower absolute peak amplitudes were more spatially dispersed **(Fig. S10E**). Region-wise correlation analysis between the peak amplitude percentile bin and the proportion of electrodes whose SREP peak amplitude fell in that bin confirmed a spatial progression of saccadic modulation: significant positive correlation for the temporal pole (**Fig. S10F,** *ρ* = 0.95, Bonferroni-corrected *p* = 0.002, number of comparisons = 20), inferior temporal (*ρ* = 0.95, corrected *p* = 0.001), and middle temporal (*ρ* = 0.93, corrected *p* = 0.005) gyri.

In summary, SREPs exhibit a spatially structured progression, with earlier and stronger SREPs recorded in the temporal cortex, particularly the neocortex, followed by later involvement of broader cortical and subcortical regions.

### Saccade-related neural dynamics (SRNDs) predict successful visual memory encoding

Our findings suggest SREPs were accompanied by coordinated pre-saccadic oscillatory activity; we therefore refer to their combination as saccade-related neural dynamics (SRNDs). To make our central hypothesis that SRNDs predict visual memory encoding testable, we performed dimension reduction. Because each SREP waveform is a high-dimensional time series, mapping the full waveform to memory outcomes would be computationally inefficient and difficult to interpret biologically. Therefore, we defined a physiologically meaningful model of SRND for each saccade at each location using five parameters: SREP onset latency, SREP peak-to-trough amplitude, SREP peak time, and the power and phase of the ongoing oscillation at half OCs before saccade onset (**Figure 5A**). We then employed random forest classifiers to test whether SRNDs predict memory encoding(*32*). The random forest classifiers were trained on SRND parameters for individual electrode contacts, with each saccade labeled according to the subsequent memory outcome of the trial in which the saccade occurred. This electrode-contact-level model learned location-dependent SRND-memory mappings, which formed the basis of a hierarchical prediction framework. Because we simultaneously recorded SRNDs from multiple electrode locations, and multiple saccades could occur during each image presentation, we can aggregate predictions across electrodes to yield saccade-level predictions, and then across saccades to yield trial-level predictions. This hierarchical approach enabled us to address three distinct questions about how SRNDs support memory encoding: (1) Are SRNDs at a single electrode location sufficient to predict memory outcome? (2) Is memory encoding jointly shaped by SRNDs across the brain? If so, aggregating prediction probabilities across electrode locations should improve prediction; and (3) Does aggregating predictions across saccades further improve the prediction of the memory outcome of the viewed image? We address each of these questions in turn below.

**Figure 5.**
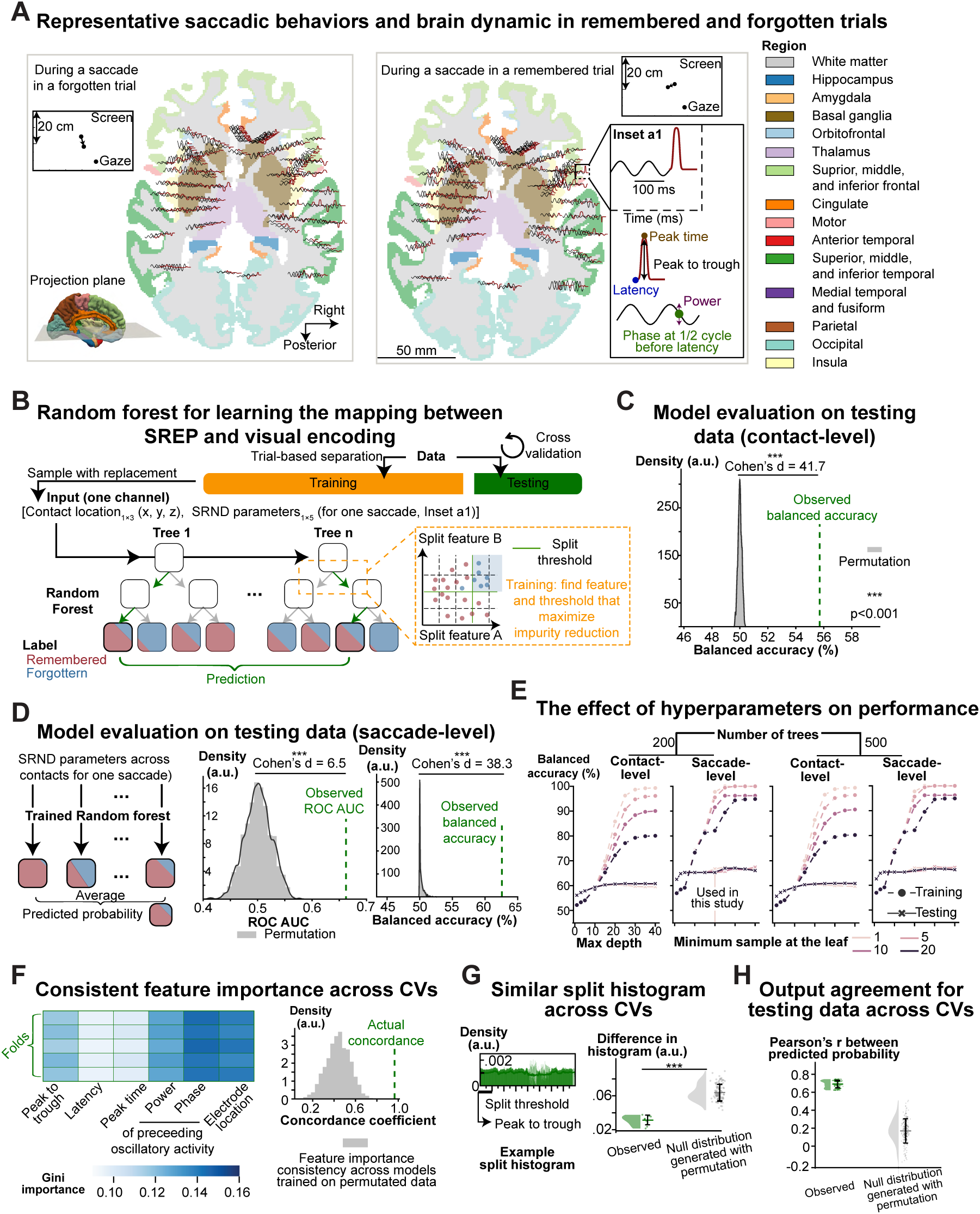
Machine learning reveals that SREPs predict visual memory encoding. **(A)** Illustration of brain dynamics during a remembered and a forgotten trial. Each trace plotted on top of a horizontal brain slice represents the SREP waveform and the preceding oscillatory activity recorded in an electrode contact. The color of the brain slice denotes anatomical regions. We reconstructed SREP waveforms as sigmoidal rises and falls based on the SREP onset latency, peak time, and peak-to-trough amplitude. We reconstructed the preceding oscillatory activity using a 10 Hz sine wave parameterized by instantaneous power and phase at half an oscillation cycle before the SREP onset latency. We later trained linear and nonlinear models to predict visual encoding using these five saccade-related neural dynamic (SRND) parameters (SREP onset latency, peak-to-trough amplitude, peak time, and the phase and power of preceding neural oscillation) at each location (see text for details). The two insets at the top show fixation before and after a saccade, with arrows indicating saccade direction. **(B)** Model architecture for learning the mapping from SRND parameters to visual encoding. The input to the random forest was a vector including the spatial locations of electrode contacts and SRND parameters. As the remembered and forgotten trials were imbalanced, we resampled both classes to achieve a balanced input. In addition, we added normally distributed spatial jitter to the electrode contact locations to enable learning that is specific to spatial location while preventing the exact electrode contact location from being associated with memory outcomes. For prediction, each decision tree outputs a probability for the remembered and forgotten classes, reflecting the proportion of training samples from each class that reach the terminal leaves. The final predicted probability is the average of these vectors across all trees. **(C)** Balanced accuracy of the random forest in predicting visual encoding performance on testing data. We performed 200 permutations to generate a null distribution of balanced accuracy. The observed balanced accuracy was 55.7%, exceeding all values in the null distribution. **(D)** Saccade-level model performance. Using these saccade-level predicted probabilities, we computed the area under the receiver operating characteristic curve (ROC AUC). An illustration of how to calculate ROC AUC is shown in Figure 2C. The observed ROC AUC on test data (0.662) exceeded all values from a null distribution generated by training and evaluating models on data with permuted labels. The rightmost panel shows the observed saccade-level balanced accuracy (62.6%), compared to the null distribution. To calculate the saccade-level balanced accuracy, we determined the optimal threshold, sweeping from 0 to 1, that maximized balanced accuracy. We applied this process to both observed data and data with permutated labels. **(E)** Effect of hyperparameters on model performance during training and testing. We evaluated how different hyperparameter settings affected the performance of random forests. The hyperparameters include the number of trees, the maximum tree depth, and the minimum sample size at a leaf. In general, deeper trees with a small sample size can learn more complex patterns but risk overfitting. In each panel, color indicates the minimum sample size at the leaf, the x-axis shows the maximum depth, and the y-axis shows balanced accuracy. From left to right, the four panels show: (1) electrode contact-level performance using 200 trees, (2) saccade-level performance using 200 trees, (3) contact-level performance using 500 trees, and (4) saccade-level performance using 500 trees. In this study, we used the default hyperparameter settings: 200 decision trees, a maximum depth of 20, and a minimum leaf size of 1. This set of hyperparameters achieved moderate balanced accuracy during training and moderate generalization on the test data, with no evidence of overfitting. Therefore, we used the default parameters for consistency and computational efficiency, although performance on test data slightly improved with 500 decision trees. **(F)** Similar mapping from SRND parameters to visual encoding outcome across cross-validation folds. The left panel shows feature importance based on Gini impurity reduction. We used Kendall’s coefficient of concordance to quantify the concordance of feature importance across CVs and across permutated data. The right panel shows that the concordance coefficient across CVs (Kendall’s W = 0.966) for the actual data exceeds all values in the null distribution (mean Kendall’s W = 0.439, SD = 0.110). **(G)** Consistent decision boundaries across cross-validation folds. For each model trained on the actual data, we computed the histogram of split thresholds for every feature. To assess consistency, we compared split histograms between pairs of folds and quantified their similarity using the area under the curve of absolute difference. The right panel shows that the split histogram for models trained on actual data across CVs was more similar than the null distribution generated based on permuted data (U = 0.000, p < 0.001, Mann-Whitney U test, Cohen’s d = -3.398, *n_actual_* = 10, *n_null_* = 100). **(H)** Consistent predictions of random forest models across CV folds. To assess the output agreement, we randomly sampled an observation from the test set (in one of the folds) and passed it to two models trained from different CV folds. We repeated this procedure 200 times, both for models trained on actual data and on models trained on permuted data. We used Pearson’s correlation coefficient between the predicted probabilities of a pair of models to quantify the agreement in their outputs. Each data point in the plot represents one such correlation. Models trained on actual data had higher output agreement across CVs than those trained from permuted data (U = 40000, p < 0.001, Mann-Whitney U test, Cohen’s d = 5.208, *N_actual_* = 200, *N_null_* = 200).

In 5-fold cross-validation (CV), the trained model achieved 55.7% balanced accuracy and 70.3% accuracy at the electrode-location level, exceeding all values in the null distribution obtained by permuting the memory-encoding outcome labels (**Figure 5C, Fig. S11A**). These results show that SRNDs at individual electrode locations carry information predictive of visual encoding.

To assess whether memory encoding depends on coordinated SRNDs across locations, we evaluated the predicted probability of successful memory encoding at the saccade level. Saccade-level prediction was above chance. The observed area under the ROC curve on test data (0.662) exceeded all values from its null distribution generated by training and evaluating models on data with permuted labels (**Figure 5D**). The observed saccade-level balanced accuracy corresponding to the optimal decision threshold was 62.6%, exceeding all values in its permutation-based null distribution (**Figure 5D**). Moreover, for each CV fold, both electrode-contact- and saccade-level balanced accuracy exceeded all values in their respective permutation-based null distribution (**Fig. S11F**). These results show that SRNDs across the brain jointly contribute to memory encoding.

To assess whether aggregating predictions across saccades further improves the prediction of the memory outcome, we computed the trial-level average predicted probability from SRND parameters across all electrode contacts and all saccades. The trial-level predicted probability for testing data was higher in remembered trials than in forgotten trials (**Fig. S11C**, U statistic = 84652, p < 0.001, rank-biserial correlation = 0.333, *N_remembered_* = 635, *N_for_*_g*otten*_ = 200, Mann–Whitney U test). At the trial level, the observed testing area under the ROC curve was 0.678, and balanced accuracy was 63.1%, both exceeding all values in their respective null distributions (**Fig. S11B**, permutation test). Compared to the saccade-level model performance, aggregating SRND parameters across multiple saccades in a trial did not substantially improve memory outcome prediction.

This latter finding suggested that successful memory encoding may not require that every saccade during image viewing be accompanied by encoding-conducive SRNDs. Because each image is sampled through a sequence of saccades, SRNDs may influence memory cumulatively(*33*). We therefore asked whether remembered images were associated with a higher proportion of saccades with SRNDs classified as encoding-conducive. Indeed, remembered trials contained a larger fraction of such saccades than forgotten trials (81% versus 56%; **Fig. S11D**), and predicted saccade labels were significantly associated with trial outcome (χ²(1) = 242.861, p < 0.001). To account for the potential stochasticity in random forest classifier training (i.e., random feature subsampling at each split), we repeated 5-fold CV 100 times. All iterations showed a 25–27% higher proportion of encoding-conducive SRNDs in remembered than forgotten trials. These results clearly show that remembered trials had a greater proportion of SRNDs associated with successful memory encoding.

To ensure that the trained random forest classifier was neither underfitting nor overfitting, we systematically evaluated its performance across a range of hyperparameters. Classifiers with greater complexity showed improved training balanced accuracy until saturation, and testing balanced accuracy increased in parallel without subsequent decline (**Figure 5E**). Under the hyperparameter configuration used in this study, the random forest classifier achieved both higher training and testing balanced accuracy than other configurations, with no evidence of divergence between them across the surrounding hyperparameter space. Overall, these results establish SRNDs as a physiological mechanism that supports visual memory encoding.

### Earlier and greater-amplitude SREPs predict recognition memory

Having confirmed that SRNDs predict recognition memory, we sought to understand the physiological features that are associated with successful memory encoding. Therefore, we applied the Shapley additive explanation framework to the trained random forest classifiers, which assigns each feature a contribution to the model-predicted probability of successful encoding(*34*). To ensure generalizable interpretation of the SRNDs-memory mapping, we tested whether the mapping learned by the classifier across CV folds was consistent. For this purpose, we quantified (1) the concordance of feature importance, (2) the similarity of decision boundaries, and (3) the agreement of model outputs across CV folds.

The observed concordance of feature importance across CV folds (Kendall’s concordance coefficient W = 0.966) was higher than all values in the null distribution (mean Kendall’s W = 0.439, SD = 0.110), suggesting that the models across CVs consistently identified the same features as important and weighted them similarly (**Figure 5F**).

To examine whether random forest classifiers’ across CV folds used similar decision boundaries, we summarized the distribution of split thresholds across features using split histograms. If classifiers learned a stable SRND-memory mapping, these split histograms should be more similar across folds than those derived from permutated data. We quantified between-fold dissimilarity as the area under the absolute difference curve between split histograms. The split histograms for models trained on actual data across CVs were more similar than the null distribution generated based on permuted data (U = 0.000, p < 0.001, Mann-Whitney U test, Cohen’s d = -3.398, *N_actual_* = 10, *N_null_* = 100), suggesting that random forests trained on actual data exhibit consistent decision boundaries across CV folds (**Figure 5G**).

To assess agreement between model outputs, for each pair of models trained on different CV folds, we randomly sampled observations not included in either model’s training and calculated Pearson’s correlation coefficient between the predicted probabilities. Models trained on actual data had higher output agreement across CV folds than those trained from permuted data (**Figure 5H**, U = 40000, p < 0.001, Mann-Whitney U test, Cohen’s d = 5.208, *N_actual_* = 200, *N_null_* = 200). Additionally, the proportion of testing samples with different predictions was significantly lower than the null distribution derived from models trained with permuted data (**Fig. S11H**, U = 0, p < 0.001, Mann-Whitney U test, Cohen’s d = -7.656, *N_actual_* = 200, *N_null_* = 200). Together, these results demonstrate that the mapping learned by the random forest classifiers between SRND parameters and memory outcome generalizes across CV folds.

To characterize SRND profiles associated with successful memory encoding, we first verified that the derived Shapley values capture our random forest’s decision structure for predicting memory outcomes, and then constructed an interpretable representation of the relationship between SRND parameter values and Shapley values. Shapley values quantify how much a given feature value for an individual observation increases or decreases the probability of successful memory encoding, while accounting for correlations among features (**Figure 6A**). In this study, the SRND parameter–Shapley value relationship was consistent with partial dependence plots (**Figure 6B**), which summarize the average effect of each SRND parameter on predicted probability, and model-predicted probabilities across SRND parameter values (**Figure 6C**).

**Figure 6.**
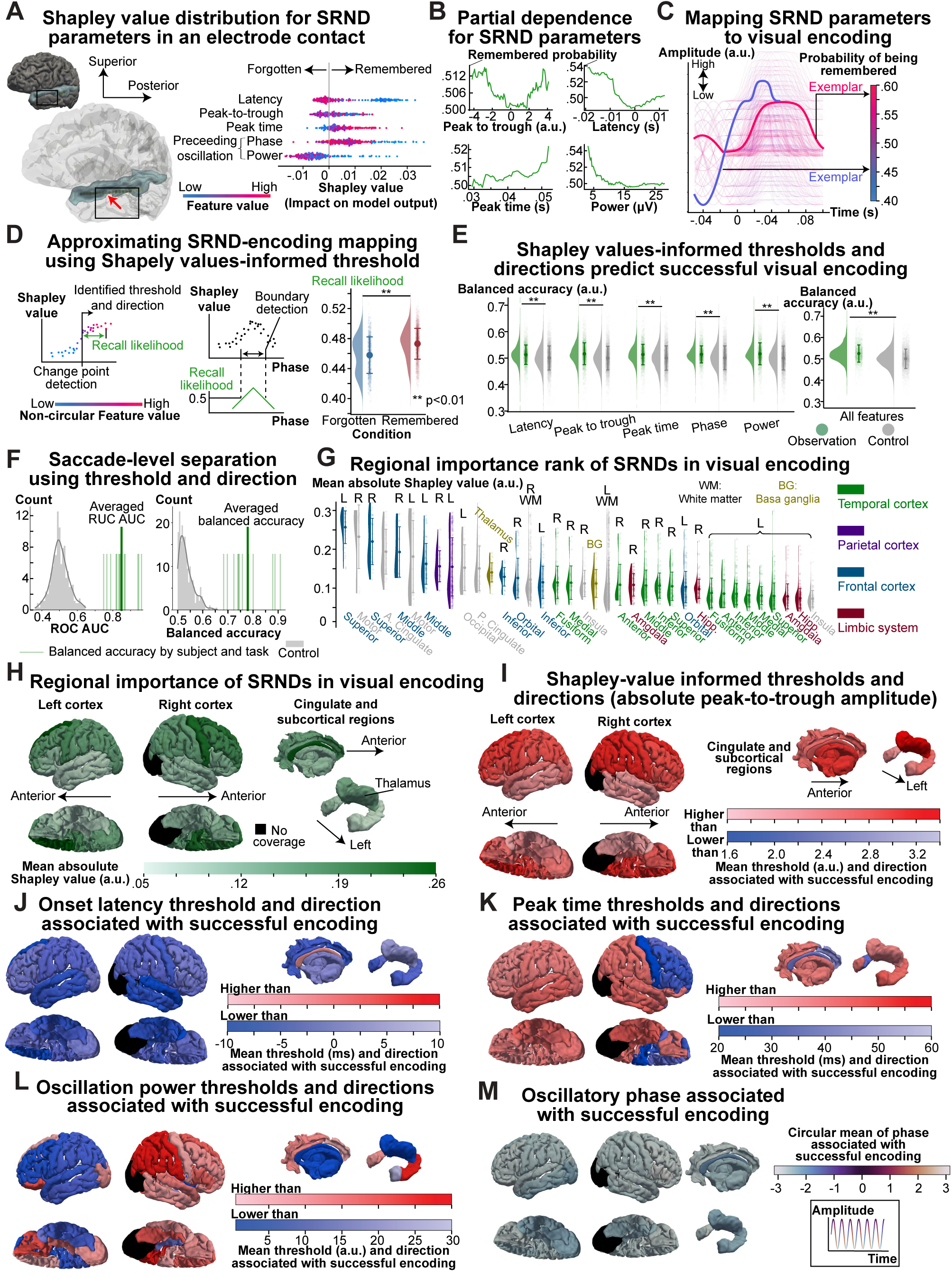
Explanatory analysis of the mapping between saccade-related neural dynamic (SRND) model parameters and visual encoding. **(A)** Illustration of how Shapley values explain the learned relationship between SRND parameters and visual memory encoding. This panel shows the Shapley values for SRND parameters recorded at a representative electrode contact in the superior temporal gyrus. Each point represents the Shapley value of a feature value, quantifying how much this feature value contributed to the model’s (e.g., random forest model) prediction. The color represents the relative feature value, mapped separately for each feature from low to high. A positive Shapley value indicates that the feature value increased the predicted probability of successful visual encoding. **(B)** Partial dependence plots for SRND parameters recorded in the same electrode contact are shown in panel A. Each curve represents the marginal effect of a feature on the predicted probability of successful visual encoding by averaging out the effects of all other features in the model. This panel illustrates how each feature influences a model’s prediction, aiming to provide a better understanding of the Shapley values presented in panel A. For example, an earlier SREP (lower latency) is associated with a higher Shapley value, which is confirmed by the partial dependence plot in this panel. **(C)** Reconstructed SRND model and associated probability of successful encoding for the electrode contact shown in panel A. We systematically varied SRND parameters, including instantaneous phase and power at 0.5 OCs before SREP onset latency, SREP onset latency, peak-to-trough amplitude, and peak time. We input the feature combinations and the electrode contact’s spatial coordinates into the trained random forest model to obtain the predicted probability of successful visual encoding. Each curve represents the reconstructed SREP waveform combined with preceding oscillatory activities. We reconstructed the preceding oscillatory activities using a 10Hz sinusoidal wave, with instantaneous phase and power measured at half a cycle before the SREP latency. We reconstructed the SREP using two sigmoid functions: one for the rising phase, beginning from the (SREP onset latency, oscillation amplitude at the SREP onset latency) to (SREP peak time, SREP peak-to-trough amplitude); and one for the falling phase from the peak time onward. Reconstructed SREP and preceding oscillation are color-coded by their predicted encoding probability. **(D)** Location-specific interpretation of the learned mapping using Shapley value-informed thresholds and directions. The left panel schematically illustrates how thresholds and encoding-favorable directions were derived from Shapley values. The right panel shows the distribution of recall likelihood, computed from the derived thresholds and directions and averaged across electrode contacts, for saccades occurring during remembered and forgotten trials (*N_remember_* = 3052, *N_forgotten_* = 906, U = 1.872 × 10^6^, p < 0.001, Mann-Whitney U test, rank-biserial correlation coefficient = 0.354). For non-circular features, such as absolute peak-to-trough amplitude, we ordered observations by feature values. The left plot shows the Shapley values over the ordered feature values. We detected a feature value that minimized the total sum of squared differences between Shapley values and their respective means within segments separated by this value. For the circular feature, the instantaneous phase of the preceding oscillatory activities, we searched for a window (e.g., two boundaries on the unit circle) that minimized the sum of squared Shapley value differences from the mean within and outside the window. For each feature, we defined the direction associated with successful encoding by comparing the average Shapley values on either side of the threshold. For the circular feature, we compared Shapley values inside and outside the identified window. To confirm that the Shapley value-informed thresholds and directions explain visual encoding, we calculated recall likelihood, a probability-like measure of successful encoding based on the thresholds and directions. For each observation, we computed recall likelihood as the distance between the feature value and the threshold, scaled by the standard deviation of the feature values. Depending on the Shapley value-informed direction, we added or subtracted the scaled distance from the prior probability (i.e., 0.5) and then clipped the result to the range [0, 1]. For circular features, the same logic was applied using the shortest angular distance between a given feature value and the boundaries of the encoding-favorable window. **(E)** Balanced accuracy of distinguishing between remembered and forgotten trials using recall likelihoods computed from Shapley value-informed thresholds and directions. For each saccade, we computed the recall likelihood of each SRND parameter at each electrode contact and compared it with a baseline probability of 0.5. The left plot shows the distribution of balanced accuracy using actual and control Shapley value-informed thresholds and directions. We generated the control thresholds and directions by randomly sampling thresholds from the observed feature values. For all SRND parameters, balanced accuracy was higher when using recall likelihood based on actual Shapley value-informed thresholds and directions (*N_observation_* = 2707, *N_control_* = 5414, p < 0.001, t-test, Cohen’s d ranged from 0.296 to 0.390). The right plot shows the balanced accuracy for separating successful from unsuccessful visual memory encoding using recall likelihood for all features (*N_observation_* = 2707, *N_control_* = 5414, U = 9.767 × 10^6^, p < 0.001, Mann-Whitney U test, Cohen’s d = 0.571). These results suggest that Shapley value-informed thresholds and directions reflect the cognitive state associated with successful visual encoding. We considered an observation of SREP at an electrode contact to be associated with successful visual encoding if more than half of the feature values exceeded the 0.5 baseline probability. **(F)** Saccade-level separation between remembered and forgotten trials using Shapley value-informed recall likelihood. For each saccade, we calculated the proportion of SRND parameters associated with successful visual encoding across electrode contacts. We generated the receiver operating characteristic (ROC) curve by varying the proportion threshold. The left plot shows the distribution of area under the ROC curve (AUC), and the right plot shows the distribution of balanced accuracy at the optimal proportion threshold. These two plots show saccade-level separation between remembered and forgotten trials using Shapley value-informed thresholds and directions, versus thresholds randomly sampled from the feature values. Individual green lines represent the observed AUC or balanced accuracy for each subject and each task. For each plot, the thicker green line represents the averaged observed AUC (84.9%) or balanced accuracy (77.9%). The rank-biserial correlation was 1.000, and Cohen’s d was 6.789 for the averaged AUC compared to the control distribution. Rank-biserial correlation was 1.000, and Cohen’s d was 8.761 for balanced accuracy compared to the control distribution. **(G)** Contribution of SRND parameters to visual encoding across anatomical regions. To quantify the regional contribution of SRND parameters to visual encoding, we computed the mean absolute Shapley value for each anatomical region by averaging across electrode contacts within this region and across all SRND parameters. A higher mean absolute Shapley value indicates that the SRND parameters at this region have a greater influence on visual encoding performance. **(I–M)** Spatial distribution of SRND parameters associated with successful visual encoding across anatomical regions. The five panels visualize the Shapley value-informed directions and thresholds for SRND parameters associated with successful visual encoding at each brain region. We used two color schemes. The red color map (light to dark red) indicates regions where higher feature values were associated with successful encoding. The blue colormap (light to dark blue) indicates regions where lower feature values were associated with successful encoding. The colors in the two color-maps reflect the threshold values, with red and blue corresponding to the most extreme thresholds. For example, a red region indicates that feature values above a relatively high threshold are associated with successful visual encoding. For the instantaneous phase at 0.5 OCs before SREP onset latency, which is circular, the color denotes the angular mean of the window of phase associated with successful visual encoding. We color-coded a sinusoidal signal by phase to facilitate interpretation. The orientation of the 3D brain models matches the orientation annotated in panel **I**.

To render the SRND parameter value–Shapley value relationship interpretable, we constructed a lower-dimensional representation of this relationship for each parameter and each electrode contact. This representation consisted of a threshold and a direction derived from Shapley values, partitioning each SRND parameter into encoding-conducive and non-encoding-conducive ranges (see **MATERIALS AND METHODS:** Shapley value-informed thresholds and directions); therefore, it reduces the decision trees in the trained random forest classifier to a single decision boundary per parameter and location (**Figure 6D**).

We then assessed whether Shapley value-informed thresholds and directions captured a meaningful relationship between SRND parameters and memory outcome. For this purpose, we compared recall likelihood between remembered and forgotten trials, a probabilistic measure of successful memory encoding derived from Shapley value-informed thresholds and directions (see **MATERIALS AND METHODS:** Shapley value-informed thresholds and directions). We found that recall likelihood averaged across electrode contacts was significantly higher in remembered trials compared to forgotten trials (**Figure 6D**, *N_remembered_* = 3052, *N_for_*_g*otten*_ = 906, U = 1.872 × 10^6^, p < 0.001, Mann-Whitney U test, rank-biserial correlation coefficient = 0.354). We next assessed whether Shapley value-informed thresholds and directions predicted successful memory encoding. To assess whether this prediction exceeded what would be expected from arbitrary thresholds, we performed surrogate testing by randomly sampling thresholds from the observed feature values. For each saccade, we compared the recall likelihood of every SRND parameter at every electrode contact against a baseline probability of 0.5. The balanced accuracy for individual SRND parameters based on actual Shapley value-informed thresholds and directions was significantly higher than the surrogate control (*N_observation_* = 2707, *N_control_* = 5414, p < 0.001, t-test, Cohen’s d ranged from 0.296 to 0.390). When we leveraged recall likelihoods across all SRND parameters for each saccade (i.e., classifying an instance of SRND as encoding-conducive if the majority of parameters exceeded 0.5), the actual balanced accuracy was significantly higher than the control (*N_observation_* = 2707, *N_control_* = 5414, U = 9.767 × 10^6^, p < 0.001, Mann-Whitney U test, Cohen’s d = 0.571). **Fig. S12A** shows the area under the ROC curve for classifying whether SRNDs recorded at an electrode contact during a saccade occurred in a remembered or forgotten trial, based on recall likelihoods. The area under the ROC curve was significantly higher using recall likelihood based on actual Shapley value-informed threshold and direction (*N_observation_* = 2707, *N_control_* = 5414, p < 0.001, Mann-Whitney U test, U = 1.067 × 10^7^, Cohen’s d = 0.817), compared to the surrogate control. These results demonstrate that Shapley value-informed thresholds and directions provide an interpretable summary of the SRNDs associated with successful visual memory encoding.

We found that SRNDs across the brain better predict memory outcome than SRNDs in a single location. If the Shapley value-informed thresholds and directions accurately identify the range of encoding-conducive SRND parameters at each location, then the extent to which encoding-conducive SRNDs are present across locations should better predict, for each saccade, whether the viewed image was recognized. To test this hypothesis, we calculated, for each saccade, the proportion of SRND parameters that fell within the Shapley value–informed, encoding-conducive ranges across all electrode locations. To evaluate the prediction performance, we constructed an ROC curve by varying the proportion threshold. The averaged area under the ROC curve across subjects was 84.9%, exceeding all values in the control distribution generated by randomly sampling thresholds from the feature values (**Figure 6F**, rank-biserial correlation = 1.000, Cohen’s d = 6.789). The average balanced accuracy at the optimal proportion threshold across subjects was 77.9%, which also exceeded all values in the control distribution (rank-biserial correlation = 1.000, Cohen’s d = 8.761). Notably, Shapley values–informed thresholds and directions better predicted successful visual encoding at the saccade level than the random forest classifier trained on SRND parameters (**Figure 5D**). This result shows that the threshold–direction representation, by distilling the complex decision rules of the random forest into a single decision boundary, may better capture the generalized mapping between memory encoding and SRNDs.

We next used Shapley value–informed thresholds and directions to identify brain regions in which SRNDs most strongly influenced visual memory encoding. For each electrode contact, we computed the mean absolute Shapley value across saccades to quantify how strongly that contact’s SRND parameters contributed to the memory outcome. The mean absolute Shapley values for SRND parameters across CV folds were consistent with feature importance based on Gini impurity reduction (**Figure 5F**), and consistent across CV folds (**Fig. S11G**, Kendall’s coefficient of concordance = 0.966, p = 0.0, permutation test). We ranked the average values across electrode contacts within a functional-anatomical region to assess the spatial distribution of influence (**Figure 6G**). We found that SRNDs in the frontal and parietal regions had the strongest influence on visual encoding (**Figure 6H**). Regional averages may obscure within-region heterogeneity. Therefore, we visualized mean absolute Shapley values at the electrode-contact level (**Fig. S12C**), which confirmed that SRNDs in the frontal and parietal cortices and nearby white matter exerted the strongest influence on visual encoding.

Finally, we delineated the SRND profiles associated with successful visual encoding by quantifying Shapley value–informed thresholds and directions for each SRND parameter across sampled brain regions (**Figure 6I–M**, summarized in **Table 1**). SREPs with greater magnitude, particularly in the frontal and parietal regions, were associated with successful encoding (**Figure 6I**). This was consistent with the electrode contact-level SREP magnitude profile associated with successful visual encoding shown in **Fig. S12D**. Earlier SREP latencies across the cerebrum were associated with successful encoding (**Figure 6J, Fig. S12E**). Later SREP peak times in the left hemisphere were associated with successful visual encoding (**Figure 6K**, **Fig. S13A**). Oscillatory power at 0.5 OCs before SREP onset latency showed hemispheric asymmetry: lower power in the left hemisphere and higher power in the right hemisphere were associated with successful encoding (**Figure 6L, Fig. S13B**). At the same pre-SREP timepoint, oscillation in the trough-to-peak rising phase (i.e., [−*π*, 0] radians) was associated with successful memory encoding across most brain regions, regardless of SREP polarity (**Figure 6M, Fig. S13C, Fig. S14A–B**). Overall, Shapley value analyses reveal that SREP profiles marked by early latency and strong amplitude are associated with successful visual memory encoding.

**Table 1.** Summary of the identified mapping between SRND parameters and successful memory encoding. This table provides a concise summary; a more comprehensive representation of these relationships is shown in **Figure 6**.

| SRND parameters | Location | Successful-memory-encoding profile |
| --- | --- | --- |
| SREP magnitude | Cerebrum (especially frontal and parietal cortex) | Higher magnitude |
| SREP onset latency | Cerebrum (especially temporal cortex) | Earlier onset latency |
| SREP peak time | Right precentral gyrus and lateral parts of the right frontal cortex | Earlier peak time |
|  | Other parts of the cerebrum | Later peak time |
| Preceding oscillatory power | Left cortex (in general) | Lower power |
|  | Right cortex (in general) | Higher power |
| Preceding oscillatory phase | cerebrum | Trough-to-peak rising phase (i.e., $[-\pi, 0]$ radians) |

### SREPs encode either the current or subsequent saccade direction very rarely, if at all

We have demonstrated that SREPs reflect neural states supporting visual encoding and characterized SREP profiles associated with successful visual encoding. The remaining question is whether these SREPs represent saccadic corollary discharge or a novel memory-related mechanism. If SREPs were a narrowly defined corollary discharge signal, they should convey information about saccade direction. However, across held-out testing data, SRND parameters did not predict saccade direction above chance, either at the electrode-contact level or after aggregating predictions across contacts at the saccade level (**Fig. S15**, **Supplementary Materials**). This absence of directional information was robust across hyperparameter analyses and alternative modeling approaches, including neural networks, angular-loss models, von Mises kernel regression, and binary left–right direction classification (**Fig. S16A–I**, **Supplementary Materials**). SRNDs also failed to predict the direction of the subsequent saccade, arguing against a prospective motor-planning explanation (**Fig. S16J**). Together, these analyses suggest that SRNDs do not generally encode saccade direction and are unlikely to reflect a corollary discharge signal. Instead, SREPs index a saccade-related neural state that supports visual memory encoding.

## Discussion

Here, we provide direct evidence that saccade-related evoked potentials (SREPs) represent saccadic modulation of large-scale neural activity in humans. Corroborating this interpretation, we demonstrate an interplay between SREP waveforms and ongoing oscillatory neural activities throughout the brain, together termed saccade-related neural dynamics (SRNDs). Our explanatory analysis of machine learning models validates the hypothesis that SRNDs contribute to visual memory encoding. Fronto-parietal SRNDs predict successful memory encoding to a greater degree than hippocampal SRNDs. SRNDs across the brain do not predict saccade direction, indicating that their disassociation from corollary discharge. Coordination between active sensing mechanisms (e.g., saccades, sniffing, echolocation) and underlying neural activity is commonly found across species (e.g., non-human primates, rodents, and bats), presumably to optimize information processing and support navigation and foraging(*19*, *35*, *36*). Our findings demonstrate that an analogous mechanism exists to support human visual encoding: larger SREPs likely reflect a transient increase in synchronized synaptic activity, creating a widespread network state favorable for visual memory encoding. This network is spatiotemporally organized: SREPs emerge earliest and with the greatest amplitude in the lateral temporal cortex. These findings open new avenues for memory-enhancing neuromodulation, for example, through stimulation at saccade onset to amplify SREPs.

### Presence, spatiotemporal structure, and behavioral relevance of SREPs

Studies of neural mechanisms in human visual encoding have focused on brain responses to various visual stimuli(*37*, *38*). Although SREPs were previously reported and have been shown to modulate neural activity in non-human primates, direct evidence that these findings translate to humans has remained scarce(*4*, *7*, *39*). This knowledge gap persists due to methodological constraints, such as the limited availability of human intracranial LFPs and the potential contamination of ocular-muscular artifacts in non-invasive electrophysiology. We leveraged a unique research opportunity in neurosurgical patients undergoing intracranial monitoring, and our findings provide direct evidence for the presence of SREPs in humans. These results corroborate and extend a recent study by Staudigl et al., which showed that saccades coordinated amygdala-hippocampal interactions in humans(*40*). When considered alongside findings from non-human primates, the available data suggest that saccadic modulation operates at multiple physiological scales, including neuronal spiking, evoked potentials, oscillations, and functional connectivity(*8*, *36*, *41*). Likewise, we observed consistent SREPs, along with increased phase synchronization and increased oscillation power following saccades. Our findings that SREP waveforms are systematically associated with the phase of ongoing oscillations (**Figure 3**) suggest that neural dynamics across scales at the time of saccades may reflect a latent state indexing attention or readiness for visual processing. Validating this hypothesis is important because, if correct, it would unify saccade-related neural phenomena into a coherent mechanistic framework. This hypothesis makes two falsifiable predictions: (1) experimentally inducing saccades at different phases of ongoing oscillations affects other saccade-related neural dynamics, and (2) the encoding of other sensory information, such as auditory encoding, is enhanced when SREP profiles mirror those associated with successful visual encoding.

Previous studies of saccade-related neural dynamics have primarily focused on the visual system, where they have been studied as mechanisms of visual perception, more specifically, perceptual stability(*42*, *43*). More recent work has extended this scope, showing that saccades modulate neural activity beyond the visual system, including the medial temporal lobe(*2*), the thalamus(*44*), and the auditory cortex(*21*). These findings suggest saccade-related neural dynamics may serve broader cognitive functions beyond visual perception. These studies primarily focused on individual anatomical-functional regions, potentially missing coordinated saccadic modulation across the brain. Our study demonstrates widespread SREPs across the human cerebrum and reveals their systematic spatiotemporal organization. Earlier SREPs with greater amplitude emerged in the temporal neocortex, a region implicated in high-order visual processing(*45*), followed by later SREPs across parietal and frontal regions (**Figure 4–supplementary video**). This pattern suggests a cascading dynamic across regions coordinating perceptual and encoding processes over time, which aligns with emerging evidence that memory encoding is not strictly localized, but rather reflects broad activations across the brain(*46*, *47*). Two studies demonstrated that electrical stimulation of the left middle temporal gyrus, the region where we observed early high-amplitude SREPs, selectively enhanced verbal memory(*48*, *49*). Ezzyat et al. further showed that stimulation was effective when delivered during decoded intervals of poor encoding, indicating that the middle temporal gyrus dynamically gates memory encoding(*48*). SREPs identified in our study represent a naturalistic parallel: saccades may coordinate with encoding-favorable states that stimulation sought to achieve artificially(*48*).

While SREPs are most prominent in the temporal lobe, the variations of SREPs and preceding neural oscillation in the frontal and parietal lobes have a greater trial-by-trial influence on visual memory (**Figure 6G and H**). We interpret these results as follows: early SREPs likely reflect the reconfiguration of the temporal lobe network, which may facilitate high-level visual processing(*40*). After visual inputs are integrated into meaningful percepts, fronto-parietal networks mediate downstream encoding processes through information integration, working memory, and attention regulation(*50*, *51*). An intriguing extension of this framework concerns REM sleep, a brain state characterized by rapid eye movements and involved in memory consolidation(*52*). It is possible that saccade-related neural dynamics may contribute not only to wakeful visual encoding but also to sleep-dependent memory consolidation.

Conceptually, our results invite a novel model of visual cognition: instead of being a feedforward transformation of retinal input through the visual hierarchy, visual cognition involves neural events bounded by saccade dynamics that constitute a coordinated, whole-brain mechanism. This model aligns well with the view that human vision is an active sensing process(*53*). We refer to these neural events as SREPs because, although our results show temporal consistency of LFP waveforms around saccades, they are correlational in nature. Without causal manipulation of saccades, it remains an open question whether saccades drive these neural dynamics or whether an underlying internal state coordinates both. Due to clinical constraints, our dataset has limited sampling of the primary visual cortex, which limits our ability to determine whether a latency gradient exists along the visual processing hierarchy.

### Implications for studying active visual memory encoding in humans

Our study has three primary implications for neuroscientific studies of active visual encoding in humans. First, we encourage researchers to adopt active-viewing paradigms and incorporate eye-tracking, as these approaches capture the dynamics of human visual processing in a more ecologically valid manner(*6*). Second, intracranial recordings in neurosurgical patients, such as those undergoing epilepsy monitoring, can reliably capture saccade-related neural dynamics that are free of oculomuscular contamination. Third, because saccades coincide with multiple processes, including motor commands, shifts in retinal input, and cognitive processes, careful analytical dissociation is needed to attribute neural signals to behavioral and cognitive relevance. Our study showcases two key analytical strategies to address this challenge. We examined neural activity during saccades while subjects attended to a blank screen to control retinal input. This analysis reveals that these SREPs are largely non-visually mediated, consistent with previous studies in non-human primates and cats(*9*, *54*). Additionally, we separately correlated SREP characteristics extracted from LFPs and EOG with saccadic behavior measures, such as saccade direction, thereby ruling out potential confounding from oculomuscular activity.

### Proposed mechanisms underlying SREPs and their relationship with successful visual encoding

Katz et al. initially reported saccade-related inhibition of single units in mnemonic structures and interpreted it as a corollary discharge-like signal(*8*). Our results reveal more widespread saccadic modulation and provide direct evidence that SREPs predict visual encoding. Moreover, cortical SREPs did not systematically encode saccade direction. Thus, SREPs are more consistent with the active sensing framework and the study by Leszczynski et al.(*21*), which showed that saccades modulate brain excitability during auditory processing. The existing literature and our results suggest two mechanistically distinct forms of saccadic modulation: (1) an early corollary discharge, and (2) a later modulation of neural activity to support visual encoding(*36*, *55*, *56*).

Within the active sensing framework, a theory proposed that saccades modulate sensory processing by resetting ongoing oscillations to a high-excitability phase(*3*). Our results suggest an alternative, but not mutually exclusive, mechanism of SREPs. We observed SREPs with positive and negative polarities at the same recording sites, with waveforms depending on the ongoing oscillation phase. As these SREPs were derived from locally re-referenced field potentials, they reflected the mesoscale current flows driven by synchronized synaptic currents(*57*). Based on this, we propose a model in which each brain region follows its intrinsic rhythm, with saccades acting as events that synchronize neural activity in a manner contingent on the local rhythm. The consistent LFP waveforms time-locked to saccades may result from the temporal coordination between saccade and intrinsic brain rhythms. This interpretation is supported by observations of phase clustering before saccades in this study (**Figure 3**) and in other studies(*2*, *58*). This model predicts higher intertrial coherence and elevated oscillatory power at saccade times, which are also validated in our study and previous work(*3*, *21*). Given that SREP is a widespread, possibly brain-wide phenomenon, a global, uniform phase reset would likely interfere with ongoing neural computations, such as information coding(*59*). Thus, a phase-contingent synchronization mechanism for processing incoming input may be functionally critical.

Not all visual inputs are remembered equally(*60*). Our study shows that SREP characteristics and ongoing oscillations help explain this variability. Successful encoding was associated with greater SREPs amplitude, particularly in frontal and parietal regions (**Figure 6I**). It is our interpretation that stronger SREPs represent greater attention and may signal priority for encoding in memory circuits(*61*). In addition, successful encoding is associated with earlier SREPs in general, which mirrors Staudigl et al.’s finding that neuronal responses in the medial temporal lobe associated with remembered images occurred earlier than those associated with later forgotten images(*40*). These findings suggest that earlier saccadic modulation is beneficial for visual encoding, although the exact mechanism remains to be elucidated.

It has been reported that greater oscillatory power in the hippocampus was associated with better recognition memory in non-human primates(*3*). Our results provide a comparatively widespread mapping of oscillatory activity to successful visual encoding. The normalized oscillation power at 0.5 OCs before SREP onset latency showed hemispheric asymmetry. In general, higher and lower preceding power in the right and left hemispheres, respectively, were associated with successful visual image encoding. Future studies should examine the hemispheric dominance of memory for auditory, verbal, and/or language-related information not addressed in our task. If SREPs represent synchronization, contingent on local rhythm, then regions in which higher preceding oscillation power predicts successful encoding likely belong to the encoding-relevant network. In other words, their ongoing rhythmic activity is functionally aligned with visual encoding. We note that the functional implications of oscillation are frequency-and region-dependent. For example, increased posterior alpha oscillations in the visual cortex have been linked to an enhanced signal-to-noise ratio via selective inhibition of background activity(*45*), whereas increased hippocampal theta oscillations are associated with active information encoding(*62*, *63*). Given that the dominant frequency across cerebrum resides in the theta–alpha range (5–12 Hz; **Fig. S8D**), it is plausible that both inhibitory gating and encoding mechanisms are concurrent.

### Limitations

Our study establishes SREPs as saccadic modulation of large-scale neural activities that predict human visual memory encoding. Whether visual input is encoded into long-term memory is largely governed by stimulus salience, top-down attention, amygdala-mediated emotional regulation, and hippocampal-dependent consolidation(*64–67*). Our results add saccadic modulation of neural activity as a new dimension to current frameworks of visual memory. We speculate that SREPs reflect a saccade-coupled form of top-down modulation that supports visual memory encoding, though its precise relationship to canonical top-down attentional processes remains to be determined. In this sense, the mind’s eye depends not only on what is seen but also on the neural dynamics that accompany each saccade. Causal manipulations, such as closed-loop brain stimulation timed to saccades to disrupt or augment SREPs within focal or distributed networks, will be critical to confirm this hypothesis. Regardless of how SREPs map onto existing cognitive frameworks, such as top-down attention, the encoding-conducive SREP profile identified in our study, characterized by earlier onset latency and greater peak amplitude, provides a measurable biomarker and potential target for memory-enhancing neuromodulation. However, several caveats need to be kept in mind when interpreting our findings: (1) Our electrode sampling was guided by clinical needs and lacked coverage of the primary visual cortex and brainstem, restricting our ability to localize the anatomical origin of SREPs; (2) Although the memory-coding task permitted free viewing, it did not fully recapitulate naturalistic vision, as participants viewed stimuli on a screen with limited demands for head movements; (3) the biological computations at the cellular level by which SREPs support memory remain to be elucidated; and (4) the present findings were obtained during visual memory encoding in neurosurgical patients and may not generalize to other sensory modalities or healthy populations. Despite these caveats, our findings extend the active sensing framework to human visual memory encoding: each shift in eye gaze is accompanied by a structured neural event that influences whether the encountered visual world is remembered.

## MATERIALS AND METHODS

### Study participant

Eight epilepsy patients (two males and six females) undergoing intracranial monitoring with stereoelectroencephalography (SEEG) at Washington University School of Medicine participated in this study. All participants provided written informed consent in accordance with protocols approved by the Institutional Review Board at Washington University in St. Louis (IRB protocol number: 202104033-1001). Patients were eligible for this study if they were diagnosed with epilepsy and scheduled to undergo intracranial video monitoring for seizure onset localization. None of the included patients reported subjective cognitive impairment, and no abnormalities in eye movements were noted during clinical evaluation. Patient demographics and seizure diagnosis are described in **Table S2**.

### Study design

This study is part of a broader protocol (registered clinical trial identifier: NCT05065450) investigating how amygdala stimulation affects memory formation. To assess recognition memory, subjects were asked to perform visual encoding tasks in which they viewed a series of images of everyday objects or scenes from a standard image set(*66*). Scene images were taken from the Places dataset(*68*) and object images were taken from the Stark Lab’s Mnemonic Similarity Task image-set(*69*). Memory for these images was assessed approximately 24 hours later. Patients could elect to complete multiple iterations of the study protocol across different days, depending on their availability and length of stay in the EMU. Three subjects completed two visual memory encoding tasks on two different days, one subject completed three tasks on three different days, and each of the other four subjects completed one task. Each study iteration followed a two-day structure, comprising a visual encoding task on Day 1 and a recognition memory test on Day 2. The encoding task on Day 1 was administered in two or four sessions per participant to mitigate fatigue and accommodate clinical scheduling. The encoding task comprised 180 trials in total, with each trial defined as the presentation of a single image. The task structure remained constant across iterations. Each study instance used a unique set of images. The details of the behavioral paradigm are provided below.

In less than half of the trials, one second of electrical stimulation was delivered to the amygdala following presentation of each image, followed by a 5-s intertrial interval. To study the saccadic modulation in the normative visual encoding process, we restricted analyses to trials that met the following criteria: (1) no amygdala stimulation was delivered during the trial; (2) the subject maintained their gaze on the monitor for more than half of the trial duration, as determined from eye-tracking data; and (3) the subject pressed one of the response buttons.

Although object and scene processing might engage dissociable neural mechanisms, our primary aim was to characterize neural dynamics time-locked to saccades; therefore, we pooled object and scene trials. Categories were balanced across the set, and exploratory checks did not indicate category-dependent differences in the saccade metrics. A detailed description of the data set, including the total number of study iterations per participant and the total number of encoding trials, is summarized in **Table S1**.

### Experimental setup and data acquisition

We used a desktop computer (ASUS ROG STRIX B550-E GAMING motherboard; AMD Ryzen 9 5900X 12-core processor; 32 GB DDR4 RAM; NVIDIA GeForce RTX 3070 GPU) running the BCI2000 platform to manage stimulus presentation, behavioral logging (e.g., keyboard responses), and data synchronization(*70*). We recorded and synchronized photodiode signals, eye gaze coordinates, and electrophysiological signals. A 27-inch monitor (TUF GAMING VG27AQL1A, ASUS) with a resolution of 2560 × 1440 pixels and a 170 Hz refresh rate was connected to the desktop computer and mounted to a mobile cart using an adjustable monitor arm, allowing flexible positioning to accommodate each patient’s posture and bed orientation.

A screen-based eye tracker (Tobii Pro Fusion, Tobii AB, Sweden) was mounted at the bottom edge of the monitor and recorded gaze position at 120 Hz. We performed eye-tracker calibration at the start of each task using *Tobii Pro Eye Tracker Manager* software, and repeated as needed if the relative position between the participant’s eyes and the eye tracker changed.

We recorded iEEG data using a clinical SEEG system (Neurofax EEG-1200, Nihon Kohden, Irvine, CA) with a JE-120 amplifier (256 channels). Signals from depth electrode contacts were sampled at 2,000 Hz with 16-bit resolution (±3.2 mV input range, 97.65 nV/LSB). Per standard clinical practice, two subgaleal electrodes served as the reference and ground during signal acquisition. Simultaneous scalp EEG was recorded as part of the clinical montage. We specifically used frontopolar electrodes (Fp1, Fp2) to detect oculomotor artifacts.

To capture accurate timing of visual stimulus onset, a photodiode (g.tec medical engineering GmbH, Austria) was affixed to the bottom-left corner of the screen. At the onset of image presentation, a white dot was shown in the bottom-left corner of the screen, aligned with the photodiode. The white dot disappeared at the onset of the ISI. The photodiode output was routed through a multimodal trigger conditioner box (g.TRIGbox, g.tec medical engineering GmbH, Austria) and recorded simultaneously with electrophysiological and eye-tracking recordings.

### Anatomical localization of the electrode

We localized electrodes based on post-operative CT scans co-registered to pre-operative T1-weighted MRI scans (0.5 mm isotropic resolution). We provided details of localization and validation in the **Supplementary Materials**.

To determine the brain regions associated with each electrode, we performed anatomical brain segmentation using FreeSurfer (v7.4.1), which also creates a cortical and subcortical functional parcellation based on the Desikan-Killiany atlas(*71*). We assigned functional-anatomical labels to each electrode contact using a majority-vote volumetric method, in which each electrode was assigned to the brain structure encompassing the majority of segmented voxels within a 10 mm Euclidean radius from the contact center. We chose a 10 mm Euclidean radius to approximate the spatial reach of local field potential recordings. To assess the consistency of the functional–anatomical labeling of electrode contacts, we performed two complementary analyses. First, we used a surface-based inclusion method, in which electrodes were labeled according to whether their coordinates were enclosed by the triangulated surface mesh of a segmented structure. Second, we implemented a probabilistic labeling approach, in which each structure was assigned a probability proportional to the number of its voxels within a 10 mm sphere centered on the electrode.

For group-level analysis and visualization, we mapped individual electrode locations to a common anatomical space defined by the native MRI of a reference subject. To achieve this, we used the ANTs (Advanced Normalization Tools) image registration framework to coregister each subject’s preoperative T1-weighted MRI to the reference MRI. We applied only the affine component of the transformation to the electrode coordinates to preserve their linear relationships. The affine matrices were estimated within the ANTs’ non-linear optimization pipeline to account for anatomical variability.

To characterize regional differences in electrode sampling, we calculated the weighted count of electrode contacts per brain region (**Figure 1D**). In other words, when electrode contacts (∼2 mm³ volume) overlapped multiple brain regions, their contributions were proportionally allocated among these regions. We further quantified sampling density per region by dividing the weighted contact count within each brain region by the volume of that region (**Fig. S2**).

### Assessing memory outcome

We defined two memory outcome measures in our study: 1) Recall rate as the percentage of remembered visual encoding trials over the tested visual encoding trials; and 2) Overall correct recognition rate as the sum of true positives (old stimuli correctly recalled) and true negatives (new stimuli correctly rejected) divided by the total number of test trials. To assess whether recognition memory performance exceeded chance, our primary analysis considered the expected accuracy under unbiased guessing, in which subjects respond “old” or “new” with equal probability on each trial. Under this assumption, the expected hit rate and correct rejection rate are both 50%, yielding an overall chance-level accuracy of 50%.

In cases where the recognition test included an unequal number of previously presented and novel images, we additionally evaluated two response-bias extreme strategies to characterize the range of overall recognition accuracies driven by response bias. In one extreme, a subject could respond “old” on every test trial. Under this strategy, all previously presented images would be counted as correctly recognized, whereas all novel images would be incorrectly classified as old. In our study, 5 subjects completed task iterations that included 160 previously presented images and 80 novel images. This strategy would yield a 100% correct recognition rate for previously presented images, 0% for novel images, and an overall 66.7% correct recognition rate. Based on post hoc inspection of behavioral performance (**Table S3**), one subject (Subject ID 1) exhibited a response pattern consistent with this extreme strategy during one task iteration (Task ID 2) out of the two completed tasks.

In the opposite extreme, a subject could respond “new” on every test trial. Under this strategy, all novel images would be correctly rejected, whereas all previously presented images would be incorrectly classified as new. For the task structure that includes 160 previously presented images and 80 novel images, this strategy would yield a correct recognition rate of 0% for previously presented images, 100% for novel images, and an overall rate of 33.3%. The post hoc inspection of behavioral performance indicated that no subjects exhibited response patterns consistent with this strategy.

Given that all subjects verbally reported completing the task as instructed, and that eye-tracking showed normative visual scans during the encoding phase of the task, we did not exclude data based on the post hoc inspection of behavioral performance.

### Intracranial EEG preprocessing

To ensure the analysis focused on physiologically meaningful, non-pathological signals, we systematically identified and excluded electrodes exhibiting epileptiform discharges or artifactual activity (e.g., due to poor contact). To identify electrodes with poor contact, we computed the root-mean-square (RMS) amplitude of the 60 Hz line-noise component using a narrowband infinite impulse response (IIR) filter using *iirpeak* in MATLAB with a peak frequency of 60 Hz and a bandwidth of 0.1 Hz as inputs. Electrode contacts with 60 Hz RMS amplitude deviating more than two standard deviations (SDs) from the median were flagged and excluded. To detect potential shorted (i.e., electrically bridged) contacts, we computed pairwise Pearson correlation coefficients between all contacts within each electrode trajectory after re-referencing. For re-referencing, we applied common average referencing for each electrode trajectory after excluding electrode contacts with excessive 60 Hz line noise. We identified contacts exhibiting excessively high correlation with non-adjacent neighbors (*R*^2^ > 0.999) as potentially electrically shorted and excluded them from further analysis. We did not find any shorted contacts.

To identify electrodes with epileptiform activity (e.g., sharps and spikes), we identified time periods in the signal that exhibited exceptionally sharp transients(*72*). For this purpose, we first extracted high-frequency iEEG signals (>250 Hz) using a 4^th^-order Butterworth filter (*butter* function in MATLAB). We computed the analytic envelope of the high-pass-filtered signal using the Hilbert transform (*hilbert* function in MATLAB) and identified time points at which the envelope exceeded 5 SDs above the mean. Additionally, we identified time points at which both the raw signal amplitude and its temporal gradient exceeded 5 SDs above their means. We marked the identified time points as epileptic and calculated the proportion of epileptic time per electrode. We excluded electrodes exhibiting epileptiform activity for more than 1% of the recording duration.

We applied the procedures for identifying line noise and shunted contacts to microwire recordings. We did not apply the epileptiform activity exclusion criterion to microwires, as they recorded both neuronal spiking activity and LFPs. **Table S3** summarizes per-subject counts of retained electrodes for subsequent signal processing.

To enhance spatial specificity and remove common artifactual noises, we re-referenced intracranial signals using a small Laplacian re-referencing, separately for macro electrode and micro wires. We calculated inter-contact Euclidean distances derived from the three-dimensional electrode coordinates. For each contact, neighboring electrodes were identified as those located within a predefined spatial threshold of 6 mm. We then epoched the resulting Laplacian re-referenced signals relative to stimulus onset for subsequent SREP analyses. To ensure the robustness of downstream analyses to the referencing scheme, we tracked the number of neighboring electrodes. For some electrodes located at the ends of a shank, where only a single adjacent contact was available, the small Laplacian re-referencing became bipolar referencing.

### Saccade detection and characterization

The recorded eye-tracking data included three-dimensional eye position and two-dimensional gaze direction, expressed relative to the top-left corner of the display monitor. Gaze coordinates were defined on the screen plane (Z = 0), whereas the Z-coordinate of the eye position represented the eye’s distance from the display along the axis orthogonal to the screen. To ensure that saccade detection was based on eye gaze sample where no eye blinks or off-screen gaze occurred, we preprocessed the eye-tracking data separately for each eye and marked eye tracking samples as invalid if they met two criteria: (1) the eye tracker reported loss of tracking (e.g., during blinks or partial eyelid closures), or (2) the gaze coordinates were outside the physical screen bounds. To account for signal artifacts at the edges of blinks or tracking dropouts, we extended each invalid segment by ±30 ms on both sides.

To account for the possibility that the eye tracker only captured one of the two eyes, we computed the proportion of valid samples for each eye and selected the eye with the higher proportion of valid data for further analysis. We time-locked the gaze data to the image presentation. We defined each trial as the interval from 2 s before the onset of the fixation cross presentation to 2 s after image presentation offset (**Figure 1B**). To ensure that our analyses reflected periods during which subjects were encoding visual information, we excluded individual trials with more than 50% invalid eye-tracking data.

To identify saccade onsets, we segmented the continuous gaze traces into periods of fixation and saccades using the *ClusterFix* algorithm(*73*). In brief, the algorithm identified eye movement events by clustering consecutive gaze coordinate samples based on distance, velocity, acceleration, and angular velocity to classify them as fixations or saccades. We linearly interpolated invalid samples in gaze data to maintain the temporal continuity required by the clustering algorithm. After clustering, we verified that both onset and offset for each identified saccade and fixation occurred during valid (e.g., noninterpolated) samples. Saccadic or fixation events overlapping interpolated segments were flagged as invalid and excluded from downstream analyses. This conservative approach ensured that only fixations and saccades derived from true gaze samples were included in the analysis. We validated that the velocity and duration of detected saccades were within the expected range of human eye-movement parameters (**Fig. S1**).

### Identification of saccade-related evoked potentials (SREPs)

To identify SREPs, we analyzed re-referenced LFP signals time-locked to saccades. The dataset was organized hierarchically: each subject completed one or more encoding tasks, each task consisted of multiple recording sessions, and each session contained LFP signals from multiple electrode contacts. Within each session, there were multiple trials (e.g., image presentation), during which one or more saccades occurred. For each encoding task completed by a subject, we segmented the re-referenced signals into epochs relative to saccade onsets and combined all epochs across trials and sessions within that task. This study focused on saccades occurring during the encoding of the presented image. To isolate SREPs from stimulus-driven responses, we excluded saccades and associated SREPs occurring 200 ms before or after the image onset. For control analyses, we separately analyzed saccades made during fixation-cross presentation, when the screen background was uniformly black with a central white cross occupying approximately 2% of the screen’s horizontal dimension. The cross-presentation period provided a baseline condition for evaluating whether peri-saccadic EPs reflected visual transients. For each epoch, we defined the pre-saccadic interval from -200 to -10 ms relative to the saccade onset as the baseline period. We normalized each epoch by z-scoring its LFP signal based on the baseline mean and SD. To obtain the mean SREP waveform for each electrode contact, we averaged the normalized LFP signals across all saccades, trials, and sessions.

To determine whether SREPs were consistent across epochs, we calculated the pairwise Pearson correlation coefficients among normalized epochs within the peri-saccadic window (-10 to +40 ms relative to saccade onset) and, for comparison, within the baseline window (-200 to -10 ms relative to saccade onset)(*28*). These windows were defined to capture the immediate neural dynamic following a saccade while avoiding temporal overlap between EPs arising from consecutive saccades. To ensure robustness of subsequent analyses, we systematically varied the baseline window length and confirmed that results were consistent (**Fig. S6**).

The distribution of correlation coefficients during the peri-saccadic period exhibited a bimodal pattern, suggesting SREPs of two polarities (**Figure 2C**). To statistically evaluate this observation, we quantified the extent to which the peri-saccadic correlation distribution deviated from the unimodal baseline distribution. We quantified bimodality based on the separation between the inter-trial correlation distributions during the peri-saccadic and baseline periods. We computed the intersection point of their cumulative distributions and measured the absolute difference in area under the curve (AUC) between them as their separation effect size (**Fig. S4**). This separation effect size reflected the extent to which inter-trial correlations during the peri-saccadic period were bimodal, with larger values indicating stronger bimodality. To assess statistical significance, we applied Kolmogorov–Smirnov tests to determine the separation effect size threshold corresponding to the significance level adjusted for multiple comparisons.

This analysis confirmed the presence of bimodal SREPs. To identify the two types of SREPs, we applied hierarchical clustering to the inter-trial correlation matrix using the MATLAB *linkage* and *cluster* functions with *maxclust* input set to 2. Each element of this matrix represented the correlation coefficient between a pair of epochs. In brief, hierarchical clustering groups epochs based on pairwise correlations and iteratively merges those with the most similar waveforms to form a dendrogram that captures the hierarchical relationships among epochs. To ensure the robustness of the hierarchical clustering algorithm, we required a minimum cluster size of 25% of the total number of epochs. Failure to meet this criterion could occur when the inter-trial correlation structure was weakly bimodal. In this study, only three electrode contacts had peri-saccadic correlation matrices that failed to meet this criterion. For these electrode contacts, we implemented an adaptive re-clustering procedure that iteratively removed epochs with globally weak correlations, defined as the lowest mean absolute correlation across epochs, and repeated the clustering until both clusters satisfied the size criterion.

After identifying SREPs of two polarities, we quantified the separability between peri-saccadic and baseline correlation distributions for each cluster. To do so, we computed the receiver operating characteristic (ROC) curve, which characterizes how well the two distributions can be distinguished. The ROC curve plots the true positive rate (TPR), defined as the fraction of peri-saccadic correlations exceeding a given threshold, against the false positive rate (FPR), defined as the fraction of baseline correlations exceeding the same threshold. Thresholds were sampled uniformly between −1 and 1. The area under the ROC curve (AUC) measures the separability. Larger AUC values indicate greater divergence of peri-saccadic correlation structure from baseline, reflecting more consistent SREP waveforms.

As a control, we repeated the clustering and separability quantification procedures described above after permutating the peri-saccadic and baseline correlation matrices. We applied the identical adaptive hierarchical clustering procedure on the baseline correlation matrix. We then recomputed the ROC curve following the same procedure, but with the roles of peri-saccadic and baseline correlations reversed. In other words, the TPR now represented the fraction of baseline correlations exceeding a given threshold, and the FPR represented the fraction of peri-saccadic correlations exceeding the same threshold. We then compared the observed and control AUC values to identify electrode contacts with consistent SREPs, using AUC difference as a measure of consistency. This control analysis accounts for potential bias introduced by the clustering algorithm and enables estimation of separability specifically attributed to peri-saccadic LFP waveform similarity.

We computed the SREP consistency during fixation-cross presentation periods using the same method described above.

### SREPs prevalence

To investigate whether SREPs were localized to specific anatomical circuits or broadly distributed throughout the sampled cortex, we defined SREP prevalence at anatomical regions as the proportion of electrode contacts with an area under the ROC curve in the peri-saccade condition above the mean + 2 SD of the control area under the ROC curve.

### Characterization of SREP waveforms

To test whether the observed SREPs represented oculomuscular signals or neural mechanisms of visual encoding, we quantified their waveform characteristics, including latency, peak amplitude, peak time, trough time, and peak-to-trough amplitude. We used these SREP characteristics in subsequent analyses of the correlation between SREP peak amplitude and saccade eccentricity, and in machine learning models to map this relationship to successful visual encoding. We calculated SREP characteristics at both averaged and single-trial levels. Here, averaged SREPs refer to SREPs across all saccades, trials, and sessions; and single-trial SREPs refer to the LFP trace following a single saccade.

For each electrode contact and cluster identified by the hierarchical clustering procedure described above, we computed the average SREP and quantified its characteristics. We normalized SREP amplitude using a z-score based on the baseline. We used MATLAB’s *findchangepts* function on the first derivative of the averaged SREPs to identify two change points. This function partitions the time series into segments whose means differ maximally, thus detecting points of abrupt change in slope. We defined the first change point SREP latency, corresponding to the earliest statistical deviation from baseline (**Figure 2H**). Within a ±10 ms window around the second change point, we identified the peak time as the point of maximal normalized amplitude for positive-polarity clusters, or minimal amplitude for negative-polarity clusters. We extracted the peak amplitude and peak-to-trough amplitude based on the averaged SREP. We defined trough time as the time point within a 10-ms window around latency at which the normalized amplitude was minimal for positive clusters or maximal for negative clusters.

For each single-trial SREP, we centered the ±10 ms latency search window on the latency of the averaged SREP from its assigned cluster. We applied MATLAB’s *findchangepts* function to the first derivative of the single-trial LFP to detect one change point. We defined this inflection point as the single-trial SREP latency. Similarly, we defined peak-amplitude search windows centered on the averaged SREP peak time from the associated cluster. We identified single-trial SREP peak amplitude as the maximum (for positive clusters) or minimum (for negative clusters) amplitude within this window. We identified the single-trial SREP trough between latency and the peak time, defined as the minimal (for positive clusters) or maximal (for negative clusters) amplitude in this interval. We flagged a trial as having an invalid peak–trough identification if the detected trough amplitude exceeded the detected peak amplitude for positive clusters or the detected peak amplitude exceeded the detected trough amplitude for negative clusters. This happened for 1% of the electrode contacts. To verify that the hierarchical clustering procedure reliably separated SREP waveforms of two polarities, we quantified the similarity between each single-trial SREP and its cluster-level averaged waveform using Spearman’s correlation coefficient. For subsequent analysis, unless specified otherwise, we included single-trial SREPs whose waveforms were similar to the averaged SREP, defined as those with a Pearson correlation coefficient above the median. We provide further details on quantifying intra- vs inter-contact variability of SREP characteristics and on visualizing SREP characteristics in the **Supplementary Materials**.

As waveform characterization is less reliable for single-trial SREPs that deviate substantially from the cluster-level averaged SREP due to background neural activity, we quantified the alignment between each single-trial SREP and the corresponding cluster-averaged SREP waveform using Spearman’s correlation, and used single-trial SREPs with similarity above the median (i.e., 0.413) for analysis based on single-trial SREPs. The distribution of waveform alignment was skewed towards positive values, with an area under the kernel density curve of 0.812 for correlations greater than zero (**Fig. S7A–B**). This indicates a general alignment between single-trial and averaged SREP waveforms.

### Assessing the relationship between SREP characteristics and saccadic properties

To test whether the observed SREPs reflected oculomuscular activity, we evaluated three predictions derived from an ocular artifact model. The first prediction is that cluster-level SREP amplitude, including peak and peak-to-trough amplitude, should vary systematically with eyeball-to-electrode distance. To test this prediction, we performed linear regression on cluster-level averaged SREP amplitudes with eyeball-to-electrode distance as the independent variable, using the Python library *statsmodels* (**Figure 2I**). We computed the explained variance, that is, the coefficient of determination (R^2^). As a control, we extracted electrooculogram (EOG) signals derived from the differential potential between fronto-polar scalp EEG electrodes (Fp1 and Fp2), and applied the same epoching, clustering, and waveform characterization procedures to EOG signals as for intracranial EPs. We computed R^2^ using the same method. We then compared R^2^ between intracranial electrode contacts and EOG signals using statistical tests described in the **Quantification and Statistical Analysis** section.

The second prediction of the ocular artifact model is that single-trial SREP amplitude should covary with saccade kinematics such as saccade eccentricity. To test this, we performed linear regressions between single-trial SREP amplitude, including peak amplitude and peak-to-trough amplitude, and saccade eccentricity. Data was omitted when we failed to detect trough. We conducted separate regressions on intracranial LFPs and EOG signals, treating eccentricity as the independent variable. As complementary analyses, we calculated the Pearson’s correlation coefficient between single-trial SREP amplitude and saccade duration.

The third prediction of the ocular artifact model is that SREP polarity reflects the directional movement of extraocular muscles; therefore, it should be associated with saccade direction. We examined the relationship between saccade direction and the polarity of EP at saccade times derived from EOGs and the relationship between saccade direction and the polarity of SREPs from intracranial signals. We first quantified directional clustering of saccades using the Rayleigh statistic. For each electrode contact, we extracted the saccade direction associated with each polarity and measured the degree of unimodal bias of the circular distribution of directions using Rayleigh statistics (Monopolar condition). We compared them against Rayleigh statistics computed from the same number of saccades pooled across both polarities (Combined condition), thereby controlling for sample size.

To avoid the potential confounds of sample size on the calculation of Rayleigh statistics, we resampled the same number of saccade directions for positive, negative, and combined polarities. To avoid unstable estimates due to sparse saccades associated with a single polarity, we imposed a minimum-saccade criterion based on a global quantile threshold (q = 0.2). This yields sample sizes for positive and negative EPs at saccade times from EOG (*N_Positive_* = 15, *N_Negative_* = 18, *N_Combined_* = 21). We tested for differences in the circular distributions of saccade directions between opposing polarities for SREPs derived from intracranial recordings and EOG-derived EPs at saccade times (see **Quantification and Statistical Analysis** section). We performed the analysis in two ways: (1) treating saccades during image presentation and fixation-cross presentation as separate samples, and (2) pooling saccades across both task epochs. The results were consistent across both analyses.

### Oscillation detection and quantification

To test if pre-saccadic oscillatory activity influenced SREP polarity, we first identified the dominant oscillation frequency for each electrode contact. We computed the power spectral density (PSD) using Welch’s method on continuous re-referenced LFPs. Specifically, we used MATLAB’s *pwelch* function using 1-s Hanning windows (2,048 samples) with 50% overlap and a 2-s discrete Fourier transform length (4096 samples). To separate periodic oscillatory components from the aperiodic activity, we fitted a 1/f model to the PSD in log–log space using MATLAB’s *polyfit* function. To minimize the influence of 60 Hz line noise, we fitted a linear model to the log-transformed PSD curve within the frequency range of 1-58 Hz. We calculated PSD residues as the pointwise difference between the original PSD curve and the fitted line. To ensure that oscillatory peaks did not bias the aperiodic fit, we excluded frequencies with positive residuals exceeding the median residual, and then recalculated the 1/f fit. We defined the dominant oscillation frequency for each contact as the frequency at which the residual PSD curve reached its maximum. We quantified the power of these oscillatory activities as the deviation between the power spectral density and the 1/*f* fit at the location-specific dominant frequency.

To quantify the instantaneous phase and power of the oscillation at the dominant frequency, we applied the Hilbert transform to narrowband-filtered signal segments surrounding each saccade. To ensure that estimates of pre-saccadic oscillatory activity were not affected by the presence of SREP, we segmented LFPs from -3 s before saccade onset up to SREP latency. For each electrode contact and each encoding task, we estimated the instantaneous oscillation phase and power separately for two clusters of SREPs. To minimize edge artifacts potentially introduced by filtered signals and the Hilbert transform, we symmetrically padded both ends of each segment with 500 ms of data before filtering. We applied a second-order Butterworth band-pass filter centered on the identified dominant oscillation frequency with a half-bandwidth of 3 Hz using MATLAB’s butter function. To implement zero-phase digital filtering, we used MATLAB’s *filtfilt* function to apply the filter in both the forward and reverse directions. We then used MATLAB’s Hilbert function to obtain the analytic signal. To ensure unbiased estimates, we only derived the instantaneous power and phase from the portions of the analytic signal corresponding to the unpadded original LFPs. Because the oscillation cycle (OC) duration varied with the dominant oscillation frequency, we defined pre-saccadic time points relative to each electrode contact’s OC. Our time points of interest included 0.5, 1, 1.5, 2, and 3 OC before the latency.

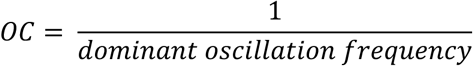

To study the interplay between SREP and oscillatory activities, we estimated instantaneous oscillation power and phase after SREP latency. For this purpose, we segmented LFPs from -300 to 300 ms relative to the saccade onset and repeated the padding, filtering, and Hilbert-transform procedures. To enable subsequent cross-session and cross-subject comparisons, we z-scored instantaneous power relative to a baseline window, defined as -200 ms to -90 ms relative to saccade onset.

### Assessing the relationship between pre-saccadic oscillation phase and SREP polarity

To test whether the oscillatory phase preceding saccades differed between SREP polarities, we first wrapped phase values from [–π, π] to [–2π, 2π] to preserve periodicity and continuity. We then estimated the probability density functions separately for each polarity. For each of the five time points: 0.5, 1, 1.5, 2, and 3 OCs before the onset of each SREP, we quantified phase separation as the area under the absolute difference between the two probability density functions of the instantaneous phase associated with each polarity. To determine statistical significance, we generated null distributions by randomly permuting SREP polarity labels (50 iterations per electrode contact) and recalculated the separation metric for each permutation. Observed and null values were compared using two-sided Wilcoxon signed-rank tests (N = 1,970 electrode contact pairs per condition). Effect sizes were quantified using the rank-biserial correlation, which increased progressively from 0.204 to 1.000 across the five pre-saccadic time points, indicating a gradual divergence of oscillatory phase between SREP polarities as saccades approached.

To evaluate whether the pre-saccadic oscillation phase influenced the polarity of SREPs, we quantified phase separation between positive and negative SREP clusters at both the single-contact and contact-averaged levels. Positive and negative SREP clusters refer to sets of SREPs with positive and negative waveform polarities. Analysis in this section used the instantaneous phase derived from LFPs truncated before the SREP latency (see **Oscillation detection and quantification**). Because the phase is circular, direct density estimation on [–π, π] can introduce boundary artifacts, as probability mass at the endpoints (–π and +π) is artificially split. To avoid this, we extended the phase range by concatenating copies of the phase values shifted by ±2π, preserving continuity across the circular boundary ±π. At the single-contact level, we computed phase histograms for positive and negative polarities using 72 equally spaced bins spanning [–2π, 2π] using the *histogram* function from the *Numpy* Python package. To obtain a continuous phase probability density function (PDF), we applied a Gaussian kernel filter (σ = 1 radian) using the *gaussian_filter1d* function from the SciPy Python package. We normalized each estimated phase PDF such that its integral over the canonical [–π, π] range equaled 1, using the *Simpson* function from the Scipy Python package. We calculated phase separation as the area under the absolute difference between the two normalized PDFs using the same *Simpson* function. At the single-contact level, we computed phase separation for each electrode contact across the five pre-saccadic time points of interest.

To determine statistical significance, we generated null distributions by randomly permuting SREP polarity labels and recalculated the phase-separation metric for each permutation. We conducted 50 permutations per electrode contact. We then compared observed and null separation values at the five pre-saccadic time points of interest, as detailed in the **Quantification and Statistical Analysis** section.

### Analysis of oscillatory power and phase synchronization associated with SREPs

To investigate how SREPs interacted with ongoing neural oscillations, we analyzed temporal changes in oscillation power and phase synchronization around saccade times. To quantify inter-trial phase alignment, we computed the inter-trial phase coherence (ITPC) for each polarity and each electrode contact, based on the instantaneous phase of the narrowband analytic signal filtered at each contact’s dominant oscillation frequency, which is detailed in the Oscillation detection and quantification section. ITPC is defined as:

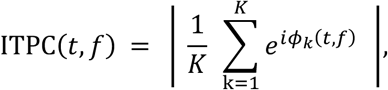

where ***φ_k_***(***t***, ***f***) denotes the instantaneous phase of saccade **k** at time ***t***, and ***f*** denotes the dominant oscillation frequency. For each electrode contact, we quantified ITPC at time points of interest, including 0.5, 1, 1.5, 2, and 3 OCs before the SREP latency, at latency, peak time, and 0.5 OCs after peak time. To understand whether phase synchronization was accompanied by increased oscillation power, we computed Pearson’s correlation between changes in ITPC and changes in instantaneous power relative to 2 OCs before SREP latency at time points of interest, using the *pearsonr* function from the *Scipy* Python library. To enable comparison across electrode contacts and subjects, we normalized the oscillatory power to its magnitude at two OCs before the SREP onset latency.

### Spatial modeling of SREP latency using Gaussian Mixture Models (GMMs)

To examine whether early SREPs were spatially localized, we stratified electrode contacts into percentile bins based on their average SREP onset latency (e.g., 0–15^th^, 30–45^th^, 60–75^th^ percentiles). For each latency percentile bin, we applied 3D GMMs to the spatial coordinates of electrode contacts in the common anatomical space. We implemented GMMs using the *GaussianMixture* function from the *sklearn* Python package, with the number of components (e.g., *n_component*) set to 2 to identify bilateral spatial clusters corresponding to the two hemispheres. We extracted the centroids of identified clusters to determine their hemispheric localization. We quantified the spatial spread as the Euclidean norm of standard deviations along X, Y, and Z axes, estimated by the GMMs. To assess if there is progressive spatial expansion of SREPs across regions, we calculated spatial spread across latency percentile bins and assessed monotonic trends using Spearman’s rank correlation coefficient. A positive trend would suggest early focal localization of SREP and later widespread involvement.

To quantify temporal involvement across anatomical regions, we computed the proportion of electrodes within each region exhibiting SREPs in each latency percentile bin, defined as the number of electrode contacts with SREPs in a given latency range divided by the total number of sampled electrodes in that region. To identify regions showing systematic early or late SREPs, we assessed monotonic change using Spearman’s rank correlation coefficient. To examine whether regions showing strong SREPs also exhibited spatial clustering, we repeated the GMM analysis and proportion–percentile correlation analyses described above for SREP peak amplitude.

### Saccade-Related Neural Dynamics (SRND) model

SREPs and the preceding oscillatory activity jointly reflect transient neural dynamics that may contribute to visual encoding. The finding that the pre-saccadic oscillation phase influenced SREP polarity, combined with the finding that SREPs were accompanied by increase in oscillation power and inter-trial phase coherence, motivated us to develop a model, the SRND model, to parameterize and reconstruct the neural dynamics around saccade onsets (**Figure 5A**). For each electrode contact and saccade, we reconstructed the saccade-related neural dynamic *SRND*(*t*) as a composite of (1) a sigmoidal SREP waveform and (2) its preceding oscillatory activity. We modeled SREP waveforms *EP*(*t*) as sigmoidal rise-and-fall functions parameterized by three temporal features: latency *t_latency_*, peak time *t_peak_*, and peak-to-trough amplitude *Amp* . As SREPs exhibited opposing polarities, we used signed peak-to-trough amplitude to describe their magnitude, capturing both polarity and peak amplitude.

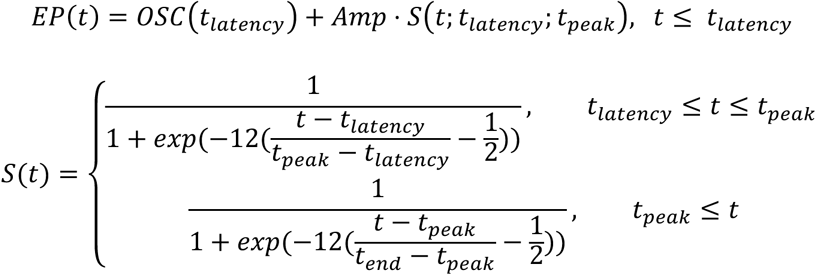

We modeled the preceding oscillatory activity *Osc*(*t*) as a sine wave parameterized by instantaneous power and phase *φ* of oscillation at the dominant frequency at half OCs before SREP latency.

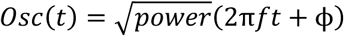

The combined LFP trace *SRND*(*t*) is the linear combination of *EP*(*t*) and *Osc*(*t*). We ensured continuity at SREP latency by adding *Osc*c*t_latency_*d to *EP*(*t*). To truly represent the transient saccadic modulation, we confined the support for time as [-50, 100] ms relative to saccade onset.

### Assessing the relationship between eye movement parameters and encoding performance

To assess whether eye movement parameters were related to visual encoding performance assessed by recognition accuracy on the following day, we compared saccade eccentricity, duration, and direction between remembered and forgotten trials. We quantified the saccade direction dissimilarity as the absolute area between circular kernel density estimates of saccade direction for the two conditions. To evaluate statistical significance, we compared the observed direction dissimilarity with a null distribution generated from 2,000 random permutations of trial labels (remembered vs. forgotten).

### Random forest classifier

To test our primary hypothesis that the SRND model predicts visual encoding performance, we trained random forest (RF) classifiers to learn the mapping between SRND parameters and recognition memory outcomes (remembered vs. forgotten), as illustrated in **Figure 5B**. In brief, an RF consists of an ensemble of decision trees, where each tree predicts the class of an observation based on feature thresholds, that is, each tree partitions the feature space through a sequence of decisions. The final prediction aggregates predicted probabilities across all trees. Each tree is trained on a bootstrap sample of the data. Training a tree involves partitioning the feature space using thresholds (i.e., identifying feature subsets and split thresholds) that maximize the reduction in impurity (Gini index) at each decision node. For a node *i* containing samples from *K* classes, the Gini index is defined as:

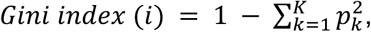

where *p_k_* denotes the proportion of samples that belong to class k. After training, each terminal leaf for each tree stores the empirical class distribution of the training samples that reached it, which defines the predicted probability for each class.

### Training and testing data structure

A sample for the RF classifier corresponded to SRND parameters derived from LFPs at a single saccade event from one electrode. We labeled the sample as “remembered” if the image viewed during that saccade’s trial was later recognized by the patient, or “forgotten” if not. Here, a “trial” denotes one image presentation during the encoding task. The input feature vector comprised electrode spatial coordinates along the right-left (*x*), anterior-posterior (*y*), and superior-inferior axes (*z*), and the five SRND parameters. The electrode coordinates (*x*, *y*, *z*) for all participants were expressed in a common anatomical space, as detailed in the **Anatomical localization of the electrode** section. The five SRND parameters included SREP latency, peak-to-trough amplitude, peak time, and instantaneous power and phase of the preceding oscillation at half OCs before SREP latency.

Because multiple saccades, and thus multiple samples across electrode contacts, could occur within a trial, we grouped samples by trial before splitting the dataset into training and testing sets. This choice ensures that all samples from the same trial are assigned exclusively to either training or testing, preventing the model from learning trial-specific idiosyncrasies, such as stimulus content. Thus, all saccades and their associated SREPs during a given image presentation were confined to either the training or the test set, so the model was never exposed to SRND from the same presented image. Therefore, by splitting data at the trial level, we evaluated whether SRND features alone carry information predictive of recognition memory. If the RF classifier successfully predicts recognition from SRND parameters at a specific location, it provides strong evidence that SRNDs reflect neural mechanisms relevant to visual encoding.

### Training RF classifiers

Given class imbalance, we implemented balanced RF classifiers using the *imbalanced-learn* and *scikit-learn* Python libraries. A balanced RF classifier draws a bootstrap sample from the minority class (e.g., forgotten) and samples with replacement the same number of samples from the majority class (e.g., remembered). To learn the regional mapping between SRND parameters and visual encoding, while avoiding overfitting to subject-specific electrode coverage, we added Gaussian spatial jitter to electrode coordinates in the training data using the *normal* function from the *Numpy.random* Python library. We sampled the random *noise* from a normal distribution with standard deviation proportional to the variance of that coordinate across electrodes.

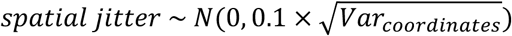

To reliably evaluate model performance, we implemented 5-fold cross-validation using the *StratifiedKFold* function from the *Scikit-learn* Python library. Unless otherwise specified, we used default hyperparameters of 200 decision trees, a maximum depth of 20, and a minimum leaf size of 1. The results were from this setting unless otherwise noted. To verify that the chosen hyperparameters achieved an appropriate model complexity (i.e., neither underfitting nor overfitting), we systematically varied them and reevaluated model performance (**Figure 5E**).

### Model evaluation

Our primary metric for model evaluation was balanced accuracy on held-out test data because the number of remembered and forgotten trials was imbalanced (**Table S3**):

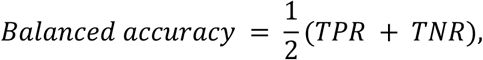

where TPR (true positive rate) is the proportion of correctly classified remembered trials among all remembered trials, and TNR (true negative rate) is the proportion of correctly classified forgotten trials among all forgotten trials. We also report the area under the receiver operating characteristic curve (ROC AUC) and standard accuracy. ROC AUC summarizes the true-positive vs. false-positive trade-off across thresholds and is not affected by prevalence. Operationally, we constructed an ROC curve by plotting the TPR against the FPR (false positive rate) as the classification threshold varied from 0 to 1.

We evaluated model performance at the location-, saccade-, and trial levels. As we observed SREPs across the cerebrum, we reasoned that if the RF model learned local mapping between SRND parameters and visual encoding outcomes, combining predictions across locations within a saccade (e.g., spatial aggregation) should yield a more reliable estimate of the visual encoding outcome than location-level prediction. At the location-level, the model predicted the probability of being remembered and the class of each sample, that is, the set of five SRND parameters at one electrode contact location for a single saccade. For the RF classifier, the predicted probability 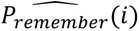 for a sample was computed as the mean of predicted probabilities estimated by all decision trees in the ensemble. Tree *t*’s predicted probability for sample *i*, *p_i_*_,*t*_, equals the proportion of training samples belonging to the “remembered” class in the terminal leaf where that sample falls.

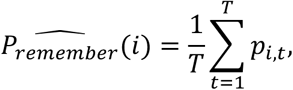

where *T* is the number of trees and *p_i_*_,*t*_ denotes the proportion of “remembered” samples in the terminal leaf reached by sample *i* in tree *t*. For each sample, we determined the predicted class label 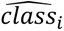 by comparing the predicted probability with 0.5:

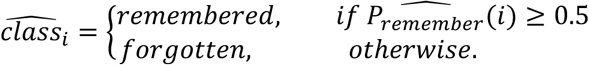

We computed location-level balanced accuracy and ROC AUC directly from these sample-level predictions.

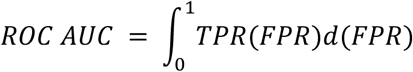

At the saccade level, we averaged the model’s predicted probabilities across all electrode contacts during that saccade to obtain one probability of successful encoding per saccade. Because this saccade-level predicted probability was continuous, classifying the memory outcome (remembered vs. forgotten) associated with each saccade requires a decision threshold.

To evaluate classifier performance in a threshold-independent manner, we calculated the area under the ROC curve, which summarized sensitivity–specificity tradeoffs across all possible decision thresholds. We used the optimal balanced accuracy as secondary performance metric. At the trial level, we aggregated predicted probabilities within a trial to yield a single predicted probability per trial. We then derived the predicted class and computed balanced accuracy and ROC AUC accordingly.

To test whether performance exceeded chance under the null hypothesis of no relationship between SRND features and successful visual encoding, we generated a null distribution by permuting labels. We then retrained and reevaluated the RF models on the permuted data using the same pipeline detailed in **Training and testing data structure**, **Training RF classifiers**, and **Model evaluation sections**. We compared the observed performance metrics, including balanced accuracy and ROC AUC, against their corresponding null distributions derived from permuted data.

To confirm that the trained random forest was neither underfitting nor overfitting, we systematically evaluated the influence of three key hyperparameters on model performance (i.e., testing balanced accuracy): the number of trees, the maximum tree depth, and the minimum leaf size. Leaf nodes are the terminal decision points of each tree, and their size controls how finely the model partitions the data. Leaf nodes are the terminal decision points of each tree, and their size controls how finely the model partitions the data. We reasoned that increasing tree depth and decreasing the minimum leaf size allowed the model to learn more complex relationships. Consistently, classifiers trained on SRND parameters with greater tree depth and smaller leaf size achieved higher training accuracy until saturation (**Figure 5E**). For primary analyses, we used the default hyperparameters of the *imbalanced-learn* Python package (200 trees, maximum depth = 20, minimum leaf size = 1).

To assess whether the model’s probabilistic outputs meaningfully aligned with behavioral outcomes, we compared the average predicted probability between remembered and forgotten trials, as a complementary diagnostic of model validity. In addition, we quantified and compared the proportions of predicted remembered and forgotten saccades across actual visual-encoding outcomes. To assess inter-subject generalizability, we evaluated the model’s performance separately for each subject’s test data. For each subject, we computed the mean saccade- and contact-level balanced accuracy across CVs. To determine whether observed performance exceeded chance, we generated a subject-specific null distribution by retraining RF models on permuted labels using the identical pipeline. We quantified the percentile rank of each subject’s observed performance relative to the null distribution at the individual level (**Fig. S11E**). We provide details for assessing the reproducibility of learned mappings between SRND and visual encoding outcome in the **Supplementary Materials**.

### Logistic regression for linear mapping between SRND parameters and visual encoding outcomes

To determine if a linear relationship could explain the mapping between SRND parameters and visual encoding outcomes, we trained logistic regression models using the same dataset and class labels as those used for training RF models (see **Training RF classifiers** section). Given that the brain’s functional organization is spatially nonlinear, we expected the logistic regression models to perform worse than the RF models. Nonetheless, this analysis served as a control to quantify the extent to which a linear model could capture the relationship between SRND features and visual encoding outcomes.

We implemented logistic regression with a logit link function and L2-norm regularization using the *LogisticRegression* class from the *Scikit-learn* Python package. The training and testing data structure followed the same rationale and procedures described in the **Training and Testing Data Structure** and **Training RF Classifiers** sections. To address class imbalance between samples labeled “remembered” and “forgotten”, we applied the Synthetic Minority Over-Sampling Technique (SMOTE) using the *SMOTE* function from the *imblearn.over_sampling* package. In brief, SMOTE augments the minority class by generating synthetic samples in feature space(*74*). For each minority-class sample, SMOTE generated synthetic examples along the line segments connecting that sample to any of its k minority class nearest neighbors (default k = 5), thereby reducing classifier bias toward the majority class.

As in RF mode training, we introduced spatial jitter to the electrode coordinate features to mitigate overfitting to exact electrode coordinates, and we implemented 5-fold cross-validation (see the **Training RF classifiers** section). After training the logistic regression model, we evaluated its performance using balanced accuracy and ROC AUC on the held-out test set. The quantification of these performance metrics and assessment of their statistical significance (i.e., comparing performance metrics to their permutation-based nulls) are detailed in the **Model evaluation** section. To interpret the learned mapping, we extracted the regression coefficients for each SRND parameter at each electrode contact location. Positive coefficients indicated that higher feature values increased the likelihood of successful encoding, whereas negative coefficients indicated the opposite. Because logistic regression assumes linearity and monotonic effects, caution is warranted when interpreting coefficients for circular variables, that is, the phase of preceding oscillatory activity.

### Shapley additive explanation

To interpret how specific SRND parameters contributed to the random forest (RF) model’s predictions of visual encoding outcomes, we employed the Shapley additive explanation framework, a model-agnostic approach for explaining machine learning models(*34*). The Shapley value is the average marginal contribution of a feature value across all possible coalitions, quantifying how much a given feature value increases or decreases the predicted probability of a machine learning model. In **Supplementary Materials**, we provide the mathematical definitions and properties of Shapley values as formulated initially by Lloyd Shapley.

### Shapley value-informed thresholds and directions

To better interpret the RF model’s mapping between SRND parameters and visual encoding outcomes, we derived Shapley value–informed thresholds and directions for each feature at each electrode contact location. Conceptually, this procedure mimics the decision process of a random forest and allows us to identify the range of SRND parameters conducive to successful visual encoding. For non-circular features, such as peak-to-trough amplitude, we sorted observations by their feature values. We then identified a change point *τ_f_* that minimized the within-segment variance of Shapley values, partitioning values into encoding-favorable versus unfavorable ranges.

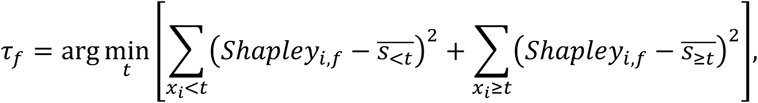

where *Sℎapley_i_*_,*f*_ is the Shapley value for sample *x_i_* and feature *f*, and 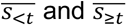 are mean Shapley values on either side of the candidate threshold *t*.

For the circular feature (i.e., instantaneous phase of preceding oscillations), we identified the angular window 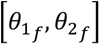 on the unit circle that minimized the variance of Shapley values inside versus outside the window.

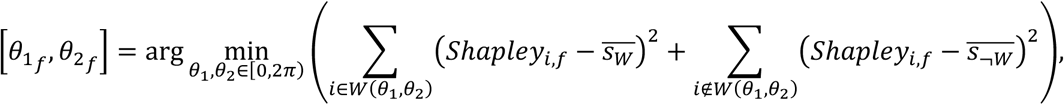

where *W*(*θ*_1_, *θ*_2_) denotes the set of samples whose feature value lies within the angular interval [*θ*_1_, *θ*_2_]. 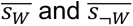 denote the mean Shapley values of samples inside and outside the candidate window, respectively.

For each feature, we defined the direction (higher vs. lower or inside vs. outside) associated with successful encoding by comparing the mean Shapley values for observations on either side of the identified threshold, or, for circular features, inside vs. outside the angular window. For non-circular features, if the mean Shapley value on one side was higher, feature values that are higher (or lower) than *τ_f_* contributed to successful encoding. For circular features, we computed the mean Shapley value within and outside the angular window 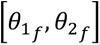. We defined the Shapley value-informed direction as the phase range yielding the higher mean Shapley value. We then quantified how strongly each observation’s feature value aligned with the encoding-favorable direction by computing a recall likelihood *L_recall_*(*i*). For a non-circular feature and a sample *x_i_*,

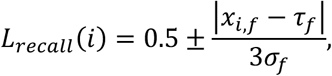

where the sign depends on the feature’s direction (“higher” or “lower” associated with successful encoding), and *σ_f_*is the standard deviation of the feature computed from the training data. For circular features, let the phase be *φ_i_* ∈ [0,2*π*) and the window *W_f_* = [*θ*_1,*f*_, *θ*_2,*f*_]. For sample *x_i_* with direction as “within”,

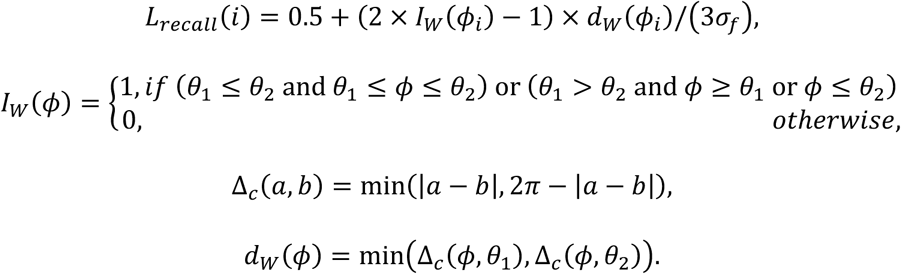

*d*_W_(*φ*) represents the minimum circular distance from an angle *φ* to either boundary of the encoding-favorable phase window. We clipped *L_recall_*(*i*) to the interval [0,1].

To validate that the Shapely value-informed thresholds and directions reflected meaningful SRND-visual encoding mappings, we compared recall likelihood averaged across electrode contacts between remembered and forgotten trials. In addition, we evaluated classification performance based on Shapley value-informed thresholds and compared the performance metrics, including balanced accuracy and ROC AUC, to their surrogate null distribution. Specifically, for each feature, we generated surrogate models by uniformly resampling feature thresholds within a range determined by the empirical minimum and maximum of the observed thresholds, while randomly permuting the direction. We then recompute recall likelihoods based on the surrogate configuration. Based on recall likelihood, for each sample and each feature, we classified it as “remembered” if *L_recall_*(*i*) ≥ 0.5, and those with *L_recall_*(*i*) < 0.5 as forgotten. We then computed balanced accuracy based on class predictions and ROC AUC based on recall likelihood at the electrode-contact, saccade, and trial levels. Please see **Model evaluation** section for computation details.

### RF regression for predicting saccade direction

To test whether SRND parameters contain information about saccade direction, we split the training and testing datasets at the saccade level and trained RF regressors. Similar to the random forest model for classifying remembered vs. forgotten trials, the random forest regressor is an ensemble of decision trees, with each tree recursively splitting the data based on feature thresholds. During training, the random forest regressor learned a set of features and split thresholds that minimize the mean squared error at each split node. After training, the output at each leaf node was the average target values across the training samples that reached that node.

The input feature set was identical to that used to predict visual encoding outcomes (i.e., SREP latency, peak-to-trough amplitude, peak time, phase, and power of preceding oscillatory activity). Training RF regressors largely followed the same framework described in the **Training RF classifiers** section, including adding Gaussian spatial jitter to electrode contact location and using 5-fold cross-validation. In addition, we verified that the RF regression model was neither overfitting nor underfitting by systematically varying key hyperparameters (**Fig. S16A**). The differences, compared to training an RF classifier to learn the SRND-visual encoding mapping, were the target variable, objective function, and model output. Because saccade direction is a circular variable, its numerical representation in radians is discontinuous at the {0, 2π} boundary. For instance, 0 rad and 2π rad correspond to the same direction but differ numerically by 2π. To allow the RF regression model to optimize the loss function in Euclidean space and avoid treating saccade direction as a linear quantity, we transformed each saccade’s direction θ into a two-dimensional direction vector in Cartesian space:

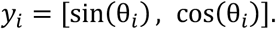

This maps angular saccade directions to continuous coordinates in Euclidean space, where opposite directions remain far apart in vector space, while equivalent directions are identical, preserving angular continuity. After transformation, we trained RF regression models using a standard mean squared error (MSE) objective function. We implement the RF regression model using *RandomForestRegressor* from the *Scikit-learn* Python package. In brief, each decision tree learned feature-split thresholds that minimized MSE over the two-dimensional direction vectors. For each sample, the trained RF model routes it to a terminal leaf in every individual decision tree. The predicted output for that sample from a single tree is the mean direction vector computed across all training samples that reached the same terminal leaf. The final ensemble prediction is the average predicted direction vector across all trees *y_i_* . We reconstructed the predicted saccade direction *θ*^¶^*_i_* for sample *i* in angular space using the two-argument arctangent function:

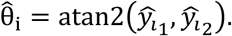

### RF regression evaluation

We evaluated RF regression model performance on held-out testing data using both MSE and mean absolute error (MAE). Specifically, we calculated the MSE between the predicted and true direction vectors.

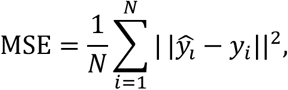

where || · || denotes the Euclidean norm. Lower MSE values indicate better prediction accuracy. In addition, we computed the MAE between the reconstructed angular predictions and actual saccade directions

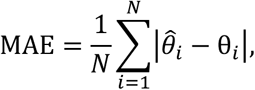

where angular differences θ̂*_i_* − θ*_i_* were wrapped into the range [−π, π] before taking the absolute value. We computed the MSE and MAE at both the location and saccade levels. For saccade-level evaluation, we averaged the predicted direction vectors across all electrodes within each saccade. To assess statistical significance, we generated null distributions for MSE and MAE by repeating the cross-validation training procedure with permuted saccade directions. Specifically, for each of 100 permutations, we randomly shuffled target direction vectors *y_i_*, repeated the RF regression model training procedure, and computed the performance metrics.

### Quantifying directional modulation

To confirm that SRND parameters did not encode saccade direction for most electrode contacts, we compared the observed testing MSE for each electrode contact against its null distribution from permutations. We calculated the contact-wise percentile rank of testing MSE relative to its null distribution. We identified direction-informative electrode contacts as those with observed MSEs below the 5th percentile of their null distribution. To complement the model-based approach, we quantified directional non-uniformity of SRND parameters. For each electrode contact and feature, we estimated the circular kernel density of the feature values as a function of saccade direction. Operationally, we discretized the angular space into 60 equal bins covering 0 − 2π radians. For each bin centered at angle **φ**_j_, we computed the mean feature value of saccades within a ±6° window around **φ**_j_. For circular features (i.e., instantaneous phase), we computed the circular mean using the *circmean* function from the *scipy.stats* Python package. We then applied von Mises kernel regression, implemented via *vonmises* from the *scipy.stats python package* (concentration parameter κ = 15).

For each feature, we quantified directional non-uniformity *D_f_* as the integral of the absolute difference between the circular kernel density and its mean across the angular domain:

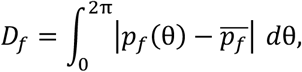

where *p_f_*(θ) denotes the kernel density estimate for feature *f*, and 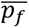 is its circular mean. A higher *D_f_* value indicates greater directional modulation.

To evaluate statistical significance, we generated null distributions of *D_f_* by permuting saccade directions 200 times and recomputing the directional non-uniformity for each permutation. We then computed the percentile rank of directional non-uniformity for each electrode contact and feature relative to the corresponding null distribution. For each feature, we computed the proportion of electrode contacts with directional non-uniformity below the 95th percentile of their respective relative null distributions, indicating a lack of direction-specific modulation.

### Neural networks for identifying saccade direction information in SRND

To validate the observed lack of saccade-direction information in SRND parameters, we implemented several complementary modeling approaches. These analyses aim to demonstrate that the lack of saccade-direction encoding across nonlinear, linear, and binary classification frameworks, and therefore confirm that SRND primarily reflects cognitive rather than motoric components of saccade processing. Specifically, we trained (1) neural networks with either saccade direction or transformed direction vector as output, (2) a linear model for angular prediction, and (3) a binary RF classifier to distinguish leftward versus rightward saccades. These analyses aimed to detect linear or nonlinear mappings between SRND parameters and saccade direction.

As a nonlinear control model, we implemented a feedforward neural network consisting of three fully connected hidden layers, each followed by a Rectified Linear Unit activation function using the *PyTorch* Python package. The input layer received the same feature set as the RF models, including electrode spatial coordinates and SRND parameters, described in the **Training and testing data structure** section. The network’s output layer contained two units representing the two-dimensional Cartesian saccade direction vector [sin(θ), cos(θ)] . To ensure that predictions represented saccade directions, we required the network to normalize its outputs to the unit circle:

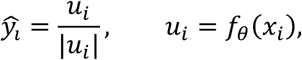

where *f*_θ_(*x_i_*) denotes the network output for input *x_i_*. We trained this neural network to minimize MSE between predicted and true direction vectors. Consistent with previous machine learning model training and evaluation, we used the 5-fold cross-validation procedure. We trained the network using the Adam optimizer from the *PyTorch* Python package with a learning rate of 1 × 10^−3^, a batch size of 256, and up to 50 epochs.

To confirm that the transformation of saccade direction into a two-dimensional Cartesian direction vector did not prevent the neural network from learning the relationship between SRND parameters and saccade direction, we implemented a complementary neural network trained to minimize angular loss directly. This model had the same three-layer architecture and inputs as above, but used a single-unit output layer representing the angular direction in radians. We used a loss function to represent the wrapped angular distance between the predicted direction (*θ̂*) and true directions (*θ*):

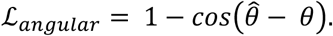

This formulation penalizes large angular deviations and is continuous at the 0 − 2π boundary. We evaluated both neural networks at the electrode contact and saccade levels using MSE, computed analogously to the RF regression evaluation (see the **RF regression evaluation** section). For the network with two outputs, we reconstructed the predicted angular direction from the predicted direction vector using the two-argument arctangent function. We then computed the MSE between reconstructed and true saccade directions after wrapping angular differences into the range [−180°, 180°]. To evaluate whether prediction accuracy exceeded chance, we generated null distributions of MSE by training neural networks with the same architecture on data with permuted saccade directions. Specifically, we randomly shuffled target direction vectors [sin(θ), cos(θ)] across saccades 100 times and repeated the full cross-validation training and evaluation procedure for each permutation.

### Kernel regression by minimizing the von Mises negative log-likelihood

To test whether SRND parameters could linearly explain saccade direction, we implemented a linear regression by minimizing the von Mises negative log-likelihood. The von Mises distribution is the circular analog of a Gaussian distribution for angular data, modeling angular data defined on the unit circle while accounting for the periodicity at the 0–2*π* boundary. Formally, we modeled the saccade direction θ*_i_* as a circular variable whose mean direction µ*_i_* depends linearly on the SRND features:

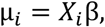

where *X_i_* is the feature vector comprising SRND parameters and electrode coordinates, and β is the vector of regression coefficients to be estimated. Under the von Mises assumption, the probability density of observing θ*_i_* is given by

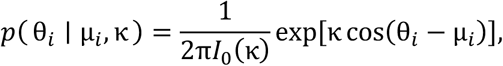

where κ is the concentration parameter which we set to 1. *I*_0_(κ) is the modified Bessel function of the first kind of order 0. We estimated the regression parameters by minimizing the von Mises negative log-likelihood:

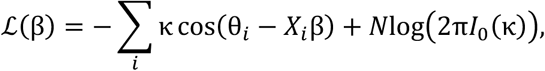

To fit this model, we used the BFGS optimization algorithm implemented via the *minimize* function from the *scipy.optimize* Python package. We obtained the predicted saccade direction from the model as

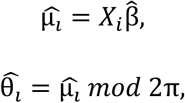

where β̂ denotes the estimated model parameters, and *mod* denotes the modulo operation, which ensures that predicted directions remain within the circular domain [0,2π).

Consistently, we evaluated model performance using contact- and saccade-level MSEs between predicted and true saccade directions. We assessed the statistical significance of prediction accuracy by comparing MSEs to their null distributions generated via 100 permutations of saccade labels.

### Binary classification of left vs. right saccades

To parallel the classification framework used for predicting successful visual encoding, we also trained a balanced RF classifier to discriminate between leftward and rightward saccades. We binarized saccade directions as left and right based on their angular position. All procedures for data splitting, feature selection, model training, evaluation, and statistical testing followed those described in the **Random forest classifier**, **Training and testing data structure**, **Training RF classifiers**, and **Model evaluation** sections.

### Prediction of subsequent saccade direction

Finally, to determine whether SRND reflects neural computations underlying the planning of subsequent eye movements, we trained an RF regressor to predict the direction of the next saccade from SRND parameters at the current saccade. We defined subsequent saccades using combined temporal and spatial proximity criteria: (1) For the temporal proximity criterion, for each valid saccade, we searched for the next non-fixation saccade occurring within the same trial and within 500 ms of the current saccade offset; (2) The spatial proximity criterion required that the difference between the gaze positions at saccade offset for the current and subsequent saccades be less than 20% of the screen dimension in both the x and y axes.

For each valid pair of saccades, we used the direction of the next saccade as the target variable in the RF regression model. Model training and evaluation followed the same procedures as described in the **RF regression for predicting saccade direction** section. We quantified prediction performance using MSE between predicted and true subsequent saccade direction vectors and MAE between predicted and true subsequent saccade directions in radius. We assessed statistical significance by comparing observed performance against null distributions derived from models trained on data with permuted saccade directions.

### Quantification and Statistical Analysis

All data were analyzed and plotted with MATLAB and Python. For comparison between two conditions, when assumptions of normality and equal variances were met, we used Student’s unpaired t-test. When samples were paired (e.g., within-subject or within-electrode comparisons) and these assumptions were met, we used paired Student’s t-tests. When these assumptions were not met, we used the Mann–Whitney U test for comparisons between independent samples, and the Wilcoxon signed-rank test for paired comparisons. For comparison of the mean direction between two (or more) circular distributions, we used Watson-Williams F test. For comparison of the full circular distributions (i.e, differences in shape), we used Mardia–Watson–Wheeler test. For testing if the distribution of intertrial correlation differed from baseline, we used Kolmogorov-Smirnov tests.

For linear regression analyses, we assessed the statistical significance of regression coefficients using two-sided t-tests. For correlation analyses, we converted Spearman’s or Pearson’s correlation coefficient to t-statistics and assessed their significance against the Student’s t-distribution. For testing if a learning model’s performance exceeded chance, we performed permutation testing, in which data labels were randomly permuted to generate a surrogate null distribution; and we compared the true performance metric value to this distribution to obtain a percentile-based p-value.

Statistical significance is defined as NS (non-significant), ∗ p < 0.05, ∗∗ p < 0.01, ∗∗∗p < 0.001. We report statistical details in the Results section and figure legends, including the statistical tests used, statistics, effect size, and exact value of n. Effect sizes are reported as: regression coefficients for linear models, Pearson’s or Spearman’s r for correlation analyses, Cohen’s d or rank-biserial correlation for pairwise comparisons, and percentile score relative to the null distribution for permutation tests. When more than 10 comparisons were used to support a conclusion, Bonferroni correction was applied.

## Data Availability

All data produced in the present study are available upon reasonable request to the authors

## Declaration of AI-assisted technologies

The author(s) used ChatGPT (GPT-5.4) to assist with preparing code for a public repository and with drafting documentation. The original analysis and coding were performed by the authors. After using these tools, the authors reviewed and edited the content as needed and take full responsibility for the content of the published article.

## Funding

This work was supported by NIH/NIMH (R01-MH120194), NIH/NIBIB (P41-EB018783, R01-EB026439), McDonnell Center for Systems Neuroscience, Fondazione Neurone, NIH/NINDS (U24-NS109103, U01-NS108916, U01-NS128612, R21-NS128307), and NIH/NEI (R01-EY037195). The manuscript was partly edited by the Scientific Editing Service of the Institute of Clinical and Translational Sciences at Washington University, which is supported by an NIH Clinical and Translational Science Award (UL1 TR002345). We thank Robert T. Knight for his helpful feedback, which clarified the conceptual contribution and significance of the work.

## Author contributions

Conceptualization: G.T.; Data curation: G.T., P.B., and J.T.W.; Formal analysis: G.T.; Funding acquisition: E.C.L., P.B., J.T.W., C.S.I., J.R.M., and S.B.H.; Investigation: G.T., P.B., J.T.W., and E.C.L.; Methodology: G.T., P.D., H.C., Y.L., and Z.L.; Software: G.T., P.B., and J.R.S.; Supervision: P.B., J.T.W., and E.C.L.; Validation: G.T., P.B., H.C., P.E.C., S.S.S., H.P., X.L., Z.L., and E.C.L.; Visualization: G.T., P.D., and P.B.; Writing – original draft: G.T.; Writing – review & editing: G.T., J.T.W., P.B., and E.C., Task design: J.R.S., K.L.W., M.K.H., and J.M.C.

### Competing interest

Eric Leuthardt holds equity in Aurenar Inc. Gansheng Tan, Peter Brunner, and Eric Leuthardt filed a provisional patent related to this work in November 2025 (Application number: 63/910,588). The other authors declare no competing interests.

### Data, code, and materials availability

All analysis scripts and data for statistical analysis and figure plotting are available on a GitHub repository (https://github.com/neurotechcenter/Saccade-related-evoked-potential-predicts-visual-memory). Raw imaging and electrophysiological data can be de-identified and provided to qualified researchers upon request, given the large file size (∼95 TB) and in accordance with our IRB-approved protocol. Anonymized data for one subject, and a script illustrating the data-processing pipeline are available on a GitHub repository (https://github.com/neurotechcenter/Saccade-related-evoked-potential-predicts-visual-memory).

## Notes

### Competing Interest Statement

The authors have declared no competing interest.

### Clinical Trial

NCT05065450

### Author Declarations

Washington University Institutional Review Board (IRB protocol number: 202104033-1001 gave ethical approval for this work.

### Summary of Updates

Streamline Result section to improve the flow

