## Supplementary materials for "Mind’s eye: widespread saccade-related evoked potentials support visual memory encoding in humans"

Science Advances
AAAS

### Supplementary Materials for

#### Mind's eye: widespread saccade-related evoked potentials support visual memory encoding in humans

Example: Gansheng Tan *et al.*

*Jon T. Willie and Peter Brunner

**This PDF file includes:**

Supplementary Text
Figs. S1 to S15
Tables S1 to S4
Movies S1
Audio NA
Data NA
References (1 to 1) (if applicable—these should refer only to references in the SM)

**Other Supplementary Materials for this manuscript include the following:**
Movies S1

#### Supplementary Text

##### Brief amygdala stimulation did not exert carryover effects upon visual memory encoding and saccades

This study is part of a clinical trial that investigated the effect of amygdala stimulation on declarative memory (NCT05065450). Brief electrical stimulation of the amygdala was delivered immediately following image presentation in half of the visual memory encoding trials, followed by a 5-s intertrial interval. To study the saccadic modulation in the normative visual encoding process, we restricted analyses to trials that met the following criteria: (1) no amygdala stimulation was delivered during the trial; (2) the subject maintained their gaze on the monitor for more than half of the trial duration, as determined from eye-tracking data; and (3) the subject pressed one of the response buttons. Applying these criteria yielded 1130 valid visual encoding trials, comprising 7292 saccades (**Table S1**).

While amygdala-stimulated trials were excluded, we assessed whether recent amygdala stimulation exerted carryover effects upon subsequent non-stimulated trials by comparing saccade behavior and recognition memory performance between trials with and without a preceding amygdala-stimulated trial. The effect size (i.e., Cohen's d) for duration differences between trials with and without a preceding amygdala stimulation trial was trivial (-0.015 for saccades and -0.024 for fixation, **Fig. S1C–D**). The circular distribution of saccade direction did not differ significantly between trials following an amygdala stimulation trial (N = 3657) and those not following amygdala stimulation (N = 3635, Watson–Williams test, p=0.879, F = 0.023, **Fig. S1A–B**). The correct recall rate for trials that followed a preceding amygdala-stimulated trial, and those that did not, were both 75% (**Fig. S1E)**. These results reject the possibility that recent brief amygdala stimulation exerted confounding carryover effects upon visual encoding and saccades during the trials included in subsequent analyses.

##### Threshold-independent estimate of SREP prevalence

To obtain a threshold-independent estimate of SREP prevalence across regions, we varied the thresholds from the mean to the mean + 4 SD of the control area under the ROC curve, and integrated the resulting SREP prevalence over this range, yielding a summary prevalence metric. This analysis revealed a similar spatial pattern in SREP prevalence (**Fig. S3**). We ranked brain regions by the area under the ROC curve for peri-saccadic intertrial correlations and by their difference from the control area under the ROC curve (**Fig. S4C–E**). The differences in area under the ROC curve were generally higher in the lateral, basal, and polar temporal lobes and the parietal lobes, followed by the frontal lobes and subcortical regions. The electrode contact-level spatial pattern of differences in area under the ROC curve was consistent with previous findings (**Fig. S5A–E**). The peri-saccadic area under the ROC curve averaged across polarities was also higher than baseline for LFPs recorded from micro-wires, but this difference was less pronounced compared to LFPs recorded from macro electrode contacts (**Fig. S5F–G**, Cohen's d = 0.227, two-tailed paired t-test, N = 115, t statistic = 2.088, p = 0.039; subregion-level comparisons also showed larger differences in area under the ROC for macro than micro recordings, for examples, $N_{micro}^{right\ hippocampus} = 16$, $N_{macro}^{right\ hippocampus} = 162$, $\overline{Difference}_{micro}^{right\ hippocampus} = 0.034$, $\overline{Difference}_{micro}^{right\ hippocampus} = 0.075$; $N_{micro}^{left\ orbitofrontal} = 8$, $N_{macro}^{left\ orbitofrontal} = 112$, $\overline{Difference}_{micro}^{left\ orbitofrontal} = 0.088$, $\overline{Difference}_{micro}^{left\ orbitofrontal} = 0.132$). These results demonstrate that SREPs are widespread across the sampled human cerebrum, but are pronounced especially in the temporal and parietal lobes.

##### Interplay between ongoing oscillations and SREP

Active sensing theory proposes that neuronal oscillations create cyclic windows of efficient sensory processing that are temporally coordinated with sampling behaviors(*32, supplementary reference 2*). We tentatively interpreted the observed phase separation between positive and negative SREPs as evidence that an oscillatory cycle contains two such windows for visual memory encoding. This interpretation predicts that the oscillation phase associated with the same SREP polarities align across saccades. To test this prediction, we quantified phase alignment using the inter-trial phase coherence (ITPC), separately for positive and negative SREPs. For both polarities, ITPC increased monotonically from 2 OCs before the SREP onset latency, then decreased after the SREP peak time (**Fig. S8G**). The effect sizes (i.e., Cohen's d) between ITPC at 2 OCs before SREP onset latency and subsequent 4 time points (from 1 OC before SREP onset latency to 1 OC after SREP peak time) were 1.214, 1.625, 1.739, 1.704, and 1.427. These findings support the notion that SREPs are embedded in a broader window of saccade-timed neural coordination.

While phase alignment indexes the temporal coordination between underlying neural oscillations and SREPs, oscillatory power reflects the degree of synchronous local population activity accompanying SREPs(*33, supplementary reference 3*). We therefore examined the temporal dynamics of oscillatory power during SREPs. To enable comparison across electrode contacts and subjects, we normalized the oscillatory power to its magnitude at two OCs before the SREP onset latency. Oscillatory power increased from 2 OCs before the SREP onset latency and remained elevated at that level throughout the SREP period (**Fig. S8F**). To determine whether phase alignment and oscillatory power during SREPs reflect dissociable neural processes, we compared their temporal dynamics by quantifying their correlation across time points. We found the strongest positive correlation at SREP onset latency (**Fig. S8H**, $R^2 = 0.274$). The correlation was weaker at time points preceding and following SREP onset, with $R^2$ = 0.022 at 1 OC before SREP latency, $R^2$ = 0.118 at 0.5 OCs before SREP latency, $R^2$= 0.198 at the SREP peak, followed by a decline to $R^2$ = 0.046 at 0.5 OCs after the SREP peak. These results show partially divergent temporal profiles of phase alignment and oscillatory power.

#### Intra- and inter-contact variability in SREP peak amplitude

Across electrode contacts, peak amplitude of cluster-level SREPs for positive and negative polarities was significantly and negatively correlated, reflecting the opposing polarities (**Fig. S10B–C**, Pearson's correlation coefficient = -0.365, $p < 0.001$). For a pair of electrode contacts, the effect size between their single-trial SREP peak amplitude was positively correlated with the difference in peak amplitude in their averaged SREP (**Fig. S10D**, Pearson's correlation coefficient = 0.510 for negative SREPs, Pearson's correlation coefficient = 0.479 for positive SREPs, $p < 0.001$ for both polarities, based on t-distribution). Following the same reasoning as in the SREP onset latency analysis above, we quantified intra- and inter-contact variability in SREP peak amplitude and found that mean intra-contact variability was higher than mean inter-contact variability (**Fig. S10A**). While greater intra-contact variability suggests that single-trial SREP peak amplitude mostly reflects dynamic neural processes associated with each saccade, it does not preclude the presence of systematic anatomical differences in cluster-level average SREP peak amplitude.

#### Preconditions for characterizing the SREP pathway using onset latency

To use cluster-level averaged SREP onset latency for characterizing spatial structure in SREP onset latency, two preconditions should be satisfied: across electrode contacts, (1) positive and negative SREP onset latencies show positive correlation, such that pooling positive and negative SREPs does not obscure location-specific latency patterns; and (2) inter-contact differences in cluster-level averaged SREP latency show positive correlation with differences at the single-trial level, ensuring that the cluster-level averages reflect location-specific onset latency. Our evaluation of the preconditions revealed that, across electrode contacts, onset latencies for positive and negative SREPs were positively correlated (Pearson $r = 0.436$, $p < 0.001$; **Fig. S9A–B**), and we found a significant positive correlation between inter-contact differences in clustered-level averaged SREP onset latency and the corresponding effect sizes computed from single-trial SREPs (**Fig. S9C**, Pearson $r = 0.436$, $p < 0.001$, t-test). These results satisfy the preconditions for characterizing the spatial structure in cluster-level SREP onset latency.

#### Shapley value–informed thresholds and directions capture the relationship between SRNDs and successful memory encoding

Using SRND parameters across saccades as inputs to the trained random forest classifier did not substantially improve predictions of memory outcomes. If Shapley value–informed thresholds and directions faithfully represent the SRND parameter-memory outcome relationship learned by the random forest classifier, trial-level prediction using these thresholds and directions should similarly not improve beyond saccade-level prediction. We applied the same procedure described above, that is, calculated the proportion of SRND parameters falling within location-specific encoding-conducive ranges, now extended across all saccades within a trial. At the trial level, the area under the ROC curve and optimal balanced accuracy of separating remembered from forgotten trials using Shapley value–informed thresholds and directions were 0.801 and 0.732, respectively (**Fig. S12B**). While these two model performance metrics exceeded all values in the surrogate control distribution, they did not surpass saccade-level prediction, which supports that Shapley values–informed thresholds and directions capture the relationship between SRNDs and successful memory encoding

#### SREPs did not generally encode saccade direction

We have demonstrated that SREPs reflect neural states supporting visual encoding and have characterized SREP profiles associated with successful visual encoding. The remaining question is whether these SREPs represent saccadic corollary discharge or a novel memory-related mechanism. If SREPs were a narrowly defined corollary discharge signal, they should convey information about saccade direction. We tested this hypothesis using random forest regressors to examine whether SRND parameters could predict saccade direction (see **MATERIALS AND METHODS:** RF regression for predicting saccade direction). Because saccade direction is a circular variable, we transformed saccade direction $\theta$ into a continuous vector $[sin(\theta), cos(\theta)]$, which allows the use of Euclidean distance as a loss function while preserving the circular nature of the variable. **Fig. S15A** illustrates the model architecture and training and evaluation pipeline.

To assess whether SRNDs generally contained information about saccade direction, we compared the model's prediction error on held-out testing data to null distributions generated by permuting saccade directions. Across 5-fold CVs, the observed testing mean squared error (MSE) for $sin(\theta)$ fell at the $65^{th}$ percentile of the null distribution, and that for cos(θ) fell at the $93^{rd}$ percentile (**Fig. S15B**). We obtained the predicted saccade direction using the two-argument arctangent function, $atan2$, and found that the mean absolute angular error between predicted and true saccade direction was higher than the surrogate control distribution, which indicated that SRND parameters were not predictive of saccade direction (**Fig. S15C**). We next assessed if leveraging SRND parameters across electrode contacts could reveal directional encoding. For each saccade, we obtained the predicted saccade direction from the averaged predicted $[sin(\theta), cos(\theta)]$ vectors across electrodes (**Fig. S15D**). The testing mean absolute error (MAE) corresponded to the $52^{nd}$ percentile of the null distribution, indicating chance-level performance. To verify that this result was not due to suboptimal hyperparameter choices, we systematically evaluated the effect of key hyperparameters, including the number of trees, the maximum tree depth, and the minimum leaf size, on model performance. Although increasing the maximum depth and decreasing the leaf size allowed the model to be more complex and better fit the training data, the electrode contact-level and saccade-level model performance on the testing data remained consistently poor, suggesting that the failure to predict saccade direction was not due to model limitations (**Fig. S16A**). These results provide initial evidence that SRND parameters do not generally encode information about saccade direction.

Subsequently, we examined whether SRNDs in any contacts contained directional information by determining for each contact, the percentile rank of its saccade-level testing MSE within its respective null distribution. Only 5% of electrode contacts achieved a testing MSE below the 5th percentile of their null distributions (**Fig. S15E**). We used permutation testing to identify direction-informative electrode contacts by comparing the percentile rank of each contact's saccade-level test MSE within its null distribution against a Bonferroni-corrected significance threshold ($\alpha = 0.05$). **Fig. S15F** shows the ratio of saccade direction-informative electrode contacts across anatomical regions. No region exceeded a 3% ratio of direction-informative electrode contacts. A cluster of eight microwires near the left hippocampus and brainstem was initially assigned to the brainstem by automated localization, and one of these eight microwires showed directional information. Because we lack a meaningful representation of electrodes in the brainstem, we excluded it from the anatomical analysis of direction-informative contacts. To determine which SRND parameter showed directional modulation, we estimated the circular kernel density of the SRND parameters' values as a function of saccade direction (**Fig. S15F**). For each SRND parameter, we quantified directional modulation as the non-uniformity of feature values across angular space, measured as the integral of the absolute deviation from the mean. **Fig. S15G** shows the distribution of non-uniformity for the five SRND parameters. We compared them against the null distribution generated by shuffling SRND-direction pairings. For all five SRND parameters, the non-uniformity value in more than 95% of electrode contacts was below the 95th percentile of their respective null distributions (**Fig. S15G**). These results suggest that most, if not all, SRNDs across the sampled cerebrum do not convey information about saccade direction.

To confirm that the absence of directional information in SRND parameters was not an artifact of the model architecture, learning target, or prediction granularity, we implemented a series of control models: (1) a fully connected neural network predicting the trigonometric representation of saccade direction, (2) a neural network trained to minimize angular loss, (3) a von Mises kernel regression, and (4) a random forest classifier for binary saccade direction (left vs. right). We obtained null distributions of performance metrics from these models trained on permuted data. We confirmed the absence of directional information in SRND across all four models and learning schemes (See **Absence of directional information in SRND parameters across machine learning models and learning schemes** below).

To further distinguish SREPs from corollary discharge, we assessed whether SREPs reflected the motor planning for the subsequent saccade, that is, whether they encoded the direction of the upcoming saccade. We trained and evaluated random forest regressors for predicting the direction of the next saccade using SRND parameters and electrode locations. The electrode contact-level and saccade-level mean absolute errors were 90° and 90°, respectively (**Fig. S16J**). These errors corresponded to the 42nd and 39th percentiles of the null distribution, respectively, suggesting that SREPs were unlikely to reflect prospective oculomotor planning. Overall, our data do not support the interpretation of SREPs as a corollary discharge signal.

#### Absence of directional information in SRND parameters across machine learning models and learning schemes.

The first control model, a neural network, predicted the sine and cosine of the saccade direction using SRND parameters and electrode coordinates (**Fig. S16B**). The observed electrode contact-level mean squared errors (MSEs) for $sin(\theta)$ and $cos(\theta)$ were 0.924 and 1.035, corresponding to the 71st and 28th percentiles of their respective null distributions (**Fig. S16C**). The MSEs for saccade directions were

10404 and 10263 $deg^2$ at the electrode contact and saccade levels, corresponding to the 28th and 30th percentiles of their null distributions, respectively (**Fig. S16D**).

The second control model, a neural network, used the same architecture as the first control model but with an angular loss function to ensure that the model training was not constrained by the trigonometric target representation (**Fig. S16E**). This model yielded MSEs of 1066 and 10581 $deg^2$ at the electrode-contact and saccade levels, corresponding to the 17th and 37th percentiles of their null distributions, respectively (**Fig. S16F**).

The third control model used von Mises kernel regression, which natively handled the circular dependent variable (i.e., saccade direction) (see **MATERIALS AND METHODS:** Kernel regression by minimizing the von Mises negative log-likelihood). This model also failed to predict saccade direction above chance, with MSEs of 10783 and 10520 $deg^2$ at the electrode contact and saccade levels, corresponding to 22nd and 22nd percentiles of their null distributions (**Fig. S16G**).

The fourth control model, a random forest classifier, predicted binarized saccade direction (i.e., left vs. right). This approach reduced the complexity of the prediction problem and paralleled our earlier prediction of successful visual encoding (**Figure 5B**). This random forest classifier performed at the chance level. The electrode contact-level balanced accuracy was 0.501, corresponding to the 83rd percentile of its null distribution. The saccade-level area under the ROC curve and balanced accuracy were 0.517 and 0.502, corresponding to the 52nd and 73rd percentiles of their null distribution, respectively. (**Fig. S16I**). The absence of directional information in SRND parameters was robust across all four models and learning schemes.

#### The duration of SREP consistency

To quantify the duration for which SREP waveforms remained consistent across trials, we systematically varied the pre-saccadic and peri-saccadic epoch duration and compared their intertrial correlations. We defined the pre-saccadic epoch as starting 1 second prior to saccade onset and with the same length as the peri-saccadic epoch (**Fig. S6A**). To quantify SREP consistency differences between pre- and peri-saccadic epochs, we computed the difference in the area under the cumulative distributions of the intertrial correlation curve (AUC) as follows: for correlations smaller than the crossing point, we calculated the difference by subtracting pre-saccade AUC from peri-saccade AUC; for correlations greater than the crossing point, we calculated the difference by subtracting peri-saccade AUC from pre-saccade AUC. Aligning with the main findings, we found that SREPs were more consistent than pre-saccadic LFPs (**Fig. S6B**, $t = 5.349$, $p < 0.001$, Cohen's $d = 0.274$, $n = 382$, two-sided t-test). This result is consistent across analysis epochs of varying durations. Interestingly, distributions of intertrial correlations were less normally distributed when the analysis epochs were shorter, for both pre- and peri-saccadic EP (**Fig. S6A**).

These results imply that SREPs are transient. As we increased the epoch duration, we incorporated ongoing activity unrelated to the saccade, thereby reducing waveform consistency. The same pattern observed in pre-saccadic neural field potentials may result from coordinated visual memory-encoding processes preceding saccades. Our observed clustering of oscillation phases preceding saccades supports this view (**Figure 3D**). A separate methodological concern is that shorter windows, by including fewer data samples, may yield extreme intertrial correlation values(*57, supplementary reference 1*). To rule out this possibility, we performed an additional surrogate analysis by shuffling data for each saccade to extract pre- and peri-saccadic LFP waveforms. We found that the separation effect size, which measures how peri-saccadic LFP differs from baseline LFP, was significantly reduced in surrogate data compared to real data ($t = -65.086$, $p < 0.001$, Cohen's $d = -1.804$, two-sided t-test, $N = 2604$, **Fig. S6C**). The above analyses suggest that the LFP waveform following saccades is not spontaneous but represents consistent saccadic modulation. This conclusion is further supported by our finding that SREPs predicted successful visual encoding.

#### Random forest classifier's generalizability to individual subjects

We evaluated the random forest classifier's generalizability to individual subjects by computing electrode-contact- and saccade-level balanced accuracies, averaged across cross-validation iterations, separately for each subject's test data. For each subject, we generated a null distribution of balanced accuracy using models trained on permuted labels. **Fig. S11E** shows the percentile rank of balanced accuracy relative to the null distribution for each subject. The observed saccade-level balanced accuracy exceeded the null distribution in 6 out of 8 subjects, indicating relatively good generalizability. For the remaining two subjects, saccade-level balanced accuracy corresponded to the $83^{rd}$ and $72^{nd}$ percentiles of the null distribution, indicating performance above chance but with limited generalizability. This is not unexpected: although the full cohort provided broad coverage across the cerebrum, electrode coverage for individual subjects was often more limited on clinical grounds, for example, restricted to a unilateral hemisphere containing the suspected seizure onset zone. We also note that these two subjects had the fewest valid remembered and tested visual encoding trials (12 and 14), which might limit the model's ability to fully learn the mapping between SRND parameters and successful encoding in the brain regions sampled in these individuals.

#### Relationship between saccadic behavior and memory outcome

A potential confound arises if differences in saccadic behavior existed between remembered and forgotten trials and those behaviors were correlated with SRND parameters. In that case, the random forest classifier could achieve above-chance prediction by exploiting saccadic behavior-memory associations, in addition to SRNDs. Although our earlier analyses found no association between saccadic behaviors and SRND parameters, we took an additional precaution before training the classifier by verifying that saccadic behaviors themselves did not predict visual encoding (**Fig. S11J**), and we found comparable saccade eccentricity and duration between saccades during remembered and forgotten trials ($N_{remembered}$ = 3913, $N_{forgotten}$ = 1227, t = -0.325, p = 0.745, Cohen's d = -0.011, t-test for saccade eccentricity; t = -1.237, p = 0.216, Cohen's d = -0.0039, t-test for saccade duration). To assess differences in saccade direction between remembered and forgotten trials, we estimated circular densities of saccade angles and quantified between-condition differences as the absolute area between the two densities over [–π, π]. In contrast, a permutation test, in which condition labels were shuffled, showed no significant difference in saccade direction between saccades in remembered and forgotten trials (p = 0.862).

#### Assessing the potential linear relationship between the SRND parameters and visual encoding

Because the anatomical organization of brain regions and their associated functions is highly complex and not expected to follow a linear mapping, we used non-linear models (i.e., random forest models) to learn the mapping from SRND parameters recorded at specific brain regions to visual encoding. There may nevertheless be a linear relationship between the SRND parameters and visual encoding. To interrogate this possibility, we trained logistic regression models using the same inputs and labels as in training random forest models, except that we substituted peak-to-trough amplitude with its absolute value. This was done to avoid multicollinearity, as polarity information is already captured by the preceding oscillation phase. The logistic regression performed well above chance, but not as well as the random forest model (**Fig. S11I**). For testing data, the electrode-contact-level balanced accuracy was 53.1%, the saccade-level ROC AUC was 63.2%, and the saccade-level balanced accuracy was 59.5%. Assuming a linear relationship, the weight coefficient for each SRND parameter indicates that a higher absolute SREP peak-to-trough value was associated with remembered trials, aligning with the mapping identified in the random forest models (**Figure 6I**).

#### Relationship between encoding-conducive neural oscillation phase interval and SREP polarity

At 0.5 OCs before SREP onset latency, oscillations in the trough-to-peak rising phase (i.e., [-π, 0] radians) were associated with successful memory encoding across most brain regions, regardless of SREP polarity. We refer to this range of oscillation phase as the encoding-conducive phase interval. Because the pre-SREP oscillation phase was coupled with SREP polarity, it is possible that the encoding-conducive phase interval was disproportionately populated by SREPs of one polarity, which would predict a memory encoding outcome difference between polarities. Contrary to this prediction, across electrode contacts, memory encoding outcome associated with positive and negative SREP did not differ (mean [95% CI from bootstrapping]: Positive, 74% [73%, 75%]; Negative, [73%, 75%]). To identify the source of this contradiction, we examined the joint distribution of oscillation phase and the corresponding Shapley values. Across electrode contacts, although pre-SREP phase distributions differed between positive and negative SREPs (centering near 0 for negative SREPs and near $\pi$ for positive SREPs), the encoding-conducive phase interval was similar across polarities (**Fig. S14C–D**). Shapley value for the pre-SREP oscillation phase showed subtle distributional differences between polarities: moderate positive Shapley values (≈0.01) were more probable for negative-polarity SREPs, whereas higher Shapley values (≈0.03) were more probable for positive-polarity SREPs (**Fig. S14E**). These differences balanced out, yielding equivalent memory encoding outcomes across SREP polarities.

The following sections provide additional details for the methods used in this study.

#### Behavioral paradigm

Participants completed visual encoding tasks at their bedside during their stay in the EMU. Before the main task began, participants received standardized instructions and completed a brief training session to become familiar with the task structure. The images presented during training were not repeated during the main task, and data from the training session were excluded from the analysis.

After training, participants began the encoding task. A series of trial-unique color object or scene images was presented on a 27-inch monitor (TUF GAMING VG27AQL1A, ASUS, USA) mounted on a mobile cart positioned next to the patient's bed. Each image was displayed at the center of the screen for 3 seconds, followed by a 6-second inter-stimulus interval (ISI). During the image presentation, the subjects used a keyboard to indicate either "Like" or "Dislike" for object images or "Indoor" or "Outdoor" for scene images. This design was intended to maintain subjects' engagement and attention level throughout the task. The image occupied 5.56 × 5.56 inches on the screen at a viewing distance of approximately 60 cm, corresponding to a visual angle of 13.1°. Participants were instructed to freely view each image and remember it for a later memory test. A fixation cross was presented during the ISI, but participants were not required to fixate on it. In brain stimulation trials, 1 second of electrical stimulation was delivered to the amygdala during the first second of the 6-second ISI. Only non-stimulation trials were included in the current analysis. Approximately 24 hours after completing the encoding task, participants performed a self-paced recognition memory test. The test consisted of 160-240 previously seen images and 80 novel images, randomly intermixed. Each image was displayed for a mandatory 1-second viewing period, after which participants used a keyboard to indicate whether they had seen the exact image before ("Sure No", "Maybe No", “Sure Yes”, or “Maybe Yes”).

For subsequent analysis, we assigned binary labels (Remembered and Forgotten) to encoding figures by considering the ground-truth stimulus status (previously seen vs. novel) and the participant’s response. We labeled encoding trials “Remembered” when participants correctly judged the image’s status (i.e., correctly judged if they had seen the images). To evaluate recognition independent of subjective confidence, we labeled previously seen images responded to with “Sure Yes” and “Maybe Yes” as “Remembered “and novel images responded to with “Sure No” and “Maybe No” as “Remembered”, otherwise “Forgotten”. No feedback was provided during the memory test.

#### Electrode implantation and clinical procedure

The current study is part of the Amygdala Memory Enhancement trial (NCT05065450). All patients underwent implantation of multi-contact depth electrodes (sEEG Depthalon depth electrode, PMT® Corporation, MN, USA) for seizure monitoring. Each electrode had 8–16 contacts (2 mm cylindrical contacts length, 0.8 mm diameter, spaced 3.5 mm apart center-to-center). In 6 patients, two hybrid depth electrodes with macro- and micro-contacts (Behnke Fried Depth Electrode, Ad-Tech® Medical Instrument Corporation) were implanted. Each hybrid depth electrode has a microwire bundle in addition to 8 standard macro contacts. The microwire bundle consists of nine 40-µm-diameter wires, which were trimmed to extend up to 5 mm beyond the tip of the macroelectrode during implantation. Eight of the microwires are coated in polyamide insulation up to the tip, resulting in an impedance of 100–300 kΩ; the ninth microwire is uninsulated, serving as a reference.

The clinical team planned the electrode placement to ensure appropriate coverage of the brain for seizure network localization. T1-weighted imaging (MPRAGE sequence, voxel size $1.0 \times 1.0 \times 1.0\ mm^3$) was obtained in the presurgical workup and loaded into the planning workstation. The neurosurgeon implanted electrodes, guided by a stereotactic robotic system (ROSA ONE Brain, ZIMMER BIOMET, IN, USA). After surgery, patients underwent a CT scan, which was co-registered to their pre-operative MRI to obtain precise 3D coordinates for each electrode contact. We describe the electrode localization method in detail in the **MATERIALS AND METHODS: Anatomical localization of the electrode**. Patients were then awakened from general anesthesia and transferred to the Epilepsy Monitoring Unit (EMU) for long-term intracranial electroencephalography (iEEG) monitoring. The duration of EMU stay varied across patients, typically ranging from several days to weeks, depending on seizure frequency. During this period, the research team introduced the study to the patients and obtained written informed consent before beginning any experimental recordings. Following consent, research sessions were scheduled in coordination with the clinical team to ensure no disruption to standard clinical care. Standard clinical protocols may taper or withdraw antiseizure medications to provoke seizures during iEEG monitoring. In some cases, partial sleep deprivation was employed to increase seizure likelihood.

All electrophysiological data will be collected through the EMU’s clinical intracranial neural recording system and other devices that have been approved by the FDA to be sold for studies in human subjects or are commonly approved and used at other research institutions.

#### Anatomical localization of the electrode

We localized electrodes based on post-operative CT scans co-registered to pre-operative T1-weighted MRI scans (0.5 mm isotropic resolution). To co-register the CT volume to the MRI volume, we first converted the MRI and CT scans from DICOM to NIfTI format using *dicm2ni*, a tool from NeuroImaging

Tools and Resources Collaboratory. We rigidly transformed the CT volume to place its geometric center at the world coordinate origin. This pre-processing step ensured robust rigid-body coregistration using the *SPM12* mutual-information-based registration. For each subject, the pre-operative MRI was treated as the anatomical reference, and the CT image was coregistered to it.

We first manually localize macroelectrode contacts using Freeview, which is the visualization tool within the FreeSurfer software package. To improve precision and reproducibility, we implemented a semi-automated algorithm to align the manually identified contact positions with the most intense local signal in the CT volume, indicative of metal artifact. For each manually identified macro contact, we extracted a local CT subvolume (5×5×5 voxels, corresponding to approximately 16 $mm^3$) and applied a data-driven intensity thresholding procedure. Starting from an empirical threshold, estimated via kernel density estimation of voxel intensities, the algorithm progressively lowered the threshold and identified voxels that exceeded this intensity threshold until a connected component exceeding the expected physical volume of the electrode was found. Based on manufacturer specifications, the expected physical volume was a cylinder with a 1.28-mm diameter and a 1.57-mm length. We assigned the centroid of this component as the refined electrode location and validated the refined locations using Freeview.

Next, we inferred the positions of the microelectrodes from the geometry of the macro contacts. We fitted a line to each trajectory based on the macroelectrode positions using singular value decomposition. The initial estimated micro contact was located 3 mm beyond the macro contact, along the same shank axis, most distal to the entry point. The initial estimate of 3 mm was provided by the surgeon, who trimmed the micro-wires during implantation. To determine the actual microelectrode position, we localized the most intense voxel cluster in the CT volume near the initial microwire estimation, using the same intensity-guided connected-component algorithm.

To further verify the accuracy of electrode localization and to avoid any confusion in the labeling electrodes, we exported the planned electrode trajectories from the Robotic Operating Surgical Assistant (ROSA) platform, where trajectories were defined in the coordinate space of the intraoperative imaging system. To transfer these trajectories into the same anatomical reference space as the localized electrode contacts, we performed rigid-body image coregistration between the ROSA display image and the pre-operative T1-weighted MRI. The transformation was then applied to the extracted ROSA trajectories. As a result, both the planned lead trajectories and the electrode localizations were in the same right-anterior-superior (RAS) coordinate space defined by the pre-operative MRI.

#### Eyeball-to-Electrode Distance calculation

To determine whether the observed SREP could be attributed to oculomotor signals, we performed regression analysis between the eyeball-to-electrode distance and SREP characteristics such as peak amplitude. We used the Euclidean distance from each electrode contact to the nearest eye-globe center to approximate the distance between electrode contacts and the myogenic dipole source generated by extraocular muscle activation. We identified the center of each eyeball for each subject based on their pre-operative T1-weighted MRI scans and then computed the 3-dimensional (3D) Euclidean distance from the left and right eyeballs' centers to each electrode contact's coordinates. We used the minimum of the left and right eyeball distances, representing the distance from an electrode to the closest eyeball, as the eyeball-to-electrode distance for subsequent analyses.

#### Calculation of saccade eccentricity, saccade direction, and saccade and fixation duration

We calculated saccade eccentricity, saccade direction, and saccade and fixation duration based on the onset and offset time of the detected saccade or fixation. We transformed gaze coordinates into a vector representing gaze coordinates relative to eye location in the right-anterior-superior (RAS) coordinate system, and normalized it to unit length. Based on the normalized vector $\overrightarrow{(gaze_x, gaze_y, gaze_z)}$, we calculated the azimuth and elevation angle ($\theta_{azimuth}$ and $\theta_{elevation}$) in polar coordinates. In this study, we defined horizontal eye movements as saccades with azimuth angles within $\pm 20°$ of the horizontal axis, and vertical eye movements as saccades with elevation angles within $\pm 20°$ of the vertical axis.

$$\theta_{azimuth} = tan^{-1}(\frac{gaze_y}{gaze_x})$$

$$\theta_{elevation} = tan^{-1}(\frac{gaze_z}{\sqrt{gaze_x^2 + gaze_y^2}})$$

We calculated the angular change $\Delta\theta_{azimuth}$ and $\Delta\theta_{elevation}$ based on $\theta_{azimuth}$ and $\theta_{elevation}$ at saccade onsets and offsets. We defined the saccade eccentricity as $\sqrt{\Delta\theta_{azimuth} + \Delta\theta_{elevation}}$. We defined saccade direction $\theta_{dir}$ as the orientation of the 2D angular displacement vector connecting gaze positions before and after each saccade. In this convention, 0 $rad$ (e.g., 0°) corresponds to rightward saccades, $\frac{\pi}{2}$ $rad$ (e.g., 90°) to upward saccades, $\pi$ $rad$ (e.g., 180°) to leftward saccades, and $\frac{3\pi}{2}$ $rad$ (e.g., 270°) to downward saccades.

$$\theta_{dir} = tan^{-1}(\frac{\Delta\theta_{elevation}}{\Delta\theta_{azimuth}})$$

To study whether SREPs encode the direction of the next saccades, we identified consecutive saccades that occurred with both spatial and temporal continuity constraints. Specifically, we required the spatial distance between the end of the current saccade and the start of the subsequent saccade to be less than 20% of the display dimension, and the inter-saccadic interval to be shorter than 550 ms.

#### Quantification of SREP consistency and regional SREP prevalence

To identify the brain region where LFP waveforms in the peri-saccadic window were most consistent, we computed the SREP prevalence for each brain structure. The SREP prevalence was defined as the proportion of electrode contacts whose peri-saccadic AUC exceeded the mean + 2 SDs of their respective baseline AUC values. The use of proportion rather than the number of electrode contacts controls for varying sampling density across the cerebrum. We calculated the SREP prevalence for the following structures: White Matter (Left, Right), Hippocampus (Left, Right), Amygdala (Left, Right), Basal Ganglia, Thalamus, Orbitofrontal (Left, Right), Inferior Frontal (Left, Right), Middle Frontal (Left, Right), Superior Frontal (Left, Right), Anterior Cingulate, Posterior Cingulate, Motor (precentral gyrus and paracentral lobule, Left, Right), Anterior Temporal (i.e., temporal pole, Left, Right), Middle Temporal (Left, Right), Superior Temporal (Left, Right), Inferior Temporal (Left, Right), Medial Temporal (Left, Right), Fusiform (Left, Right), Parietal (Left, Right), Occipital (Left, Right), and Insula (Left, Right).

To robustly quantify the fraction of electrode contacts within a region that exhibit consistent saccade-related activity, we systematically varied the responsiveness threshold from mean + 1 SD to mean + 4 SD of baseline AUC. For each threshold, we calculated the proportion of electrode contacts exceeding the threshold. This yielded a curve where the y-axis represented the SREP prevalence defined before, and the x-axis represented the responsiveness threshold. As an integrated measure of regional LFP waveform consistency following saccades, we computed the area under this curve (AUC) across all thresholds. To control for the number of sampling electrodes across brain structures, we resampled electrodes with replacement to equalize sample sizes across regions, then repeated the AUC calculation. We computed the Pearson correlation coefficient between AUC with and without resampling, and demonstrated the stability of the AUC value with respect to regional sampling density.

To determine the spatial distribution of the degree of LFP waveforms consistency following saccades, we computed the difference in AUC between peri-saccadic and baseline inter-trial correlation distributions (**Fig. S4C**). As a complementary measure, we defined the degree of consistency as the average AUC of the probability density over inter-trial correlation following saccades, computed separately for the two clusters and then averaged (**Fig. S4D**).

#### Quantifying intra- vs inter-contact variability of SREP characteristics

We observe trial-by-trial variations in recognition memory. Thus, if SREPs contribute to encoding, within-electrode-contact SREP characteristics should vary across saccades and trials. if SREPs reflect a spatially organized neural process, we also expect across-electrode contact variability in SREP characteristics such as peak amplitude and onset latency. It is our intention to quantify intra-contact versus inter-contact (spatial) variability in SREP latency and peak amplitude to determine whether SREPs were anatomically organized. To ensure comparability, we estimated intra-contact and inter-contact variability based on matched sample sizes.

To compute intra-contact variability, we first resampled SREP characteristics, including latency and peak amplitude, within each electrode contact to the median saccade count (i.e., 144) across contacts, thereby minimizing the effect of sample size on variability estimates and ensuring comparability across contacts. We then computed the standard deviation (SD) of SREP latency and peak amplitude for each contact. To estimate inter-contact (e.g., spatial) variability with matched sampling, we resampled electrode contacts to equalize the sample number used for intra-contact SD estimation. We calculated the SD of latency and peak amplitude from the averaged SREP waveform. We repeated this procedure 100 times and averaged the SD of latency for statistical comparison.

To assess whether polarities affected SREP latency or peak amplitude distribution, at each contact, we computed SREP latency from the cluster-level waveform for each polarity and assessed Pearson correlations between paired latencies or peak amplitudes across contacts. To further validate that the SREP characteristics differences, including latency and peak amplitude differences, between contacts capture the spatial organization of saccadic modulation, we related the between-contact SREP characteristic differences derived from trial-averaged SREPs to the effect size of latency differences estimated from single-trial SREPs. Because the number of contact pairs is large, we randomly sampled 2,000 contact pairs per polarity and computed the Pearson correlation.

#### Spatial visualization of SREP characteristics

To illustrate the spatial organization of SREPs at both the brain-region and electrode-contact levels, we developed a surface-based visualization framework that maps outcome values onto a brain model, allowing us to convey findings related to the spatial organization intuitively. Using this framework, we visualized SREP characteristics, such as latency and amplitude (**Figure 4B–C**), and other outcomes, such as the proportion of electrode contact showing early SREP (**Figure 4D**) and feature importance in predicting recognition memory (detailed in MATERIALS AND METHODS: **Model Evaluation**). For region-level mapping, we assigned each electrode contact an anatomical label using the majority-vote volumetric approach, detailed in the MATERIALS AND METHODS: **Anatomical localization of the electrode** section. We computed the average outcome value, such as the mean SREP latency, across electrode contacts within each region. For each brain region, we reconstructed and rendered three-dimensional surfaces from a reference MRI volume using the PyVista Python package. We mapped regional average values onto the corresponding cortical or subcortical surface meshes and color-coded them using a sequential colormap (e.g., from pale to dark green).

To provide higher-resolution spatial information, we constructed 2D glass-brain heatmaps projected from the 3D brain model. Briefly, we projected electrode coordinates in the common anatomical space onto a 2D plane orthogonal to a vector representing the viewing angle. Given the electrode size and approximately 1-cm reach of signals. We discretized this 2D space into grid elements, each as a 5 mm × 5 mm square, unless specified otherwise. For each grid element, we computed the local mean outcome values, for example, mean EP latency, across nearby contacts. For each grid cell, we defined nearby contacts as those located within a 10-mm radius. We color-coded each grid element based on the local mean outcome values using a white-to-red colormap. Grid elements without nearby electrode contacts were rendered transparent to truly represent electrode coverage.

#### Assessing the reproducibility of learned mappings between SRND and visual encoding outcome

To verify that RF models learned the underlying mapping between SRND parameters and recognition memory performance, we quantified the reproducibility of learned mappings across cross-validations (CVs). We first analyzed the consistency of feature importance across folds. For each fold, the importance of a feature $f_j$ was the mean reduction in Gini index across all decision nodes where the feature was used:

$$FI(f_j) = \frac{1}{T}\sum_{t=1}^{T}\sum_{n\in N_{j,t}}\frac{N_n}{N_t}\,\Delta G(n),$$

where $T$ is the number of trees, $N_{j,t}$ is the set of nodes in tree $t$ that split on feature $f_j$, $N_t$ is the total number of samples used to train tree $t$, and $N_n$ is the number of samples reaching node n, thus, $\frac{N_n}{N_t}$ represents the proportion of samples affected by the split at node $n$. $\Delta G(n)$ denotes the reduction in Gini impurity index at node $n$ before and after the split:

$$\Delta G(n) = Gini\ index_{\text{parent}}(n) - \left(\frac{N_L}{N_n}Gini\ index_L + \frac{N_R}{N_n}Gini\ index_R\right),$$

where $Gini\ index_{\text{parent}}(n)$ is the Gini impurity index of the parent node $n$ before splitting. $Gini\ index_L$, and $ini\ index_R$ denote the Gini impurity index of the left, and right children's node, respectively, where $N_L$ and $N_R$ denote their respective sample counts, and $N_n = N_L + N_R$.

To complement Gini index-based feature importance measures, we also evaluated the reproducibility of Shapley value–derived feature contributions. Originating from cooperative game theory, the Shapley value for a given sample and a feature represents the feature's marginal contribution to the model output(*36*). For instance, for a sample $x$ with model output $f(x)$, the Shapley value $Shapley_j(x)$ for a feature $j$ is the average change in the prediction when adding the feature $j$ to all possible subsets of the other features. (see **Shapley additive explanation** section for detailed computation). For each CV iteration, we computed the mean absolute Shapley value of each feature across training samples, which represents a model-agnostic quantification of each feature's marginal impact on the model output.

We normalized feature importances using the *norm* function from *Numpy.linalg* Python package and ranked them within each CV iteration. We quantified the concordance of feature rankings across folds, for both Gini index-based and Shapley-value derived feature importance, using Kendall's coefficient of concordance ($W$). Kendall's coefficient of concordance is a rank-based measure of agreement that ranges from 0 (no concordance) to 1 (perfect concordance).

$$W = \frac{12S}{m^2(n^3 - n)}$$

$$S = \sum_{j=1}^{n} \left(R_j - \bar{R}\right)^2$$

$m$ is the number of folds, $n$ is the number of features, $R_j$ is the sum of ranks assigned to feature $j$ across folds, and $\bar{R} = n(m + 1)/2$ is the expected rank sum under uniform ranking.

To evaluate the statistical significance of the observed concordance, we generated a null distribution of $W$ values derived from RF models trained on data with permuted class labels. For each permutation, we performed a 5-fold CV and obtained a rank vector of feature importance for the five trained RF models. We then computed $W$ values for 1,000 randomly sampled sets of five permuted-label models, yielding an empirical null distribution for comparison against the observed $W$.

In addition, we verified that trained RF models across CVs learned similar decision rules. Each decision rule corresponds to a split threshold on a specific feature that maximizes the reduction in impurity. We reasoned that if independently trained models converged on the same underlying mapping between SRND parameters and memory outcomes, the distributions of these split thresholds should be consistent across folds. For each RF model and each feature, we extracted split thresholds across all nodes and trees. To enable comparability across models, we established a set of reference bins for each feature. Specifically, we computed 100 quantile-based bin edges using the empirical distribution of split thresholds from a reference model (i.e., the model from the first fold). To ensure strictly increasing bin boundaries, we added a small constant ($10^{-8}$) to the second of any consecutive quantiles with equal values. We then ordered features along the x-axis and constructed a composite split histogram for each RF model, where each bin index represented a unique combination of feature identity and threshold range. In this representation, each histogram summarizes how frequently each feature contributed to splits within specific threshold intervals for a given fold.

To ensure comparability across folds, we normalized each composite split histogram by its total number of splits so that the resulting density distribution summed to one. We quantified the similarity of decision rules between pairs of RF models by computing the area under the absolute difference curve (AUC) between their corresponding kernel density estimates. We obtained kernel density estimates using the *gaussian_kde* function from the *scipy.stat Python* package, with a bandwidth parameter of 0.01.

To assess the statistical significance of the observed decision-rule similarity between pairs of RF models, we compared observed similarities from the actual data to a generated null distribution. Specifically, we permuted the class labels 20 times and applied 5-fold cross-validation to each permutation, yielding 100 RF models trained under random label conditions. From this pool, we randomly selected pairs of RF models and computed the decision rule similarity using the same procedure as for the actual data.

To further demonstrate that RF models convergently learned the mapping between SRND parameters and visual encoding outcome, we examined the consistency of predicted probabilities and classes across models trained on different CV iterations. For each pair of models trained on actual data but in different folds, we input 200 testing samples from the shared test set, which are observations that were not included in the training data of either model. We then computed Pearson's correlation coefficient between the predicted probabilities of "remembered" for the input testing samples using the *pearsonr* function from the *scipy.stats* Python package. For the same model pairs, we also computed the disagreement ratio as the proportion of test samples for which the predicted classes differed, defined as disagreement ratio. This disagreement ratio captures divergence in binary decisions rather than probabilistic calibration. To assess the statistical significance of the observed output correlation and disagreement ratio across CV iterations, we generated their corresponding null distributions by randomly pairing RF models trained on permuted labels 200 times and recalculating these two output (dis)agreement metrics.

#### Shapley additive explanation

For a given sample $x_i$ and feature $j$, the Shapley value $Shapley_{i,j}$ quantifies the *expected* marginal contribution of feature $j$ to the model's predicted probability of successful visual encoding. In other words, Shapley values quantify how much a given feature value increases or decreases the probability of successful memory encoding for individual observations, while accounting for correlations among features. The marginal contribution of a feature is defined as the *expected* change in the model's prediction when that feature is included compared with when it is excluded, given a specific subset of other features $S \subseteq F \setminus \{j\}$. Here, $F$ is the full feature set, and $S$ is a subset of $F$ excluding feature $j$. Operationally, we computed the Shapley value by averaging marginal contributions across all possible subsets $S$, weighted by the probability of each subset that would occur if we randomly ordered all features. Specifically, a subset $S$ of size $|S|$ would occur with probability $\frac{|S|!(|F|-|S|-1)!}{|F|!}$, which is the number of ways features could be ordered such that the features in $S$ appear before feature $j$, and the remaining features in $F \setminus S$ appear after $j$. For example, if $F = (j_1, j_2, j_3, j)$, and $S = \{j_1, j_2\}$, there are $|S|! \times (|F| - |S| - 1)! = 2! \times 1! = 2$ permutations where features in $S$ appear before feature $j$: $(j_1, j_2, j, j_3)$and $(j_2, j_1, j, j_3)$. Mathematically, Shapley value is defined as:

$$\text{Shapley}_{i,j} = \sum_{S \subseteq F \setminus \{j\}} \frac{|S|!\,(|F| - |S| - 1)!}{|F|!} \left[f_{S\cup\{j\}}\left(x_{i,S\cup\{j\}}\right) - f_S\left(x_{i,S}\right)\right]$$

where |S| is the number of features in subset S, and |F| is the total number of features. The term $x_{i,S}$ denotes the subset of feature values from sample $x_i$ corresponding to the feature set $S \subseteq F \setminus \{j\}$, while $x_{i,S\cup\{j\}}$ denotes the same sample but with feature $j$ additionally included. $f_S\left(x_{i,S}\right)$ represents the expected model prediction when only features in $S$ are known.

$$\begin{aligned} f_S\left(x_{i,S}\right) &= E_{x_{F\setminus S}}\left[f\left(\left[x_{i,S}, x_{i,F\setminus S}\right]\right)\right] \\ &= \int f\left(\left[x_{i,S}, x_{i,F\setminus S}\right]\right) p\left(x_{F\setminus S} \middle| x_{i,S}\right) dx_{F\setminus S}\,, \end{aligned}$$

where $x_{F\setminus S}$ represents all the other feature values that are not in $S$, $x_{i,S}$ is the subset of feature values for sample $i$ corresponding to features in $S$. Therefore, $f\left(\left[x_{i,S}, x_{i,F\setminus S}\right]\right)$ is the model output from a complete feature vector where the features in subset $S$ take the actual values from sample $i$ (i.e., $x_{i,S}$), and the remaining features in $F \setminus S$ take some possible values drawn from the distribution$\sim p\left(x_{F\setminus S} \middle| x_{i,S}\right)$. Thus, $f_{S\cup\{j\}}\left(x_{i,S\cup\{j\}}\right) - f_S\left(x_{i,S}\right)$ represents the marginal contribution of feature $j$ to the model's prediction for

$x_i$. The weighting term $\frac{|S|!(|F|-|S|-1)!}{|F|!}$ guarantees additivity, meaning that each sample's prediction can be exactly decomposed into the sum of feature contributions. By summing the Shapley value for a sample $x_i$ across all features, we get:

$$\sum_{j\in F} Shapley_{i,j} = \sum_{j\in F}\sum_{S\subseteq F\setminus j} \frac{|S|!\,(|F|-|S|-1)!}{|F|!}\left[f_{S\cup j}(x_{i,S\cup j}) - f_S(x_{i,S})\right]$$
$$= f(x_i) - f_\emptyset \;=\; f(x_i) - E[f(x)]$$

In tree-based models, $f_S(x_{i,S})$ can be expressed as the expectation of the model's output conditioned on $x_S$.

$$f_S(x_{i,S}) = \sum_{L\in\text{leaves}} P(L|x_{i,S})\, v_L,$$

where $x_{i,S}$ denotes the subset of feature values from sample $x_i$ corresponding to the feature set $S \subseteq F$, $v_L$ denotes the predicted value (e.g., predicted probability of successful visual encoding) at leaf $L$. $P(L|x_{i,S})$ represents the probability of reaching leaf $L$ given only the observed features $x_{i,S}$.

For a given leaf $L$, we computed this probability $P(L|x_{i,S})$ using the chain rule of probability applied along the path from the root node to $L$, that is, the product of conditional probabilities of following each successive branch along this path. For node $n$ splits on a feature in $S$, the observed feature value $x_{i,S}$ deterministically specifies which branch the sample follows. The probability of taking the branch toward $L$ is 1 if $x_{i,S}$ satisfies the split condition for that branch, and 0 otherwise. If node $n$ splits on a feature not in $S$, the conditional probability of following the branch toward $L$ is empirical frequency $r_{n\to L}/r_n$ from the training data, where $r_n$ is the number of training samples reaching node $n$, and $r_{n\to L}$ is the number that proceed down the branch toward $L$. In other words, given training data, the tree defines a discrete probability distribution over leaves $P(L \mid x_{i,S})$, the likelihood that a random training sample reaches leaf $L$ when only features in $S$ are known.

$$P(L \mid x_{i,S}) = \prod_{n\in path(L)} \begin{cases} 1, & split\ feature(n) \in S\ AND\ x_{i,S}\ directs\ x_i\ toward\ L \\ 0, & split\ feature(n) \in S\ AND\ x_{i,S}\ down\ a\ different\ branch \\ r_{n\to L}/r_n, & split\ feature(n) \notin S \end{cases}$$

To illustrate, consider a simple decision tree in which the root node (Node 0) splits on feature A with a threshold of 5. The left branch ($A < 5$) leads directly to a single leaf $L_1$ with an output value $v_{L_1}$= 0.8, while the right branch ($A \geq 5$) leads to another leaf $L_2$ with $v_{L_2}$ = 0.2. 60 % of the training sample falls on the left side of the split ($L_1$) and 40 % on the right ($L_2$). These proportions define the empirical branch probabilities $r_{\text{left}}/r_{\text{root}} = 0.6$ and $r_{\text{right}}/r_{\text{root}} = 0.4$. The table below summarizes the tree structure.

| Node | Split | Left | Right | Training proportion |
|---|---|---|---|---|
| 0 | Left if $A < 5$ | $L_1(v_{L_1} = 0.8)$ | $L_2(v_{L_2} = 0.8)$ | $r_{0\to L_1}/r_0 = 0.6$<br>$r_{0\to L_2}/r_0 = 0.4$ |

To calculate $Shapley_{i,A}$ for $x_{i,A} = 3$, we have

$$Shapley_{i,A} = \sum_{S\subseteq F\setminus\{A\}} \frac{|S|!\,(|F|-|S|-1)!}{|F|!}\left[f_{S\cup\{A\}}(x_{i,S\cup\{A\}}) - f_S(x_{i,S})\right].$$

As $F = A$, $S$ can only be $\emptyset$. *Note that* $|\emptyset| = 0$, *therefore*

$$Shapley_{i,A} = \frac{|A|!\ (|A| - |\emptyset| - 1)!}{|A|!}\left[f_A(x_{i,A}) - f_\emptyset\right]$$

$$= f_A(x_{i,A}) - f_\emptyset$$

$$f_\emptyset = \sum_{L \in leaves} P(L|x_{i,\emptyset})v_L$$

$$= r_{0\to L_1}/r_0 \cdot v_{L_1} + r_{0\to L_2}/r_0 \cdot v_{L_2}$$

$$= 0.6 \times 0.8 + 0.4 \times 0.2 = 0.56$$

$$f_A(x_{i,A}) = \sum_{L \in leaves} P(L|x_{i,A})v_L$$

$$= r_{0\to L_1}/r_0 \cdot v_{L_1} + r_{0\to L_2}/r_0 \cdot v_{L_2}$$

$$= 1 \times 0.8 + 0 \times 0.2 = 0.8$$

Now consider a deeper decision tree in which the root node (Node 0) splits on feature A at a threshold of 5. Samples with $A < 5$ proceed to the left child node (Node 1), whereas those with $A \geq 5$ terminate at a right leaf $L_3$ ($v_{L_3}$= 0.4). At Node 1, the data are further partitioned based on feature $B$ with a threshold of 2: samples with $B < 2$ reach leaf $L_1$ with $v_{L_1}$=0.6, and samples with $B \geq 2$ reach leaf $L_2$ with $v_{L_2}$= 0.5. At the root node, the left and right branch proportions are $r_{0\to 1}/r_0 = 0.6$ and $r_{0\to L_3}/r_0 = 0.4$ respectively. At Node 1, the split on feature B partitions the data evenly, with $r_{1\to L_1}/r_1 = 0.5$ and $r_{1\to L_2}/r_1 = 0.5$. The table below summarizes the tree structure.

| Node | Split | Left | Right | Training proportion |
|---|---|---|---|---|
| 0 | Left if $A < 5$ | $Node\ 1$ | $L_3(v_{L_3} = 0.4)$ | $r_{0\to 1}/r_0 = 0.6$<br>$r_{0\to L_3}/r_0 = 0.4$ |
| 1 | Left if $B < 2$ | $L_1(v_{L_1} = 0.6)$ | $L_2(v_{L_2} = 0.5)$ | $r_{1\to L_1}/r_1 = 0.5$<br>$r_{1\to L_2}/r_1 = 0.5$ |

We illustrate how to calculate $Shapley_{i,A}$ with $x_{i,\{A,B\}} = [1,1]$. As $F = \{A, B\}$, S can be $\emptyset$ or $\{B\}$.

$$Shapley_{i,A} = \sum_{S \subseteq F \setminus \{A\}} \frac{|S|!\ (|F| - |S| - 1)!}{|F|!}\left[f_{S\cup\{A\}}(x_{i,S\cup\{A\}}) - f_S(x_{i,S})\right]$$

$$= \frac{1}{2}\left[f_A(x_{i,A}) - f_\emptyset\right] + \frac{1}{2}\left[f_{\{A,B\}}(x_{i,\{A,B\}}) - f_B(x_{i,B})\right]$$

Similarly, we have

$$f_\emptyset = \sum_{L \in leaves} P(L|x_{i,\emptyset})v_L$$

$$= P(L_1|x_{i,\emptyset})v_{L_1} + P(L_2|x_{i,\emptyset})v_{L_2} + P(L_3|x_{i,\emptyset})v_{L_3}$$
$$= r_{0\to1}/r_0 \cdot r_{1\to L_1}/r_0 \cdot v_{L_1} + r_{0\to1}/r_0 \cdot r_{1\to L_2}/r_0 \cdot v_{L_2} + r_{0\to L_3}/r_0 \cdot v_{L_3}$$
$$= 0.6 \times 0.5 \times 0.6 + 0.6 \times 0.5 \times 0.5 + 0.4 \times 0.4 = 0.49$$

$$f_A(x_{i,A}) = \sum_{L \in leaves} P(L|x_{i,A})v_L$$
$$= P(L_1|x_{i,A})v_{L_1} + P(L_2|x_{i,A})v_{L_2} + P(L_3|x_{i,A})v_{L_3}$$
$$= 0.5 \times 0.6 + 0.5 \times 0.5 + 0 \times 0.4 = 0.55$$

$$f_B(x_{i,B}) = \sum_{L \in leaves} P(L|x_{i,B})v_L$$
$$= P(L_1|x_{i,B})v_{L_1} + P(L_2|x_{i,B})v_{L_2} + P(L_3|x_{i,B})v_{L_3}$$
$$= 0 \times 0.6 + 0 \times 0.5 + 1 \times 0.4 = 0.4$$

$$f_{\{A,B\}}(x_{i,\{A,B\}}) = \sum_{L \in leaves} P(L|x_{i,\{A,B\}})v_L$$
$$= P(L_1|x_{i,\{A,B\}})v_{L_1} + P(L_2|x_{i,\{A,B\}})v_{L_2} + P(L_3|x_{i,\{A,B\}})v_{L_3}$$
$$= 1 \times 0.6 + 0 \times 0.5 + 0 \times 0.4 = 0.6$$

Therefore, $Shapley_{i,A} = \frac{1}{2}[0.55 - 0.49] + \frac{1}{2}[0.6 - 0.4] = 0.13$

Similarly, we have

$$Shapley_{i,A} = \frac{1}{2}\left[f_B(x_{i,B}) - f_\emptyset\right] + \frac{1}{2}\left[f_{\{A,B\}}(x_{i,\{A,B\}}) - f_A(x_{i,A})\right]$$
$$= \frac{1}{2}[0.4 - 0.49] + \frac{1}{2}[0.6 - 0.55] = -0.02$$

We have $Shapley_{i,A} + Shapley_{i,B} = f(x_i) - E[f(x)] = 0.6 - 0.49 = 0.11$.

We implemented this analytical solution using *TreeExplainer* function from the *SHAP* Python package. For each input sample, this function recursively traverses all decision paths through each tree, computing $P(L|x_{i,S})$ for each feature subset $S$. Given that our previous analyses demonstrated high reproducibility of learned mapping across CV iterations, and that Shapley value computation is computationally intensive, we computed Shapley values for the trained RF model in a randomly selected CV iteration. This analysis allows us to identify feature values that increase or decrease the model's predicted probability of successful encoding, thereby translating the RF classifier into interpretable electrophysiological rules.

To further illustrate the meaning of Shapley values, we compared the feature value-Shapley value plots with partial dependence plots (**Figure 6B**). **Figure 6A** shows, for a representative electrode contact in the superior temporal gyrus, how Shapley values varied as a function of each SRND parameter's observed value. A partial dependence plot provides a global, approximate summary of how the model's output changes as that feature varies while averaging over the effects of all other features. In contrast, Shapley values capture the local (e.g., specific to an anatomical location), sample-specific contribution of each feature by considering all possible feature interactions. To visualize how multiple features jointly influenced model prediction of successful visual encoding, we systematically varied SRND parameters for each electrode contact. We input the feature combination into the trained RF model to estimate the probability of successful encoding. We then reconstructed waveforms from the SRND parameters and assigned the predicted encoding probability to each waveform (**Figure 6C**).

#### Visualization of the SRND-visual encoding relationship

After confirming that the Shapley value-informed thresholds and directions distinguished successful from unsuccessful visual encoding outcomes, we visualized them to illustrate how SRND features contributed to successful visual encoding across anatomical regions (**Figure 6I–M**). For non-circular features, we used two complementary color schemes to represent the Shapley-informed direction, that is, whether higher or lower feature values favored successful encoding. A red colormap (light to dark red) indicated that higher feature values were associated with successful encoding, whereas a blue colormap (light to dark blue) indicated that lower feature values were associated with successful encoding. We used color saturation to reflect the magnitude of the threshold, with more saturated hues corresponding to more extreme thresholds, in other words, higher thresholds for "higher" directions and lower thresholds for "lower" directions.

For circular features (i.e., the instantaneous phase at 0.5 OCs before EP latency), we computed the circular mean of the encoding-favorable phase window $\left[\theta_{1_f}, \theta_{2_f}\right]$ when the direction was "within," or its complementary phase range $\neg\left[\theta_{1_f}, \theta_{2_f}\right]$ when the direction was "outside." We color-coded the circular mean of the encoding-favorable phase using the *twilight* circular colormap from the *Matplotlib* Python Package.

To visualize the SRND-visual encoding relationship at electrode contact resolution, we projected the 3D cortical and subcortical surfaces, along with electrode coordinates, onto a 2D plane and discretized this space into a 5 mm × 5 mm grid, as detailed in the **Spatial visualization of SREP characteristics** section. We assigned each electrode contact's mean absolute Shapley value or Shapley value-informed threshold to grid cells within 10 mm. Grid cells with no assigned values were rendered transparent. The resulting heatmaps depict the regional gradient of model contributions, with color intensity encoding the mean absolute Shapley values. Background anatomical surfaces were overlaid for spatial reference and color-coded by anatomical region without encoding Shapley values.

#### RF regression for predicting saccade direction

To test whether SRND parameters contain information about saccade direction, we split the training and testing datasets at the saccade level and trained RF regressors. Similar to the random forest model for classifying remembered vs. forgotten trials, the random forest regressor is an ensemble of decision trees, with each tree recursively splitting the data based on feature thresholds. During training, the random forest regressor learned a set of features and split thresholds that minimize mean square error within each split node. After training, the output at each leaf node was the average target values across the training samples that reached that node.

The input feature set was identical to that used for visual encoding outcome prediction parameters (i.e., SREP latency, peak-to-trough amplitude, peak time, phase, and power of preceding oscillatory activity). Training RF regressors largely followed the same framework described in the **Training RF classifiers** section, including adding Gaussian spatial jitter to electrode contact location and using 5-fold cross-validation. In addition, we verified that the RF regression model was neither overfitting nor underfitting by systematically varying key hyperparameters (**Fig. S16A**). The differences, compared to training an RF classifier to learn the SRND-visual encoding mapping, were the target variable, objective function, and model output. Because saccade direction is a circular variable, its numerical representation in radians is discontinuous at the $\{0, 2\pi\}$ boundary. For instance, $0$ rad and $2\pi$ rad correspond to the same direction but differ numerically by $2\pi$. To allow the RF regression model to optimize the loss function in Euclidean space and avoid treating saccade direction as a linear quantity, we transformed each saccade's direction $\theta$ into a two-dimensional direction vector in Cartesian space:

$$y_i = [\sin(\theta_i),\ \cos(\theta_i)].$$

This maps angular saccade directions to continuous coordinates in Euclidean space, where opposite directions remain far apart in vector space, while equivalent directions are identical, preserving angular continuity. After transformation, we trained RF regression models using a standard mean squared error (MSE) objective function. We implement the RF regression model using *RandomForestRegressor* from the *Scikit-learn* Python package. In brief, each decision tree learned feature-split thresholds that minimized MSE over the two-dimensional direction vectors. For each sample, the trained RF model routes it to a terminal leaf in every individual decision tree. The predicted output for that sample from a single tree is the mean direction vector computed across all training samples that reached the same terminal leaf. The final ensemble prediction is the average predicted direction vector across all trees $\widehat{y_i}$. We reconstructed the predicted saccade direction $\widehat{\theta}_i$ for sample $i$ in angular space using the two-argument arctangent function:

$$\hat{\theta}_{\mathrm{i}} = \mathrm{atan2}(\widehat{y_{i}}_1, \widehat{y_{i}}_2).$$

#### RF regression evaluation

We evaluated RF regression model performance on held-out testing data using both MSE and mean absolute error (MAE). Specifically, we calculated the MSE between the predicted and true direction vectors.

$$\mathrm{MSE} = \frac{1}{N}\sum_{i=1}^{N} ||\widehat{y_i} - y_i||^2,$$

where $||\cdot||$ denotes the Euclidean norm. Lower MSE values indicate better prediction accuracy. In addition, we computed the MAE between the reconstructed angular predictions and actual saccade directions

$$\mathrm{MAE} = \frac{1}{N}\sum_{i=1}^{N} |\widehat{\theta}_i - \theta_i|,$$

where angular differences $\widehat{\theta_i} - \theta_i$ were wrapped into the range $[-\pi, \pi]$ before taking the absolute value. We computed the MSE and MAE at both the location and saccade levels. For saccade-level evaluation, we averaged the predicted direction vectors across all electrodes within each saccade. To assess statistical significance, we generated null distributions for MSE and MAE by repeating the cross-validation training procedure with permuted saccade directions. Specifically, for each of 100 permutations, we randomly shuffled target direction vectors $y_i$, repeated the RF regression model training procedure, and computed the performance metrics.

#### Quantifying directional modulation

To confirm that SRND parameters did not encode saccade direction for most electrode contacts, we compared the observed testing MSE for each electrode contact against its null distribution from permutations. We calculated the contact-wise percentile rank of testing MSE relative to its null distribution. We identified direction-informative electrode contacts as those with observed MSEs below the 5th percentile of their null distribution. To complement the model-based approach, we quantified directional non-uniformity of SRND parameters. For each electrode contact and feature, we estimated the circular kernel density of the feature values as a function of saccade direction. Operationally, we discretized the angular space into 60 equal bins covering $0 - 2\pi$ radians. For each bin centered at angle $\phi_j$, we computed the mean feature value of saccades within a $\pm 6°$ window around $\phi_j$. For circular features (i.e., instantaneous phase), we computed the circular mean using the *circmean* function from the *scipy.stats* Python package. We then applied von Mises kernel regression, implemented via *vonmises* from the *scipy.stats python package* (concentration parameter κ = 15).

For each feature, we quantified directional non-uniformity $D_f$ as the integral of the absolute difference between the circular kernel density and its mean across the angular domain:

$$D_f = \int_0^{2\pi} \left| p_f(\theta) - \overline{p_f} \right| \, d\theta,$$

where $p_f(\theta)$ denotes the kernel density estimate for feature $f$, and $\overline{p_f}$ is its circular mean. A higher $D_f$ value indicates greater directional modulation.

To evaluate statistical significance, we generated null distributions of $D_f$ by permuting saccade directions 200 times and recomputing the directional non-uniformity for each permutation. We then computed the percentile rank of directional non-uniformity for each electrode contact and feature relative to the corresponding null distribution. For each feature, we computed the proportion of electrode contacts whose directional non-uniformity is less than the 95th percentile of their relative null distribution, indicating a lack of direction-specific modulation.

#### Neural networks for identifying saccade direction information in SRND

To validate the observed lack of saccade-direction information in SRND parameters, we implemented several complementary modeling approaches. These analyses aim to demonstrate that the lack of saccade-direction encoding across nonlinear, linear, and binary classification frameworks, and therefore confirm that SRND primarily reflects cognitive rather than motoric components of saccade processing. Specifically, we trained (1) neural networks with either saccade direction or transformed direction vector as output, (2) a linear model for angular prediction, and (3) a binary RF classifier to distinguish leftward versus rightward saccades. These analyses aimed to detect any nonlinear or linear mapping between SRND parameters and saccade direction.

As a nonlinear control model, we implemented a feedforward neural network consisting of three fully connected hidden layers, each followed by a Rectified Linear Unit activation function using the *PyTorch* Python package. The input layer received the same feature set as the RF models, including electrode spatial coordinates and SRND parameters, described in the **Training and testing data structure** section. The network's output layer contained two units representing the two-dimensional Cartesian saccade direction vector $[\sin(\theta), \cos(\theta)]$. To ensure that predictions represented saccade directions, we required the network to normalize its outputs to the unit circle:

$$\hat{y}_i = \frac{u_i}{|u_i|}, \qquad u_i = f_\theta(x_i),$$

where $f_\theta(x_i)$ denotes the network output for input $x_i$. We trained this neural network to minimize MSE between predicted and true direction vectors. Consistent with previous machine learning model training and evaluation, we used the 5-fold cross-validation procedure. We trained the network using the Adam optimizer from the *PyTorch* Python package with a learning rate of $1 \times 10^{-3}$, a batch size of 256, and up to 50 epochs.

To confirm that the transformation of saccade direction into a two-dimensional Cartesian direction vector did not prevent the neural network from learning the relationship between SRND parameters and saccade direction, we implemented a complementary neural network trained to minimize angular loss directly. This model had the same three-layer architecture and inputs as above, but used a single-unit output layer representing the angular direction in radians. We used a loss function to represent the wrapped angular distance between the predicted direction ($\hat{\theta}$) and true directions ($\theta$):

$$\mathcal{L}_{angular} = 1 - cos(\hat{\theta} - \theta).$$

This formulation penalizes large angular deviations and is continuous at the $0 - 2\pi$ boundary. We evaluated both neural networks at the electrode contact and saccade levels using MSE, computed analogously to the RF regression evaluation (see the **RF regression evaluation** section). For the network with two outputs, we reconstructed the predicted angular direction from the predicted direction vector using the two-argument arctangent function. We then computed the MSE between reconstructed and true saccade directions after wrapping angular differences into the range [−180°, 180°]. To evaluate whether prediction accuracy exceeded chance, we generated null distributions of MSE by training neural networks with the same architecture on data with permuted saccade directions. Specifically, we randomly shuffled target direction vectors $[\sin(\theta), \cos(\theta)]$ across saccades 100 times and repeated the full cross-validation training and evaluation procedure for each permutation.

#### Kernel regression by minimizing the von Mises negative log-likelihood

To test whether SRND parameters could linearly explain saccade direction, we implemented a linear regression by minimizing the von Mises negative log-likelihood. The von Mises distribution is the circular analog of a Gaussian distribution for angular data, modeling angular data defined on the unit circle while accounting for the periodicity at the 0–$2\pi$ boundary. Formally, we modelled the saccade direction $\theta_i$ as a circular variable whose mean direction $\mu_i$ depends linearly on the SRND features:

$$\mu_i = X_i\beta,$$

where $X_i$ is the feature vector comprising SRND parameters and electrode coordinates, and $\beta$ is the vector of regression coefficients to be estimated. Under the von Mises assumption, the probability density of observing $\theta_i$ is given by

$$p(\theta_i \mid \mu_i, \kappa) = \frac{1}{2\pi I_0(\kappa)} \exp[\kappa \cos(\theta_i - \mu_i)],$$

where $\kappa$ is the concentration parameter which we set to 1. $I_0(\kappa)$ is the modified Bessel function of the first kind of order 0. We estimated the regression parameters by minimizing the von Mises negative log-likelihood:

$$\mathcal{L}(\beta) = -\sum_i \kappa \cos(\theta_i - X_i\beta) + N\log\big(2\pi I_0(\kappa)\big),$$

To fit this model, we used the BFGS optimization algorithm implemented via the *minimize* function from the *scipy.optimize* Python package. We obtained the predicted saccade direction from the model as

$$\widehat{\mu_i} = X_i\hat{\beta},$$

$$\widehat{\theta_i} = \widehat{\mu_i}\ mod\ 2\pi,$$

where $\hat{\beta}$ denotes the estimated model parameters, and $mod$ denotes the modulo operation, which ensures that predicted directions remain within the circular domain $[0, 2\pi)$.

Consistently, we evaluated model performance using contact- and saccade-level MSEs between predicted and true saccade directions. We assessed the statistical significance of prediction accuracy by comparing MSEs to their null distributions generated via 100 permutations of saccade labels.

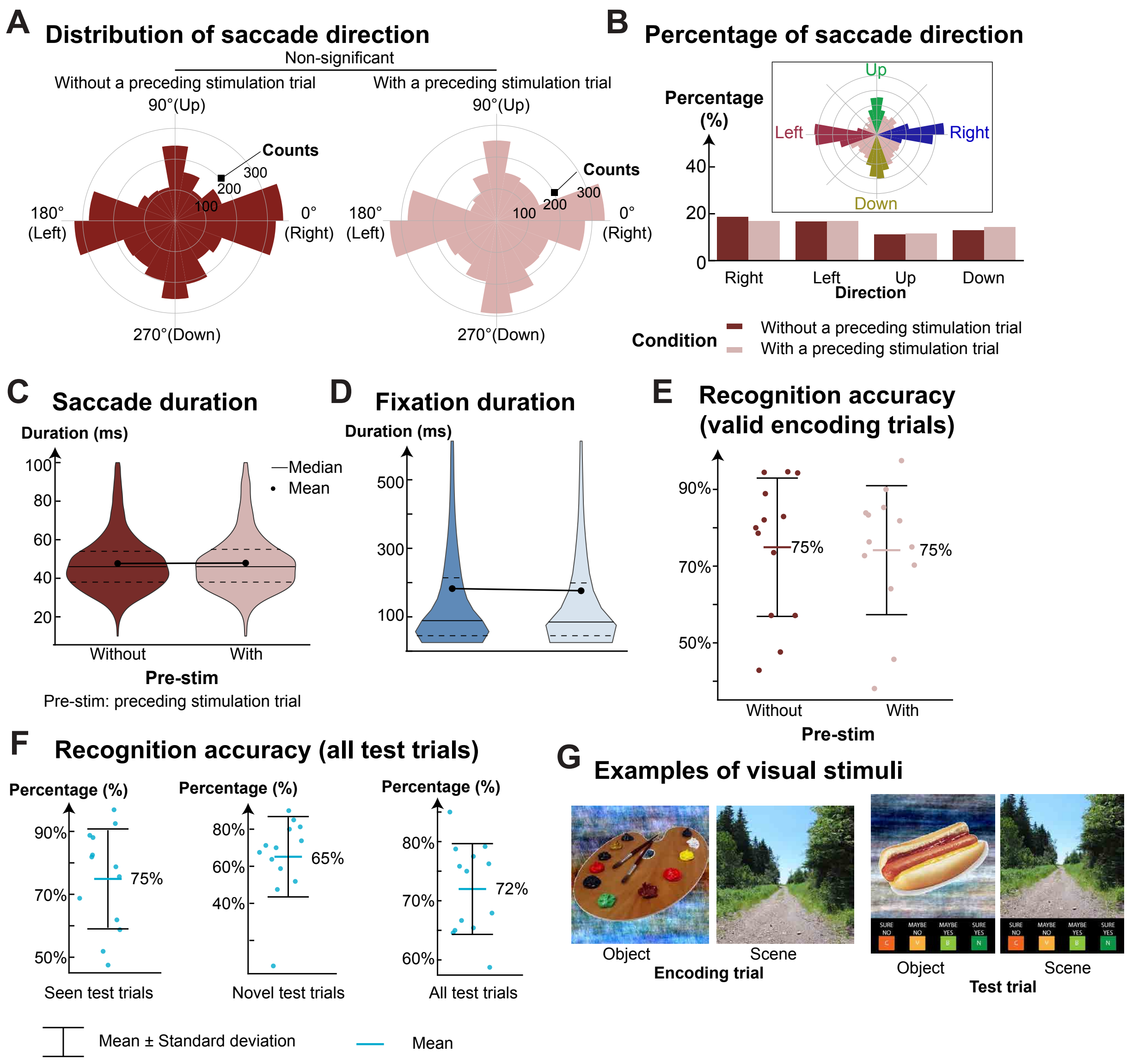


**Fig. S1 (Figure 1–figure supplement 1). Control analysis to rule out any latent effect of electrical stimulation on saccade behavior and recognition memory.** This study is part of a clinical trial investigating the effects of electrical stimulation of the basolateral amygdala on visual memory encoding (NCT05065450). Cortical stimulation was administered during 50% of encoding trials. For this study, we excluded all stimulation trials. Comparison of saccade behavior and recognition memory performance for trials without and with a preceding stimulation trial did not show latent effects of electrical stimulation. **(A)** Distribution of saccade directions. We found no significant difference in saccade directions for trials without and with a preceding stimulation trial (Watson-Williams test, F-statistics (1, 7290) = 0.023, p = 0.879). **(B)** Percentage of saccades that are categorized as upward, downward, leftward, or rightward. The inset illustrates the definition of saccade direction: a saccade is classified as “rightward” if its angle lies within ±20° around the horizontal axis (0°). **(C–D)** Saccade and fixation duration for trials without and with a preceding stimulation trial. The effect size comparing durations between the two conditions was trivial (|Cohen’s d| < 0.05). **(E)** Recognition accuracy for trials without and with a preceding stimulation trial (Cohen’s d = -0.044). **(F)** Recognition accuracy for seen (e.g., old) test trials, novel test trials, and all test trials. **(G)** Examples of visual stimuli used in the encoding and test trials.

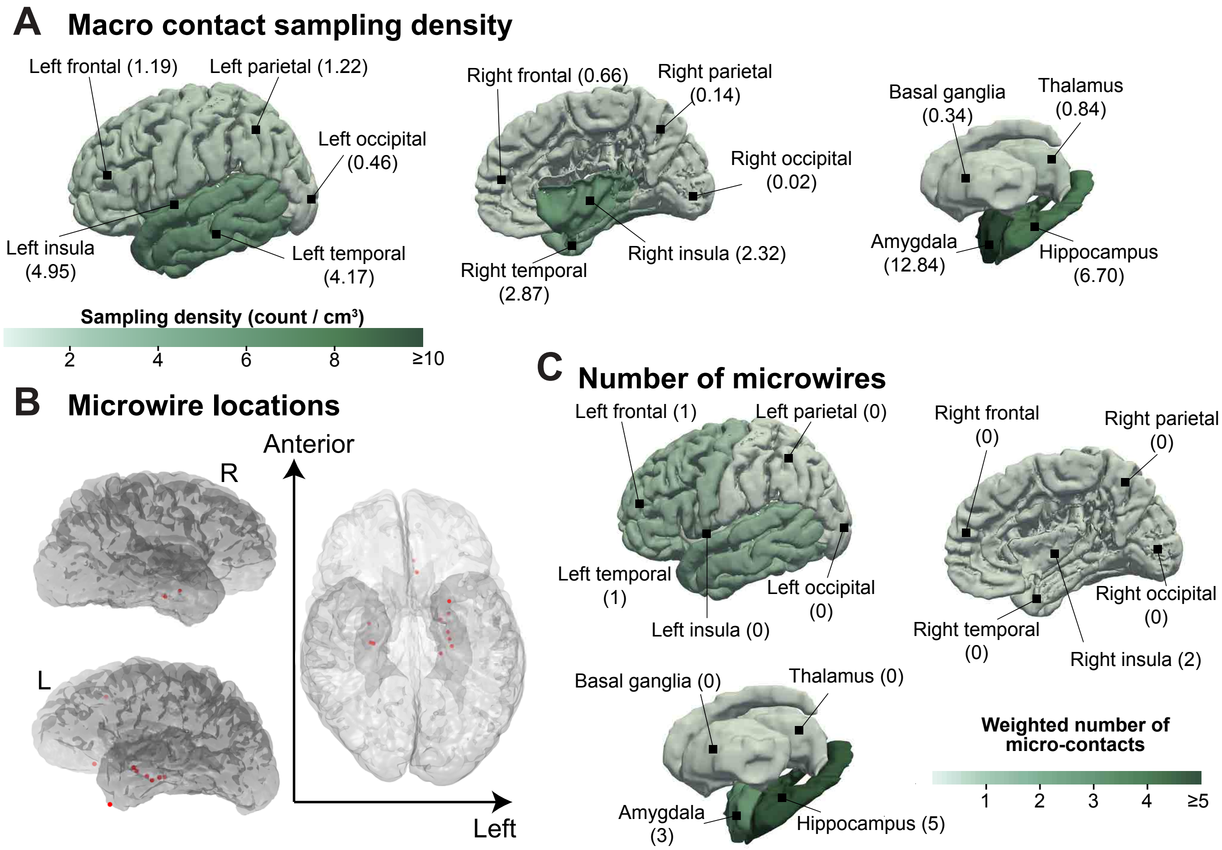


**Fig. S2 (Figure 1–figure supplement 2). Macro electrode sampling density and microwire locations**. Six subjects were each implanted with two Ad-Tech Behnke-Fried depth electrodes, each containing eight macro contacts and eight microwires. We refer to the bundle of microwires emerging from the tip of each electrode as a micro-contact location. **(A)** Sampling density for macro-electrode contacts, calculated by dividing the weighted contact count within each brain region by the volume of that region. **(B)** Location of microwires on the brain model of a reference subject. **(C)** Number of micro-contacts in major brain regions.

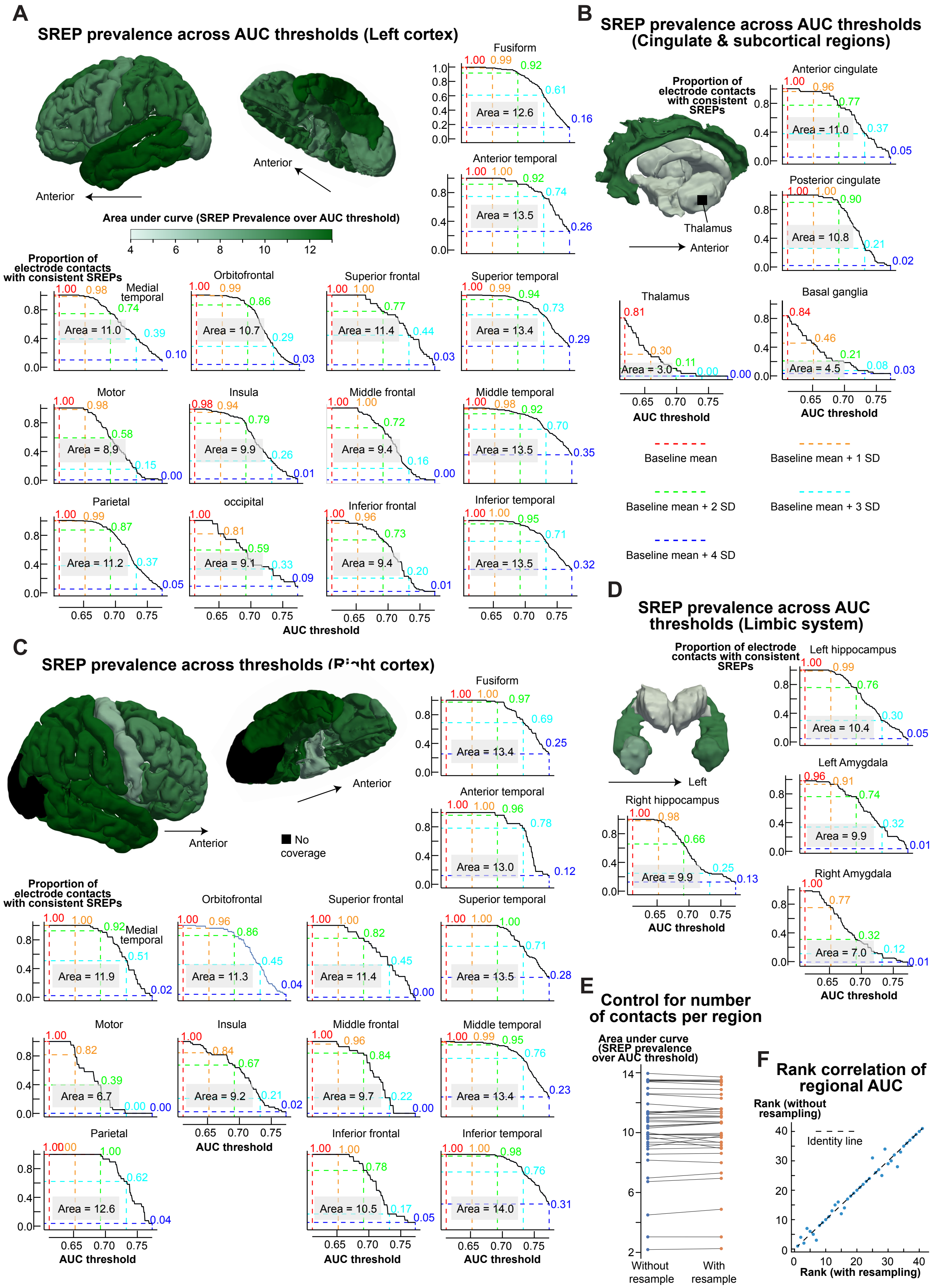
A
SREP prevalence across AUC thresholds (Left cortex)
Anterior
Area under curve (SREP Prevalence over AUC threshold)
4 6 8 10 12
Proportion of electrode contacts with consistent SREPs
Fusiform
Area = 12.6
Anterior temporal
Area = 13.5
Medial temporal
Area = 11.0
Orbitofrontal
Area = 10.7
Superior frontal
Area = 11.4
Superior temporal
Area = 13.4
Motor
Area = 8.9
Insula
Area = 9.9
Middle frontal
Area = 9.4
Middle temporal
Area = 13.5
Parietal
Area = 11.2
occipital
Area = 9.1
Inferior frontal
Area = 9.4
Inferior temporal
Area = 13.5
AUC threshold
B
SREP prevalence across AUC thresholds (Cingulate & subcortical regions)
Proportion of electrode contacts with consistent SREPs
Thalamus
Anterior
Anterior cingulate
Area = 11.0
Posterior cingulate
Area = 10.8
Thalamus
Area = 3.0
Basal ganglia
Area = 4.5
AUC threshold
Baseline mean
Baseline mean + 1 SD
Baseline mean + 2 SD
Baseline mean + 3 SD
Baseline mean + 4 SD
C
SREP prevalence across thresholds (Right cortex)
Anterior
No coverage
Fusiform
Area = 13.4
Anterior temporal
Area = 13.0
Proportion of electrode contacts with consistent SREPs
Medial temporal
Area = 11.9
Orbitofrontal
Area = 11.3
Superior frontal
Area = 11.4
Superior temporal
Area = 13.5
Motor
Area = 6.7
Insula
Area = 9.2
Middle frontal
Area = 9.7
Middle temporal
Area = 13.4
Parietal
Area = 12.6
Inferior frontal
Area = 10.5
Inferior temporal
Area = 14.0
AUC threshold
D
SREP prevalence across AUC thresholds (Limbic system)
Proportion of electrode contacts with consistent SREPs
Left hippocampus
Area = 10.4
Left Amygdala
Area = 9.9
Left
Right hippocampus
Area = 9.9
Right Amygdala
Area = 7.0
AUC threshold
E
Control for number of contacts per region
Area under curve (SREP prevalence over AUC threshold)
Without resample
With resample
F
Rank correlation of regional AUC
Rank (without resampling)
Identity line
Rank (with resampling)

**Fig. S3 (Figure 2–figure supplement 1)**. **The percentage of electrodes showing consistent SREP across AUC thresholds**. **(A)** SREP consistency in the left cortex. Each panel shows the proportion of electrode contacts whose peri-saccadic AUC is over AUC thresholds. The color-coded dashed lines represent the reference thresholds: mean, mean + 1 standard deviation (SD), mean + 2 standard deviation (SD), mean + 3 standard deviation (SD), and mean + 4 standard deviation (SD) of baseline AUC. The color of the brain model reflects the area under the SREP-prevalence-over-AUC-threshold curve, representing the robustness of the spatial distribution of SREP prevalence. **(B–D)** SREP consistency in the right cortex, subcortical regions, and cingulate cortex. **(E)** Areas under the curve of the SREP prevalence across the AUC thresholds. To control for the number of sampling electrodes across regions, we resampled electrodes with replacement to equalize sample sizes, then repeated the calculation of the area under the curve. Each dot represents the area under the curve for each region with or without resampling. **(F)** Correlation between AUCs derived from original and resampled data. The strong correlation revealed that the number of sampling electrodes has a trivial effect on the rank of SREP consistency (Spearman's $\rho = 0.987$, $p < 0.001$, n = 37, two-tailed t-test).

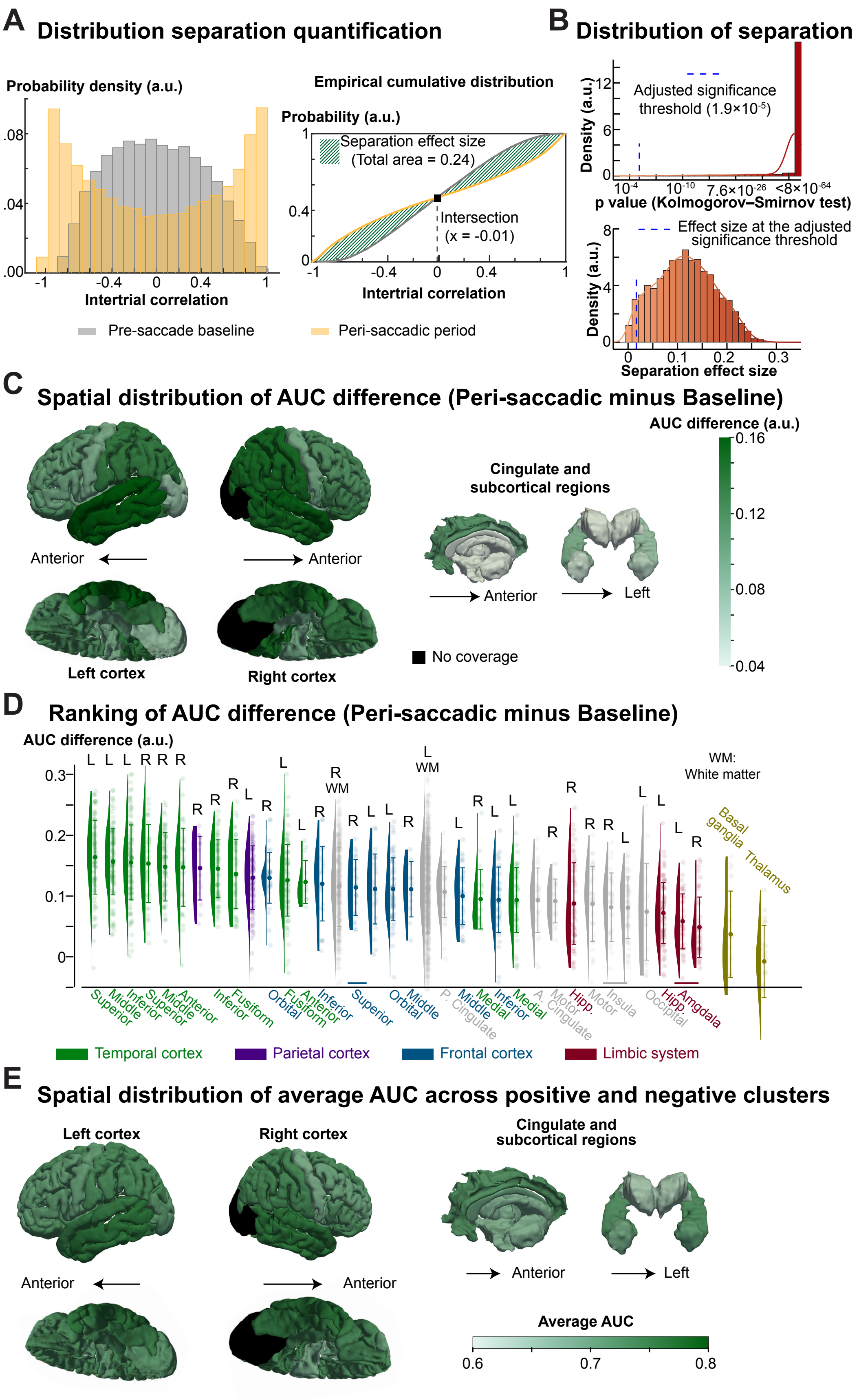
A Distribution separation quantification
Probability density (a.u.)
Intertrial correlation
Pre-saccade baseline
Peri-saccadic period
Empirical cumulative distribution
Probability (a.u.)
Separation effect size (Total area = 0.24)
Intersection (x = -0.01)
B Distribution of separation
Adjusted significance threshold (1.9×10-5)
Density (a.u.)
p value (Kolmogorov–Smirnov test)
Effect size at the adjusted significance threshold
Separation effect size
C Spatial distribution of AUC difference (Peri-saccadic minus Baseline)
AUC difference (a.u.)
Cingulate and subcortical regions
Anterior
Left
No coverage
Left cortex
Right cortex
D Ranking of AUC difference (Peri-saccadic minus Baseline)
AUC difference (a.u.)
WM: White matter
Basal ganglia
Thalamus
Temporal cortex
Parietal cortex
Frontal cortex
Limbic system
E Spatial distribution of average AUC across positive and negative clusters
Left cortex
Right cortex
Cingulate and subcortical regions
Anterior
Left
Average AUC

**Fig. S4 (Figure 2–figure supplement 2). Quantifying the regional saccade-related EP consistency based on intertrial correlation distributions**. **(A)** Quantification of the separation between intertrial correlation distributions during the baseline and peri-saccadic periods. To statistically assess the presence of robust and bimodal saccade-related EP, we quantified the separation (e.g., deviation from normal distribution toward bimodal distribution) between the intertrial correlation distributions during the peri-saccadic and baseline periods. Specifically, we computed the intersection point of their cumulative distribution functions and measured the absolute difference in the area under the curve (AUC) between them. The striped region illustrates the absolute difference in AUC. **(B)** Distributions of p-value from Kolmogorov-Smirnov tests and the distribution of the separation effect size. The significance threshold was corrected for multiple comparisons. The blue dashed line marks the bimodal deviation corresponding to the significance threshold. The effect size corresponding to the adjusted significance threshold is 0.017. **(C)** The separation between the intertrial correlation distributions during the peri-saccadic and baseline periods across the cerebrum. The color reflects the difference in the area under the curve (computed separately for the two clusters and then averaged) of probability density over intertrial correlation (**Figure 2D**). **(D)** The ranking of the magnitude of SEP consistency (AUC difference). Colors indicate anatomical regions. L: Left; R: Right. **(E)** The magnitude of SREP consistency across anatomical regions. We quantified the magnitude of EP consistency as the average area under the probability density curve for intertrial correlation following saccades across polarity clusters.

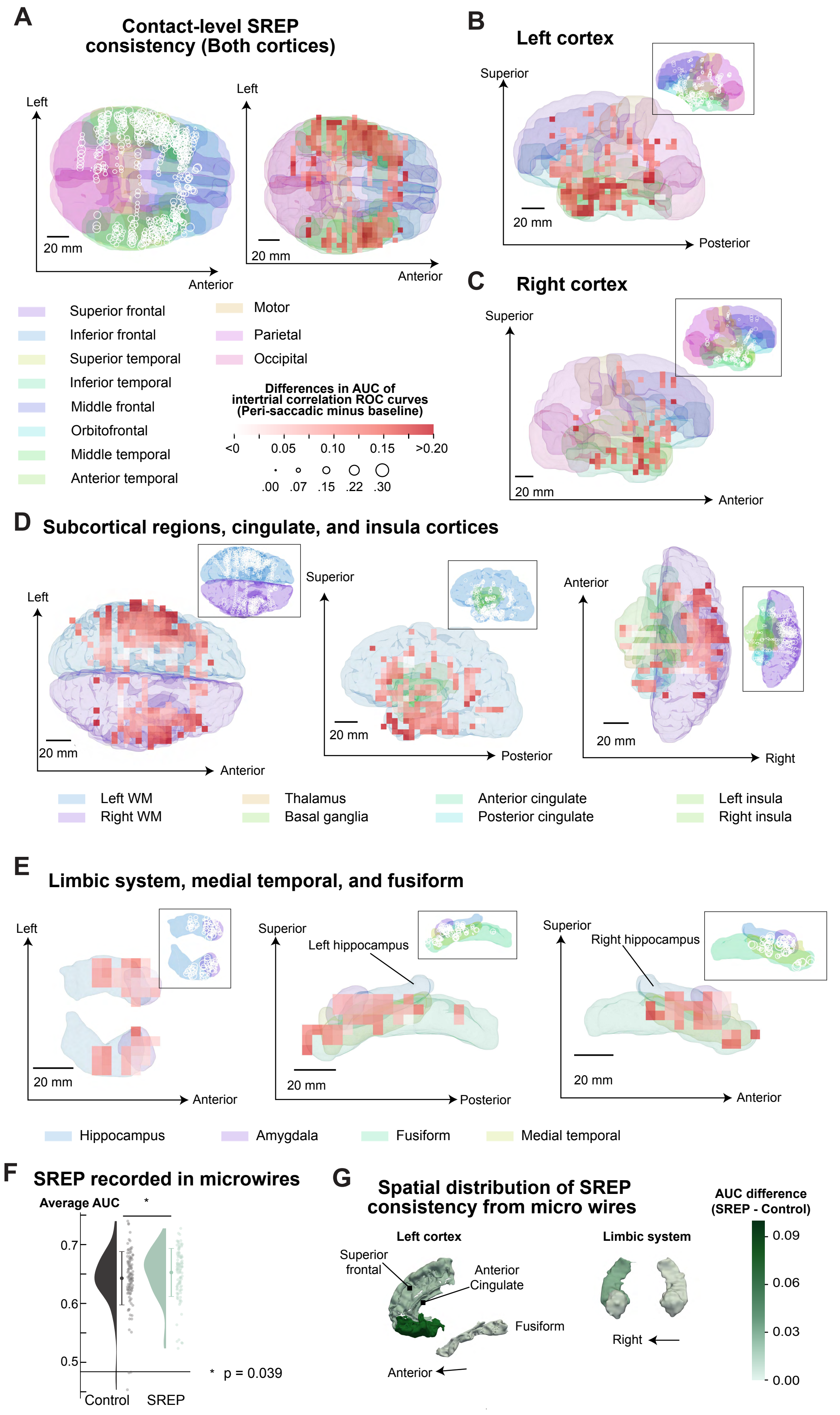
A
Contact-level SREP consistency (Both cortices)
Left
Anterior
20 mm
Superior frontal
Inferior frontal
Superior temporal
Inferior temporal
Middle frontal
Orbitofrontal
Middle temporal
Anterior temporal
Motor
Parietal
Occipital
Differences in AUC of intertrial correlation ROC curves (Peri-saccadic minus baseline)
<0 0.05 0.10 0.15 >0.20
.00 .07 .15 .22 .30
B
Left cortex
Superior
Posterior
C
Right cortex
Superior
Anterior
D
Subcortical regions, cingulate, and insula cortices
Left
Anterior
Superior
Posterior
Right
Left WM
Right WM
Thalamus
Basal ganglia
Anterior cingulate
Posterior cingulate
Left insula
Right insula
E
Limbic system, medial temporal, and fusiform
Left hippocampus
Right hippocampus
Hippocampus
Amygdala
Fusiform
Medial temporal
F
SREP recorded in microwires
Average AUC
0.7
0.6
0.5
Control
SREP
* p = 0.039
G
Spatial distribution of SREP consistency from micro wires
Left cortex
Limbic system
Superior frontal
Anterior Cingulate
Fusiform
Anterior
Right
AUC difference (SREP - Control)
0.09
0.06
0.03
0.00

**Fig. S5 (Figure 2–figure supplement 3). Electrode contact-level differences in the area under the curve (AUC) of intertrial correlation receiver operation characteristic (ROC) curves between baseline and peri-saccadic periods.** For each electrode contact and each condition (baseline vs peri-saccade), AUCs were calculated separately for the two SREP clusters and then averaged. AUC difference between conditions, the degree of SREP consistency in anatomical regions sampled by each electrode. To visualize spatial patterns and enable comparison of effects across panels (devoid of a 3D perspective), we projected the 3D brain surface and electrode coordinates onto a 2D plane. The overlay heatmap represents the spatial pattern of SREP consistency. To generate the heatmap, we discretized this 2D space into a regular grid with a 5 mm resolution. We computed the local average AUC difference within a 10 mm radius for each grid element. Heatmap elements that have no nearby sampling electrodes are fully transparent. **(A–E)**. Spatial pattern of SREP consistency in cortices and subcortical regions. The heatmap color bar is uniform across panels A–E. **(F)** Consistency of peri-saccadic SREPs recorded from micro wires. We calculated the area under the receiver operator characteristic curve from intertrial correlation distributions. We averaged the area under the curve (AUC) for the two clusters. The average peri-saccadic AUCs were higher than the control AUCs (Cohen's $d = 0.227$, two-tailed paired t-test, $n = 115$, t statistic = 2.088, $p < 0.039$). **(G)** Spatial distribution of SREP consistency across the cerebrum. Brain regions located within 5 mm of the microwire electrodes are shown, which include Left Hippocampus, Left Amygdala, Left White Matter, Right Hippocampus, Right Amygdala, Right White Matter, Left Fusiform, Left Superior Frontal, and Anterior Cingulate.

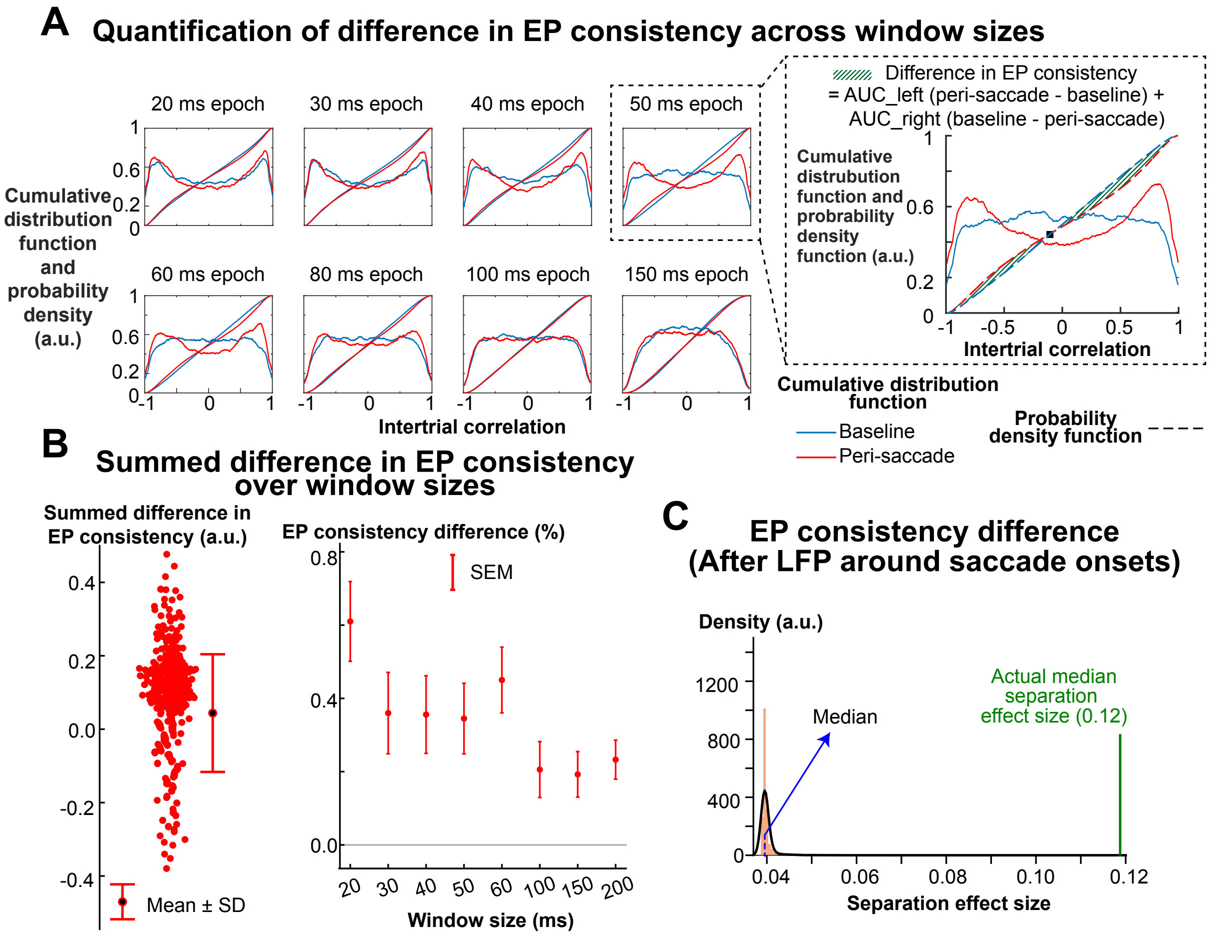


**Fig. S6 (Figure 2–figure supplement 4). Evoked potential (EP) consistency across window size. (A)** Quantification of bimodality as a measure of EP consistency. To determine the time over which the EP waveform remains stable, we calculated the intertrial correlation distribution across a range of window sizes (20, 30, 40, 50, 60, 100, 150, 200 ms). We defined the baseline (i.e., pre-saccadic) epoch starting from 1s prior to saccade onset, with the same length as the peri-saccadic epoch. This panel shows the cumulative distributions of intertrial correlations for an exemplary electrode (same electrode as shown in **Figure 2A**) across window sizes. As the window size increased, the degree of bimodality in the intertrial correlation distribution decreased. A more bimodal distribution of intertrial correlations, with values closer to 1 or -1, indicates greater EP consistency. To quantify EP consistency differences between baseline and peri-saccadic epochs, we computed the sum of two AUC differences: (1) For correlations smaller than the crossing point (black rectangle), we calculated $(peri-saccade\ AUC\ -\ pre-saccade\ AUC)$ . 2. For correlations greater than the crossing point, we calculated $(pre-saccade\ AUC\ -\ peri-$

$saccade\ AUC$). **(B)** EP consistency differences summed across window sizes. Similar to our main analysis (**Figure 2G**), we defined electrode contacts showing consistent EPs following saccades as those whose mean AUC of intertrial correlation following saccades exceeded the mean pre-saccade AUC by two standard deviations. For each such electrode contact, we computed EP consistency differences between baseline and peri-saccadic epochs, as illustrated in panel **A**, at each window size and summed these differences across window sizes. Across electrode contacts, the summed difference in EP consistency was significantly greater than 0 ($t = 5.349$, $p < 0.001$, Cohen's $d = 0.274$, $n = 382$, two-sided t-test). Across all window sizes, the peri-saccadic EPs consistently exhibited greater waveform consistency compared to the pre-saccadic baseline. **(C)** EP consistency difference following temporal shuffling of local field potentials. As a negative control, we disrupted the temporal structure by shuffling the local field potential data and repeated the analysis of the separation effect size. The median EP consistency difference between peri-saccadic and baseline EP was 0.04 in this negative control, significantly smaller than the observed separation effect size ($t = 65.086$, $p < 0.01$, Cohen's $d = 1.804$, two-sided t-test, $n = 2604$, compared to **Fig. S4A, B**), indicating our main findings remain robust.

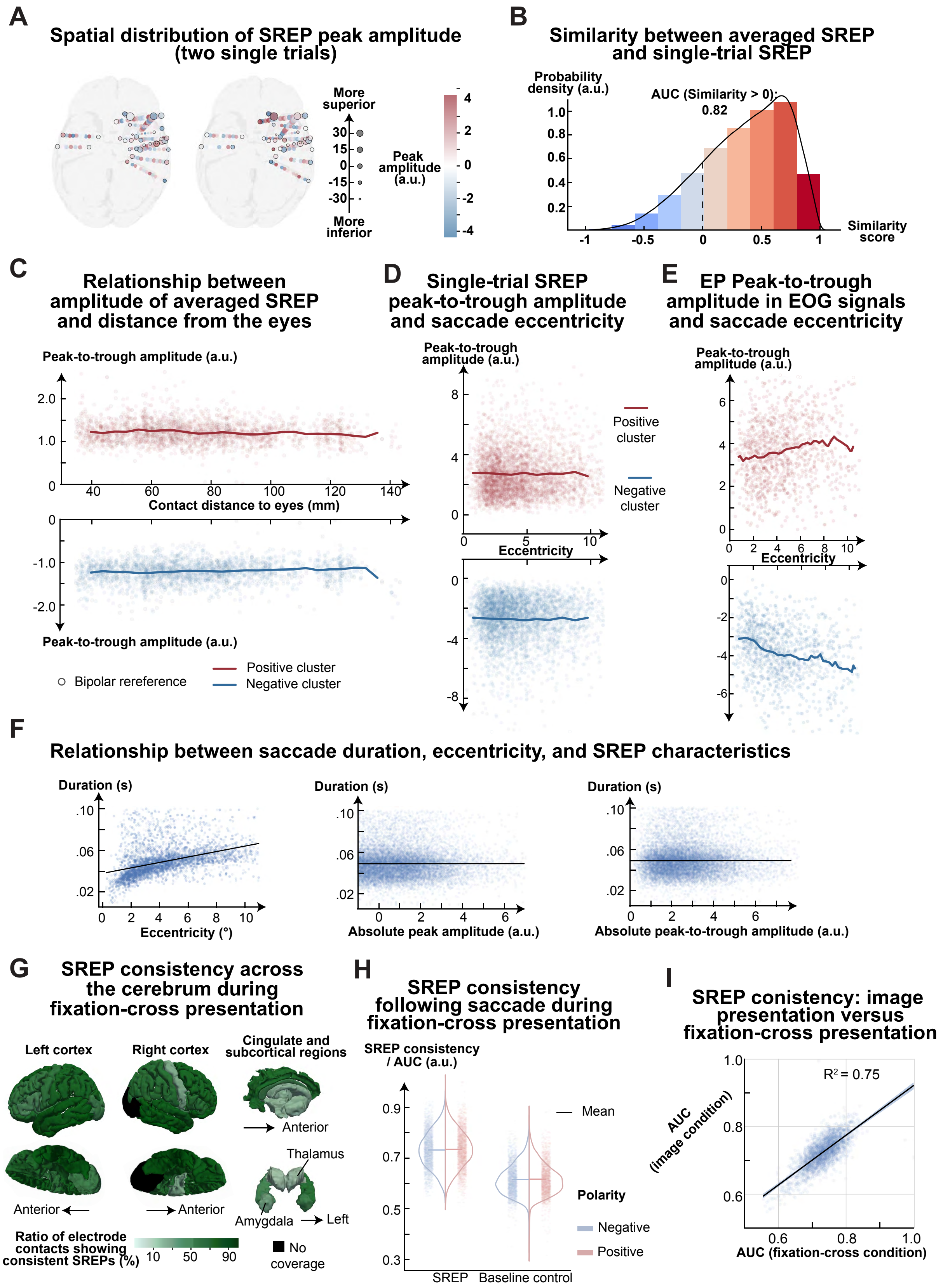
A
Spatial distribution of SREP peak amplitude (two single trials)
More superior
30
15
0
-15
-30
More inferior
Peak amplitude (a.u.)
4
2
0
-2
-4
B
Similarity between averaged SREP and single-trial SREP
Probability density (a.u.)
AUC (Similarity > 0): 0.82
Similarity score
C
Relationship between amplitude of averaged SREP and distance from the eyes
Peak-to-trough amplitude (a.u.)
Contact distance to eyes (mm)
Bipolar rereference
Positive cluster
Negative cluster
D
Single-trial SREP peak-to-trough amplitude and saccade eccentricity
Peak-to-trough amplitude (a.u.)
Eccentricity
Positive cluster
Negative cluster
E
EP Peak-to-trough amplitude in EOG signals and saccade eccentricity
Peak-to-trough amplitude (a.u.)
Eccentricity
F
Relationship between saccade duration, eccentricity, and SREP characteristics
Duration (s)
Eccentricity (°)
Absolute peak amplitude (a.u.)
Absolute peak-to-trough amplitude (a.u.)
G
SREP consistency across the cerebrum during fixation-cross presentation
Left cortex
Right cortex
Cingulate and subcortical regions
Anterior
Thalamus
Amygdala
Left
Ratio of electrode contacts showing consistent SREPs (%)
10
50
90
No coverage
H
SREP consistency following saccade during fixation-cross presentation
SREP consistency / AUC (a.u.)
Mean
Polarity
Negative
Positive
SREP
Baseline control
I
SREP conistency: image presentation versus fixation-cross presentation
R² = 0.75
AUC (image condition)
AUC (fixation-cross condition)

**Fig. S7 (Figure 2–figure supplement 5). SREPs are not oculomuscular signals**. **(A)** Spatial distribution of SREP peak amplitudes. This panel shows SREP peak amplitudes across intracranial electrode contacts for two saccades in a subject (Subject ID 1). Circle size indicates electrode depth along the superior-to-inferior axis, with smaller circles representing deeper (i.e., more inferior) electrode contacts. We did not observe any spatial pattern in polarity or peak amplitude. **(B)** Similarity between single-trial SREPs and the corresponding cluster-level averaged SREPs, assessed using Spearman's correlation. Similarity distributions were skewed towards 1 (positive correlation), with an area under the curve of 0.812 for correlations greater than 0, compared to 0.181 for correlations less than 0, indicating that most single-trial SREPs resemble their cluster-level averaged SREP. Consistency of SREP during fixation cross presentation, quantified using AUC as described in **Figure 2D**. During fixation cross presentation, SREP consistency was significantly higher than baseline (paired t-test, $p<0.001$, Cohen's $d = 2.631$, $N = 2604$). **(C)** Relationship between single-trial SREP peak-to-trough amplitude and saccade eccentricity. For both polarities, saccade eccentricity explained 0.0% of the variance in peak-to-trough amplitude. The x-axis represents the distance from the recording electrode to the nearest eye. The line represents the average peak-to-trough amplitude calculated over sliding windows of 10 mm, incremented by 5 mm. The distance explained 0.9% and 0.7% variance in the peak-to-trough amplitude of averaged SREPs for positive and negative clusters, respectively, across all electrode contacts. **(D)** Relationship between single-trial SREP peak-to-trough amplitude and saccade eccentricity. For both polarities, saccade eccentricity explained 0.0% of the variance in peak-to-trough amplitude. **(E)** Relationship between single-trial oculomuscular signal amplitudes and saccade eccentricity. We extracted electrooculogram (EOG) signals from frontopolar scalp EEG electrodes (Fp1 and Fp2) using bipolar re-referencing. Eccentricity explains 2.7% and 8.2% variance in SREP peak-to-trough amplitude for positive and negative clusters. **(F)** Relationship between saccade duration and eccentricity, SREP peak amplitude, and SREP peak to trough. Saccade duration and saccade eccentricity were positively correlated ($Pearson's\ r = 0.056$), while saccade duration was correlated less with SREP peak amplitude ($Pearson's\ r = -0.002$), or SREP peak to trough ($Pearson's\ r = 0.004$). **(G)** Response ratios during fixation-cross presentation across the cerebrum. The response ratio is defined as the proportion of electrode contacts with AUC in the peri-saccadic condition above the mean + 2 standard deviation (SD) of baseline AUC. The AUC calculation is illustrated in **Figure 2G**. The brain was segmented into

White Matter (Left, Right), Hippocampus (Left, Right), Amygdala (Left, Right), Basal Ganglia, Thalamus, Orbitofrontal (Left, Right), Inferior Frontal (Left, Right), Middle Frontal (Left, Right), Superior Frontal (Left, Right), Anterior Cingulate, Posterior Cingulate, Motor (i.e., precentral gyrus and paracentral lobule, Left, Right), Anterior Temporal (Left, Right), Middle Temporal (Left, Right), Superior Temporal (Left, Right), Inferior Temporal (Left, Right), Medial Temporal (Left, Right), Fusiform (Left, Right), Parietal (Left, Right), Occipital (Left, Right), and Insula (Left, Right). **(H)** Consistency of SREP during fixation-cross presentation, quantified using AUC as described in **Figure 2D**. During fixation cross presentation, SREP consistency was significantly higher than baseline (paired t-test, $p<0.001$, Cohen's $d = 2.631$, $N = 2604$). **(I)** Correlation between SREP consistency during image presentation and during fixation-cross presentation.

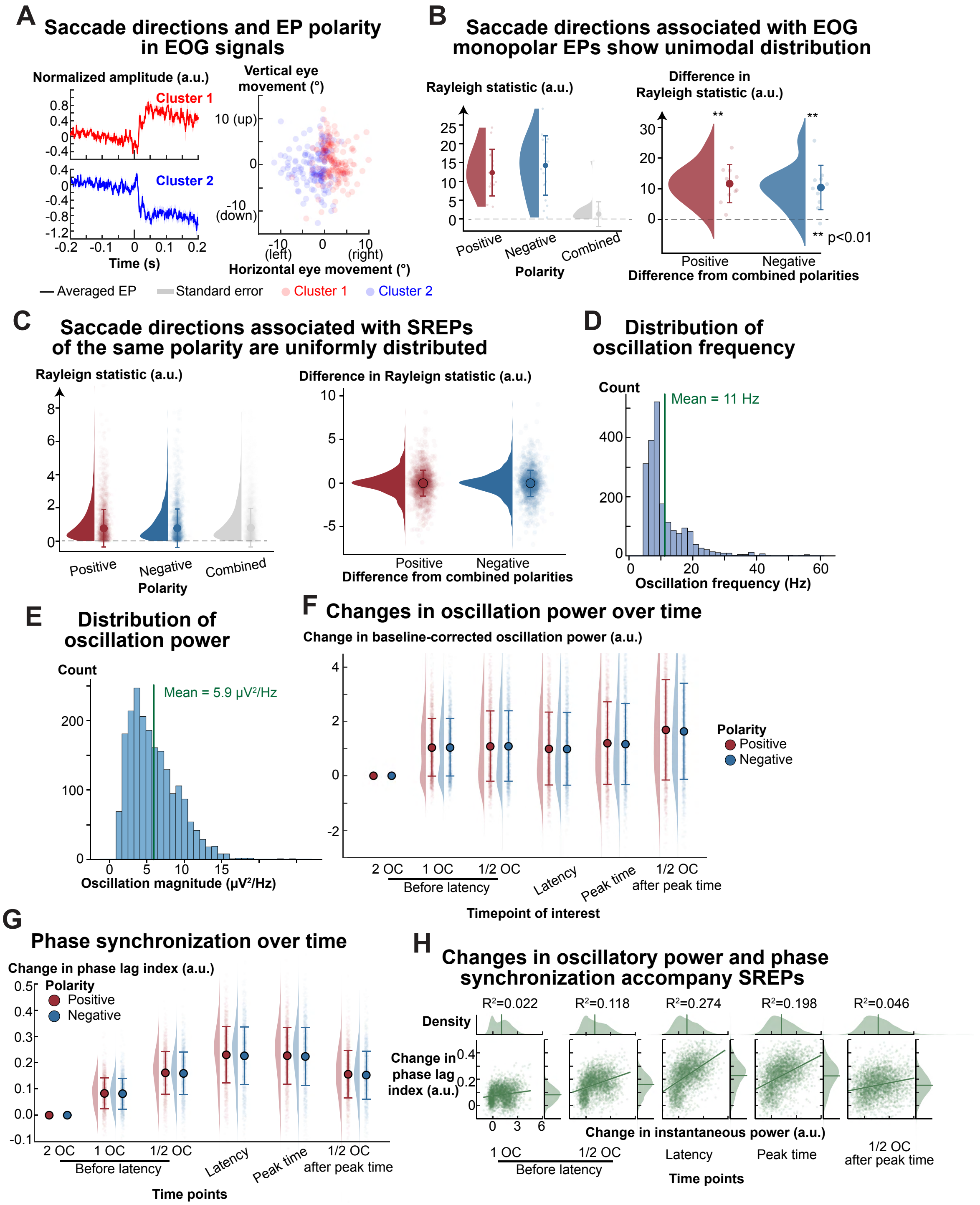


**Fig. S8 (Figure 3–figure supplement 1). Saccade direction is associated with the polarity of EOG EPs but not the polarity of SREPs extracted from intracranial LFPs. (A)** Relationship between saccade direction and EP polarity in EOG signals. We applied the same clustering and EP characterization procedures to EOG signals. The left panel shows the positive and negative EOG

EPs averaged across trials from a representative subject. Time 0 is the saccade offset. The right panel shows the saccade direction associated with the positive and negative EPs. The shaded area in the left panel represents the standard error. **(B)** Saccade directions were unimodally distributed in monopolar EOG EPs. Assuming EOG signals captured oculomuscular artifacts during saccades, we expected the circular distribution of saccade direction associated with monopolar EP to have a unimodal peak. We used Rayleigh statistics to quantify this unimodal bias. Rayleigh statistics for monopolar EPs were higher than the Rayleigh statistics for combined EPs. Each data point on the plot represents Rayleigh statistics calculated from EOG signals for one subject and one task. We ensured the same sample size when calculating the Rayleigh statistics. We defined the resampling size as the $20^{th}$ percentile of the number of trials ($n = 32$) for each subject, each task, and each electrode contact. We sampled the same number of saccade directions associated with monopolar EPs and saccade directions associated with the combined EPs. The right panel shows the distribution of Rayleigh statistic differences (monopolar – combined). For both polarities, differences in Rayleigh statistics were significantly greater than zero (For positive polarity, $t = 5.950$, $p < 0.001$, Cohen's $d = 1.881$, paired t-test, $n = 15$; for negative polarity, $t = 4.553$, $p = 0.001$, Cohen's $d = 1.440$, $n = 18$). In addition, the circular distribution of saccade direction was significantly different between positive and negative polarity ($p < 0.001$ for all subjects, 95% CI for W statistics = [33.223, 46.050], Mardia-Watson-Wheeler test). **(C)** Saccade directions associated with positive and negative SREPs measured with stereoelectroencephalography. For both polarities, Rayleigh statistic differences (monopolar – combined) were centered at zero (For positive polarity, $t=-0.953$, $p = 0.341$, Cohen's $d = -0.022$, two-tailed t-test, $N = 2068$; for negative polarity, $t = -1.028$, $p = 0.304$, Cohen's $d = -0.023$, $N = 2058$). To estimate the difference between the saccade directions associated with positive and negative SREPs, we sampled the same number of saccade directions associated with monopolar EPs. Consistently, we defined the resampling size as the $20^{th}$ percentile of the number of trials ($n = 62$) for each subject, each task, and each electrode contact. Across subjects, tasks, and electrode contacts, we found no difference in saccade direction between polarities (95% CI for W statistics = [1.926, 2.107], the corresponding 95% CI for p-value is [0.486, 0.512], Mardia-Watson-Wheeler test). **(D)** Distribution of oscillation frequencies in intracranial signals. The mean oscillatory frequency is 11 Hz. To minimize potential confounds from line noise, we identified oscillation frequency as the frequency between 1 and 58 Hz at which the difference between the power spectral density and its one-over-frequency fit was maximal. **(E)**

Distribution of oscillation power in intracranial signals. Measured as the difference in power spectral density from a one-over-frequency fit at the oscillatory frequency. **(F)** Changes in oscillation power (i.e., instantaneous power at the oscillatory frequency) over time. To enable cross-subject and cross-contact comparisons, we normalize oscillation power to power at two oscillation cycles before the EP latency. The effect sizes (Cohen's d) between phase lag index at 2 OCs before latency and subsequent time points (from 1 OC before latency to 1 OC after peak time) were 0.894, 0.780, 0.694, 0.742, and 0.860 (N = 5300). (**G**) Phase synchronization induced by SREPs. To quantify phase coherence around saccades, we estimated instantaneous power and phase using unmasked signals, in contrast to panel A. The effect size (Cohen's d) between phase lag index at 2 OC before latency and subsequent time points (from 1 OC before SREP onset latency to 1 OC after SREP peak time) was 1.214, 1.625, 1.739, 1.704, and 1.427. The corresponding rank-based effect sizes (rank-biserial correlation) were 0.924, 0.980, 0.988, 0.982, and 0.959. Phase lag index significantly increased after 2 OCs before latency ($p < 0.001$, Wilcoxon paired test, n = 5300). (**H**) Relationship between oscillation power and phase synchrony. Oscillation power, measured as instantaneous power, began increasing from 2 OCs before the SREP latency and remained elevated (see panel **F**). The positive correlation between instantaneous power and phase synchrony was strongest at SREP onset latency and weaker at other time points. The density plots at the top and right represent the unimodal distributions of instantaneous power and instantaneous phase, with their means marked.

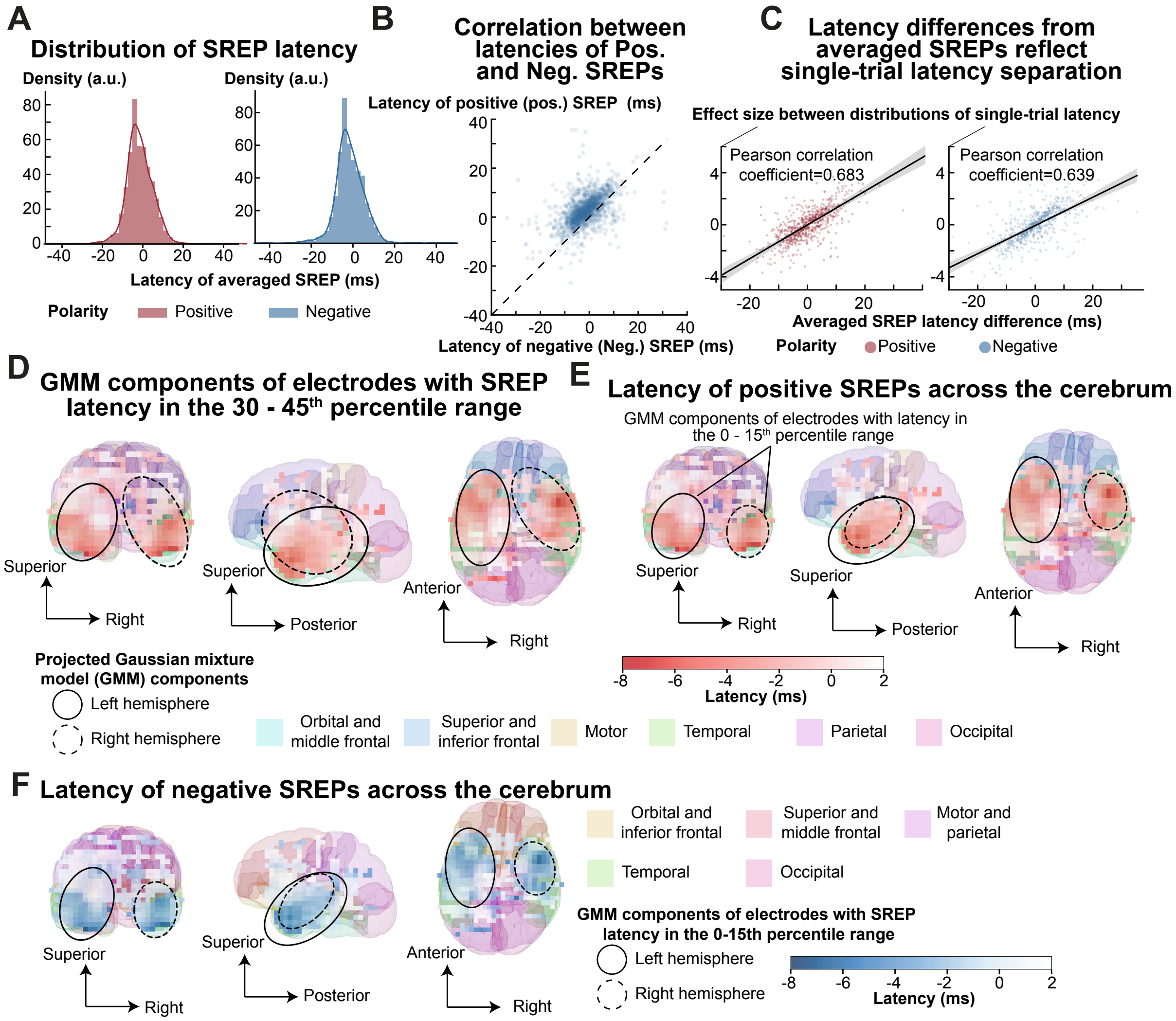


**Fig. S9 (Figure 4–figure supplement 1). SREP latency across the cerebrum. (A)** Averaged SREP latency. The mean latency was -1 ms for both positive and negative polarities. The standard deviation was 7.5 ms for positive SREPs and 7.7 ms for negative SREPs, respectively. **(B)** Correlation between latencies of averaged SREPs with positive and negative polarities. They were positively correlated (Pearson's correlation coefficient = 0.436, $p < 0.001$, exact test for Pearson's correlation, n = 1558). **(C)** Latency differences between trial-averaged SREPs across electrode contact pairs positively correlated with the effect size of latency differences estimated from single-trial SREPs ($p < 0.001$, exact test for Pearson's correlation, i.e., t-test). Given the large number of possible electrode-contact pairs, we randomly sampled 2,000 contact pairs per polarity to reduce computational cost. We randomly sampled 2000 pairs of electrode contacts for each polarity. Illustration of two Gaussian Mixture Model (GMM) components estimated from electrodes with SREP latency in the 30 – 45$^{th}$ percentile range. Each ellipse represents a GMM component, derived from the centroid and standard deviations along the three spatial axes of the 3D Gaussian distributions. The heatmap represents the average SREP latency averaged across polarities. The underlying projected 3D brain model provides an anatomical reference. As in **Fig. S5**, the color of each heatmap element (5 mm by 5 mm) represents the average latency across electrode contacts localized within 10 mm of that element. **(E)** The spatial distribution of latency for positive SREPs. **(F)** The spatial distribution of latency for negative SREPs. Darker blue in the heatmap denotes earlier SREPs. The color of the underlying 3D brain is adapted from panel E to avoid overlap with the blue-to-white latency heatmap. In E and F**,** the solid and dashed ellipses represent left and right GMM components estimated from the spatial location of electrodes with SREP latency in the 0 – $15^{th}$ percentile range.

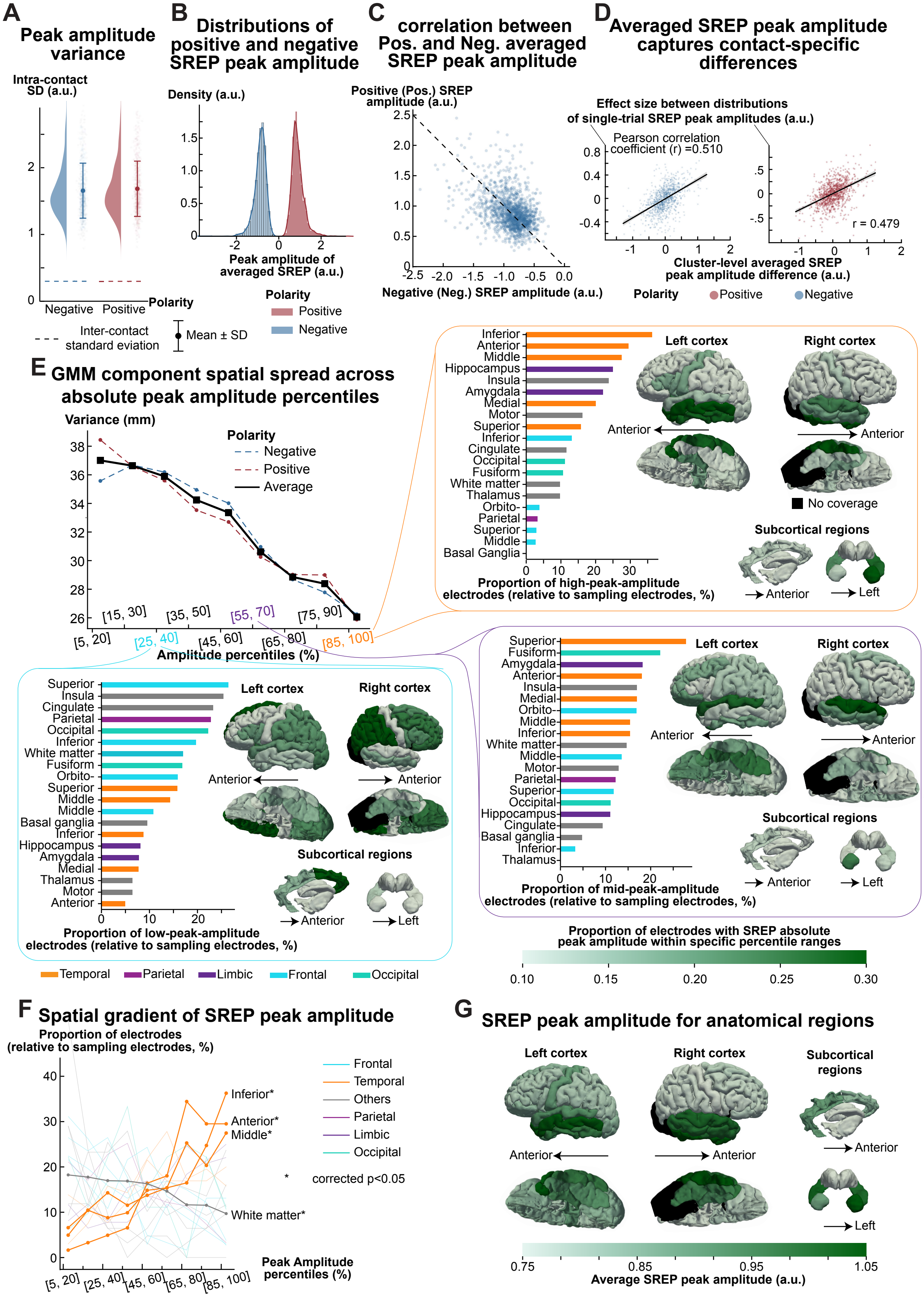
A
Peak amplitude variance
Intra-contact SD (a.u.)
Negative Positive
Polarity
Inter-contact standard deviation
Mean ± SD
B
Distributions of positive and negative SREP peak amplitude
Density (a.u.)
Peak amplitude of averaged SREP (a.u.)
Polarity
Positive
Negative
C
correlation between Pos. and Neg. averaged SREP peak amplitude
Positive (Pos.) SREP amplitude (a.u.)
Negative (Neg.) SREP amplitude (a.u.)
D
Averaged SREP peak amplitude captures contact-specific differences
Effect size between distributions of single-trial SREP peak amplitudes (a.u.)
Pearson correlation coefficient (r) =0.510
r = 0.479
Cluster-level averaged SREP peak amplitude difference (a.u.)
Polarity
Positive
Negative
E
GMM component spatial spread across absolute peak amplitude percentiles
Variance (mm)
Polarity
Negative
Positive
Average
Amplitude percentiles (%)
[5, 20] [15, 30] [25, 40] [35, 50] [45, 60] [55, 70] [65, 80] [75, 90] [85, 100]
Proportion of high-peak-amplitude electrodes (relative to sampling electrodes, %)
Proportion of mid-peak-amplitude electrodes (relative to sampling electrodes, %)
Proportion of low-peak-amplitude electrodes (relative to sampling electrodes, %)
Left cortex
Right cortex
Anterior
No coverage
Subcortical regions
Left
Temporal
Parietal
Limbic
Frontal
Occipital
Proportion of electrodes with SREP absolute peak amplitude within specific percentile ranges
0.10 0.15 0.20 0.25 0.30
F
Spatial gradient of SREP peak amplitude
Proportion of electrodes (relative to sampling electrodes, %)
Inferior*
Anterior*
Middle*
White matter*
Frontal
Temporal
Others
Parietal
Limbic
Occipital
* corrected p<0.05
Peak Amplitude percentiles (%)
G
SREP peak amplitude for anatomical regions
Left cortex
Right cortex
Subcortical regions
Anterior
Left
0.75 0.85 0.95 1.05
Average SREP peak amplitude (a.u.)

**Fig. S10 (Figure 4–figure supplement 2)**. **Spatial distribution of SREP peak amplitude**. **(A)** Comparison between intra-contact single-trial SREP peak amplitude variance and inter-contact single-trial SREP peak amplitude variance. The average standard deviation of intra-contact SREP peak amplitudes was 1.656 and 1.685 for negative and positive SREPs, respectively. The standard deviation of intra-contact SREP peak amplitudes was significantly higher compared to inter-contact SREP peak amplitudes of 0.299 and 0.298 (N = 786, W statistics = 0, p < 0.001, Rank-biserial correlation = 1 for negative polarity; N = 797, W statistics = 0, p < 0.001, Rank-biserial correlation = 1 for positive polarity; Wilcoxon signed-rank test). **(B)** Peak amplitudes of cluster-level averaged SREPs. The mean peak amplitude was 0.897 and -0.883 for positive and negative polarities, respectively. The standard deviation was 0.296 for positive and negative SREPs. **(C)** Correlation between positive and negative averaged SREP peak amplitudes. They were negatively correlated (Pearson's correlation coefficient = -0.365, p < 0.001, exact test for Pearson's correlation). **(D)** Correlation between peak amplitude differences between cluster-level averaged SREPs across electrode contact pairs and the effect size of peak amplitude differences estimated from single-trial SREPs. They were positively correlated (p < 0.001, exact test for Pearson's correlation). **(E)** Relationship between spatial spread and absolute SREP peak amplitude percentile ranges (related to **Figure 4D**). For illustration, we selected high-, mid-, and low-amplitude percentile ranges ([0–15], [30–45], and [60–75]) and plotted the proportion of electrodes exhibiting SREPs with peak amplitude within each range. We calculated the proportion as the ratio between the number of electrodes in a region with SREP peak amplitude within a percentile range and the total number of electrodes localized in that region. The accompanying 3D brain plots show the anatomical distribution of these proportions across regions. **(F)** Proportion of electrodes with absolute SREP peak amplitude at a specific percentile range per region across absolute amplitude percentile bins. Thick lines represent regions where the proportion changed monotonically across amplitude percentile ranges based on Spearman's correlation. We found that anterior ($\rho =\ 0.95,$ Bonferroni corrected asymptotic $p = 0.002,$ number of comparisons = 20), inferior ($\rho =\ 0.95,$ $corrcted\, p = 0.001$), and middle ($\rho =\ 0.93,\ corrcted\, p = 0.005$) temporal regions had a lower proportion of electrodes showing SREPs with low absolute peak amplitude and a higher proportion of electrodes showing SREPs with high absolute peak amplitude. White matter ($\rho = -1,\ corrcted\, p\ <\ 0.001$) showed the opposite pattern. **(G)** Spatial distribution of absolute SREP peak amplitudes averaged across electrode contacts localized within an anatomical region.

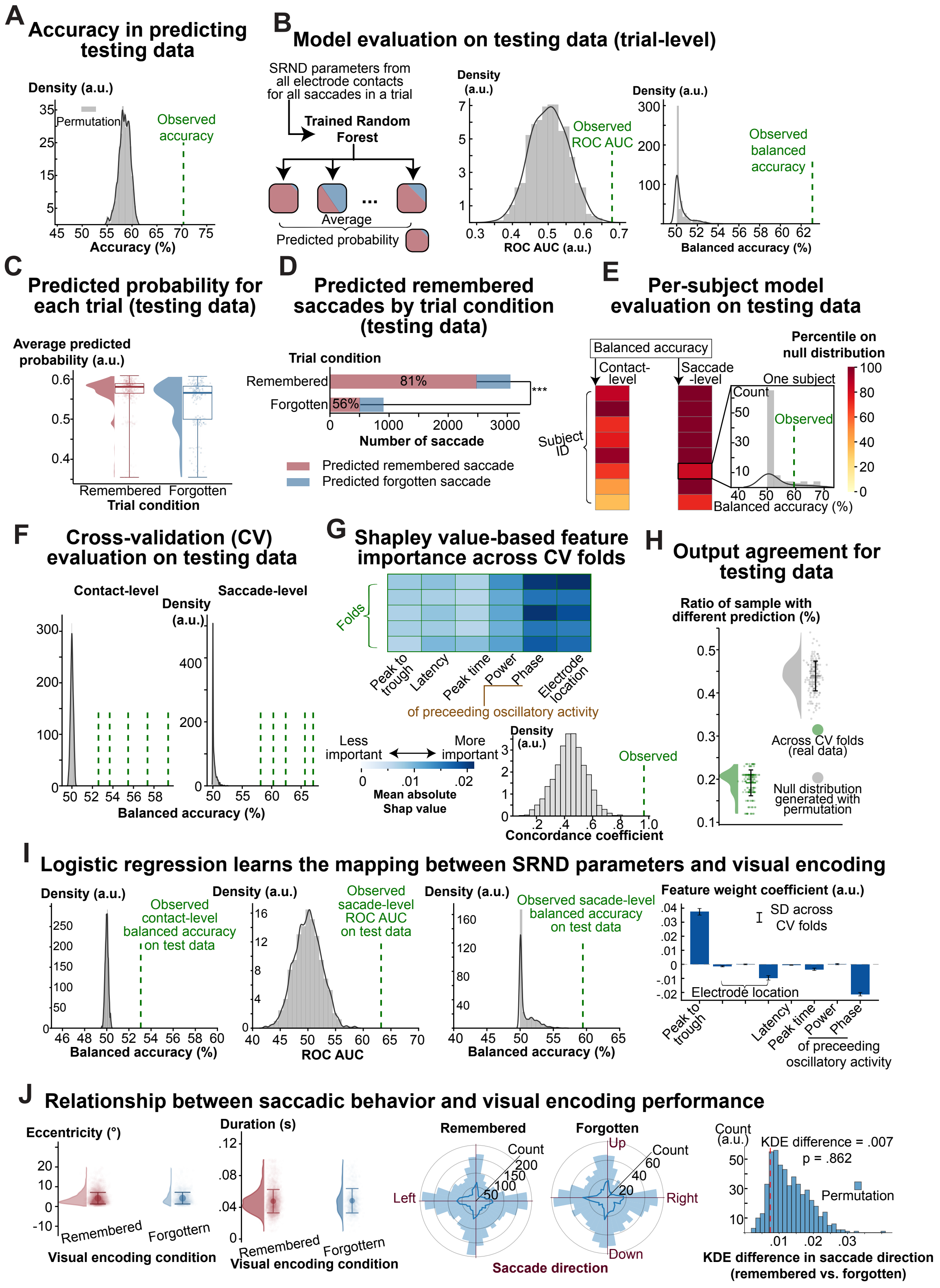


**Fig. S11 (Figure 5–figure supplement 1). Validation of the model's ability to learn the mapping between SRND parameters and visual encoding**. **(A)** Accuracy of the random forest in predicting visual encoding performance on testing data. We performed 200 permutations to generate a null distribution of accuracy. The observed accuracy was 70.3%, exceeding all values in the null distribution. **(B)** Trial-level model performance on test data. We used SRND parameters in all electrode contacts across all saccades during a trial as input to the model and used the average predicted probability as the trial-level predicted probability. Using these trial-level predicted probabilities, we computed the area under the receiver operating characteristic curve (ROC AUC). An illustration of calculating ROC AUC is shown in **Figure 2C**. The observed ROC AUC on test data (0.678) exceeded all values from a null distribution generated by training and evaluating models on data with permuted labels. The rightmost panel shows the observed trial-level balanced accuracy (63.1%), compared to the null distribution. To calculate the trial-level balanced accuracy, we determined the optimal threshold for average predicted probability, sweeping from 0 to 1, that maximized balanced accuracy. We applied this process to both observed data and data with permutated labels. **(C)** Average predicted probability in remembered and forgotten trials. Each data point in the plot represents the average predicted probability for a trial, and color denotes the trial condition. For test data, the average predicted probability was higher in remembered trials, compared to forgotten trials (U statistic = 84652, $p < 0.001$, rank-biserial correlation = 0.333, $N_{remembered}$ = 635, $N_{forgotten}$ = 200, Mann–Whitney U test). **(D)** Proportion of predicted remembered saccades and forgotten saccades in remembered and forgotten trials. The predicted label for a saccade was based on the average predicted probability across all recorded SREPs. A chi-squared test of independence revealed a significant association between trial condition and predicted saccade labels ($\chi^2(1)$= 217.89, $p < 0.001$). **(E)** Generalizability of the model across individual subjects. We evaluated the model's performance separately for each subject's test data using saccade- and electrode-contact-level balanced accuracies, averaged across cross-validation (CV) folds. For each subject, we generated a null distribution of balanced accuracy using models trained on permuted labels. The color represents each subject's percentile rank relative to the null distribution. The observed saccade-level balanced accuracy exceeded the null distribution for 6 out of 8 subjects. We note that the other two subjects had the lowest correct recall rate (48% and 59%, subject IDs 6 and 8, **Supplementary Table 3**). Correct recall rate is the proportion of previously presented images that were correctly recognized during the memory test. The inset shows the null distribution and the observed balanced accuracy for a subject (Subject ID 6). **(F)** Electrode contact- and saccade-level balanced accuracy of all random forest models trained in 5-

fold CV. These performance metrics exceeded all values in the null distribution. The green dashed lines show the observed balanced accuracy for each CV fold. **(G)** Concordance of Shapley value–based feature importance across CVs. We used Shapley values to interpret each feature's contribution to the model's predictions. Shapley values, derived from cooperative game theory, quantify each feature's marginal contribution to the model's output by averaging over all possible combinations of feature subsets. The color in the main plot represents the mean absolute Shapley value for each feature, each fold. The lower histogram shows that the Kendall's coefficient of concordance based on the mean absolute Shapley value for trained models across CVs exceeds all concordance coefficients in the null distributions. We generated the null distribution by calculating the concordance coefficient from 5 randomly sampled models trained on permuted data. **(H)** Output agreement across CVs on test data. In addition to **Figure 5H**, we assessed output agreement between models trained on different CV splits based on the ratio of samples with different predictions. For a pair of models trained on actual data in different CV folds, we passed the same test data to both models and computed the proportion of samples with different predictions. The green half violin shows the distribution of this disagreement ratio across models trained on actual data in different CV folds, and the grey half violin represents the null distribution generated from two randomly sampled models trained on permuted data. Summary bars on the right indicate the mean and standard deviation of disagreement ratios for models trained on observed or permuted labels. **(I)** Evaluation of logistic regression for mapping SRND parameters to visual encoding. As functional-anatomical regions are not linearly distributed in the brain, we used a non-linear model (random forest) to learn the mapping from SRND parameters recorded in specific brain regions to visual encoding. To assess whether there is a linear relationship, we trained logistic regression models using the same data and labels as those in **Figure 5B**. We evaluate model performance on held-out test data using electrode contact-level balanced accuracy, saccade-level ROC AUC, and saccade-level balanced accuracy, as in **Figure 5C, D**. We found that logistic regression performed well above chance, although not as well as the random forest model. The rightmost plot shows the weight coefficient for each feature. Caution is warranted when interpreting the negative coefficient for the phase of preceding oscillatory activity, as phase is a circular variable and linear models may not fully capture its periodic nature. **(J)** Relationship between saccadic behavior and visual encoding performance. From left to right, we show the distribution of saccade eccentricity, saccade duration, and saccade direction in remembered and forgotten trials. The rightmost panel shows the difference in circular distribution of saccade direction between remembered and forgotten trials. To quantify the difference in saccade direction, we estimated circular kernel density functions for remembered and forgotten trials and computed the absolute area under the difference between the two density curves. The red dotted line represents the actual difference in saccade direction between remembered and forgotten trials. The blue histogram represents the null distribution of saccade direction differences from permuted trial conditions (remembered vs. forgotten).

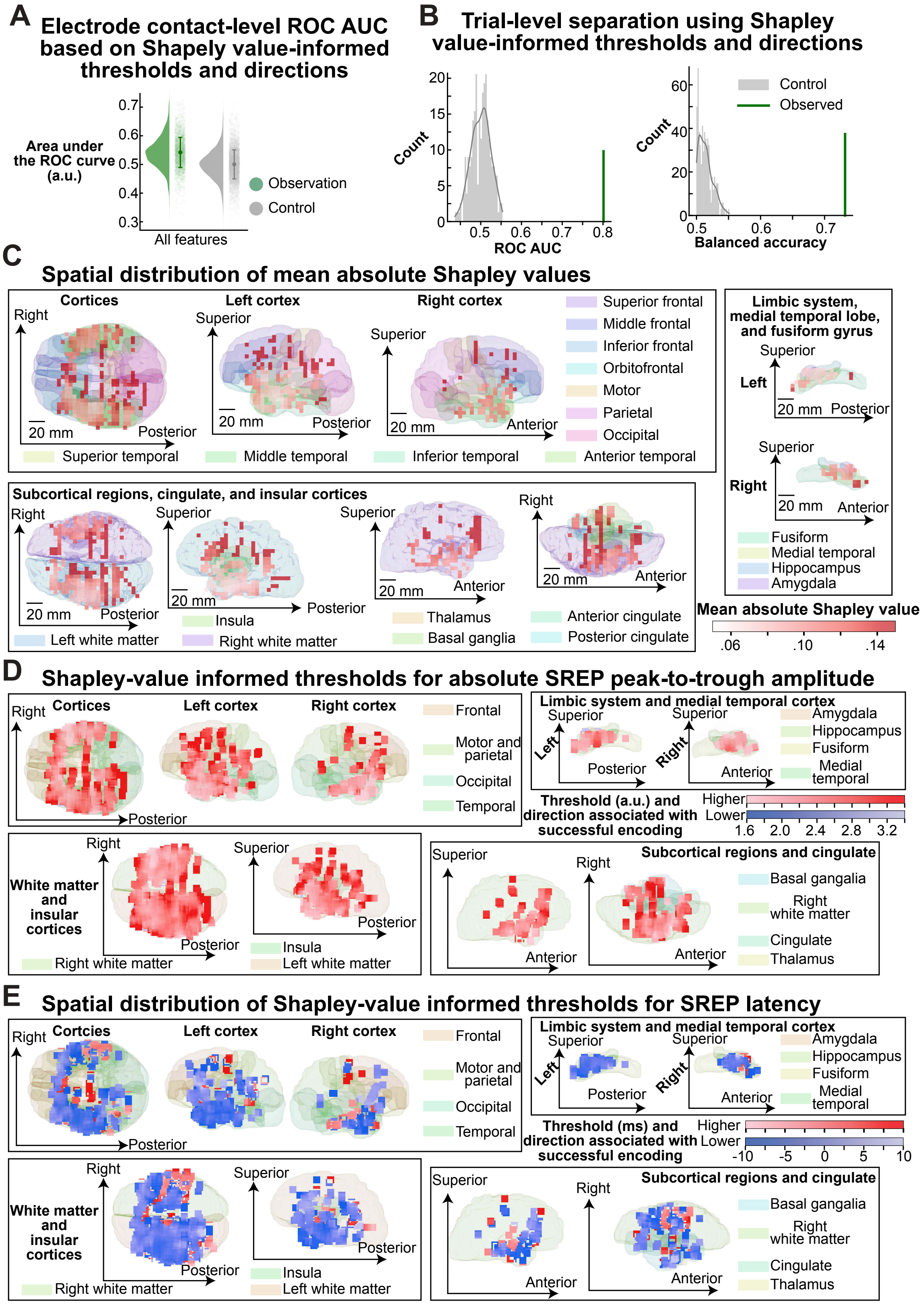

A
Electrode contact-level ROC AUC based on Shapely value-informed thresholds and directions
Area under the ROC curve (a.u.)
0.7
0.6
0.5
0.4
0.3
All features
Observation
Control
B
Trial-level separation using Shapley value-informed thresholds and directions
Count
ROC AUC
Balanced accuracy
Control
Observed
C
Spatial distribution of mean absolute Shapley values
Cortices
Left cortex
Right cortex
Superior frontal
Middle frontal
Inferior frontal
Orbitofrontal
Motor
Parietal
Occipital
Superior temporal
Middle temporal
Inferior temporal
Anterior temporal
Limbic system, medial temporal lobe, and fusiform gyrus
Fusiform
Medial temporal
Hippocampus
Amygdala
Subcortical regions, cingulate, and insular cortices
Insula
Left white matter
Right white matter
Thalamus
Basal ganglia
Anterior cingulate
Posterior cingulate
Mean absolute Shapley value
.06
.10
.14
D
Shapley-value informed thresholds for absolute SREP peak-to-trough amplitude
Frontal
Motor and parietal
Occipital
Temporal
Limbic system and medial temporal cortex
Threshold (a.u.) and direction associated with successful encoding
Higher
Lower
1.6
2.0
2.4
2.8
3.2
White matter and insular cortices
Subcortical regions and cingulate
Cingulate
E
Spatial distribution of Shapley-value informed thresholds for SREP latency
Cortcies
Threshold (ms) and direction associated with successful encoding
-10
-5
0
5
10

**Fig. S12 (Figure 6–figure supplement 1)**. **Validation and illustration of Shapley value-informed thresholds and directions for successful visual encoding**. **(A)** Area under the receiver operating characteristic (ROC) curve for classifying whether a SREP recorded at an electrode contact during a saccade occurred in a remembered or forgotten trial, using recall likelihoods (detailed in **Figure 6D**) computed from Shapley value-informed thresholds and directions. We computed the ROC by systematically varying the decision threshold applied to the recall likelihood; that is, at each threshold, we computed the true positive rate and the false positive rate. We generated the control thresholds and directions by randomly sampling thresholds from the observed feature values. The area under the curve (AUC) was higher using recall likelihood based on actual Shapley value-informed threshold and direction ($N_{observation}$ = 2707, $N_{control}$ = 5414, p < 0.001, Mann-Whitney U test, U = $1.067 \times 10^7$, Cohen's d = 0.817). **(B)** Area under the ROC curve and balanced accuracy of separating remembered from forgotten trials using recall likelihoods aggregated across all saccades and electrode contacts for each trial. The grey distribution represents the ROC curve or balanced accuracy based on surrogate recall likelihood computed from control Shapley value-informed thresholds and directions. We generated the control thresholds and directions by randomly sampling thresholds from the observed feature values. The green vertical line represents the actual ROC curve or balanced accuracy based on the actual recall-likelihood. **(C)** Degree of contribution of SRND parameters to successful visual encoding across anatomical regions. We quantified, within the brain, where SRND parameters contributed more to successful visual encoding using mean absolute Shapley values for each electrode contact. We projected the 3D brain surface and electrode coordinates onto a 2D plane. We used a heatmap to represent the spatial pattern of mean absolute Sharpey values. To generate the heatmap, we discretized this 2D space into a 5 mm × 5 mm grid. For each electrode, we assigned its mean absolute Shapley value to the nearest grid element to preserve anatomical specificity. To account for the recording sensitivity of stereo-encephalogram signals, estimated to be approximately 10 mm, we computed the local average within a 10 mm radius for each grid element. Heatmap elements that have no nearby electrodes are fully transparent. Three panels show spatial distributions of mean absolute Shapley values across (i) cortices, (ii) subcortical structures, cingulate and insular cortices, and (iii) limbic regions. The heatmap color encodes the mean absolute Shapley values. The background anatomical surfaces provide spatial reference and are color-coded by functional-anatomical region; their colors do not represent Shapley values. **(D)**

Thresholds and directions of SREP magnitude (i.e., peak-to-trough amplitude) associated with successful visual encoding across anatomical regions. Since directions may differ for adjacent grid elements, spatial averaging is not appropriate. To account for the recording sensitivity of stereo-encephalogram signals, we considered electrodes within a 10 mm radius for each grid element. We determined the dominant direction, defined as whether higher or lower absolute peak-to-trough amplitude values were more frequently associated with successful encoding. For each grid element, we computed the average of Shapley value-informed thresholds for electrode contacts within a 10 mm radius whose Shapley value-informed direction matched the majority direction. We used two color schemes to represent the thresholds of SREP magnitude associated with successful visual encoding. The red color map (light red to red) indicates regions where higher feature values were associated with successful encoding. The blue colormap (light blue to blue) indicates regions where lower feature values were associated with successful encoding. Within each color map, the color intensity reflects the threshold, with darker red and blue corresponding to more extreme thresholds. **(E)** Thresholds and directions of SREP onset latency associated with successful visual encoding across anatomical regions.

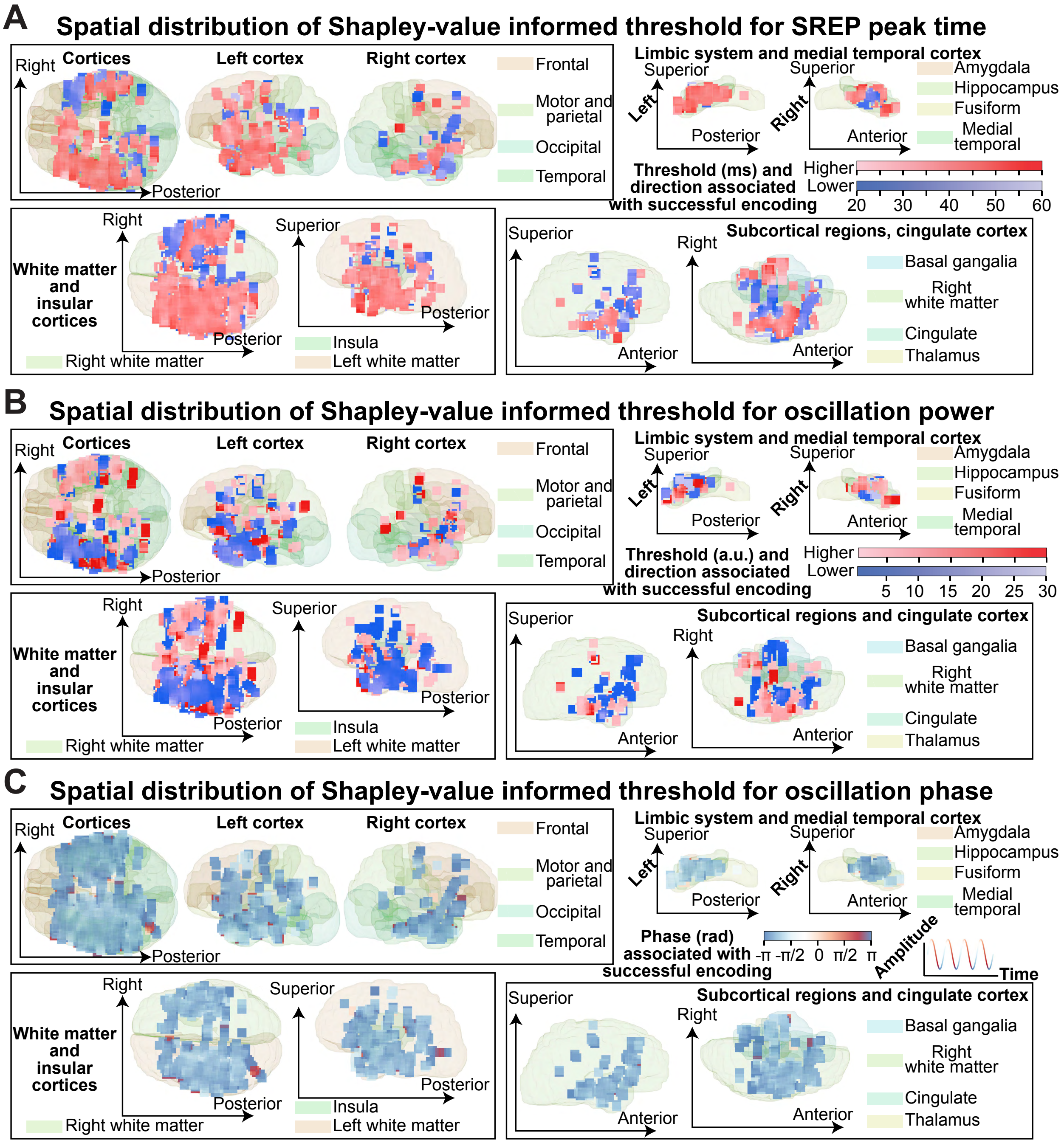


**Fig. S13 (Figure 6–figure supplement 2). SRND parameters associated with successful visual encoding across anatomical regions.** （**A – B**）. Thresholds and directions of SREP peak time and instantaneous power of the preceding oscillation associated with successful visual encoding. We computed the heatmaps using the same method as in **Fig. S12D**. **(C)** Instantaneous phase of the preceding oscillation associated with successful visual encoding. The heatmap color represents the angular mean of the instantaneous phase across electrodes within a 10 mm radius that were associated with successful encoding.

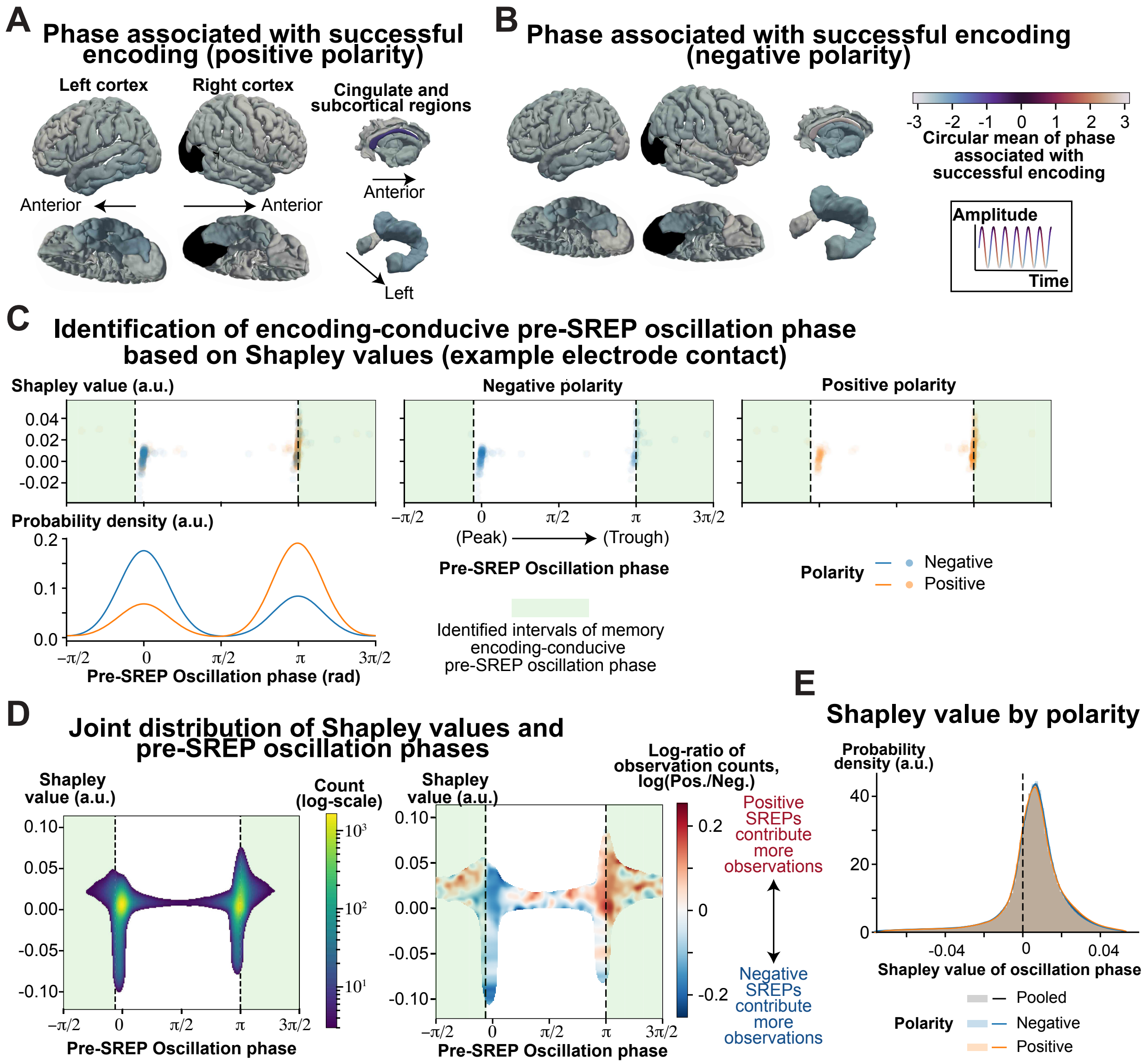


**Fig. S14 (Figure 6–figure supplement 3). Pre-SREP oscillatory phase associated with successful visual encoding across anatomical regions. (A–B)**. Spatial distribution of oscillation phase half oscillation cycle before SREP onset latency associated with successful visual encoding for positive (**A**) and negative polarity (**B**) SREPs. We computed the oscillation cycle based on the dominant oscillation frequency at each location. (**C**) Identification of encoding-conducive ranges of pre-SREP oscillation phases using Shapley values. This panel shows an example from one electrode contact in the left middle temporal gyrus. The right plot in the top row shows the Shapley values associated with the pre-SREP oscillatory phase, with color indicating SREP polarity. The middle and right plots in the top row show the same relationship, restricted to negative- and positive-polarity SREPs, respectively. Although phase distributions differed between polarities, the phase interval associated with higher Shapley values (i.e., greater predicted probability of successful encoding) was consistent across polarities. We identified encoding-conducive phase intervals by partitioning the oscillation phases into two segments that minimized the within-segment variance of Shapley values, and comparing the mean Shapley values between segments (see **MATERIALS AND METHODS:** Shapley value-informed thresholds and directions). For visualization, we displayed the phase on the interval from $-\frac{\pi}{2}$ to $\frac{3\pi}{2}$ radians to center the phase clusters observed near 0 and π radians. (**D**) Joint distribution of pre-SREP oscillatory phase and corresponding Shapley values across all electrode contacts. The left heatmap shows the joint distribution across SREP polarities, represented as a two-dimensional histogram (240 × 240 tiles) spanning oscillation phases from $-\frac{\pi}{2}$ to $\frac{3\pi}{2}$. Color represents the log-transformed count of SREP instances within each tile. Green shady regions denote the interval of memory encoding-conducive oscillation phase (i.e., oscillation phase associated with a higher predicted probability of successful memory encoding). For illustration, we applied a Gaussian kernel (σ = 3 bins) to reduce fragmentation arising from the high-resolution grid. Low-count tiles (≤1 count) were masked. The right heatmap shows the contributions of positive- and negative-polarity SREPs to the observation count at each tile across the same oscillation phase-Shapley value space. Color represents the log-ratio of observation counts between positive- and negative-polarity SREPs within each tile, $\log_{10}$(Positive / Negative). A red tile indicates that positive-polarity SREPs prevail, while a blue tile indicates bins where negative-polarity SREPs predominate. Consistent with the left plot, low-count tiles (≤1 count) were masked. (**E**) Shapley value of pre-SREP oscillation phase, stratified by SREP polarity. Curves show the probability density of Shapley values for pooled SREPs as well as for negative and positive polarity SREPs separately. Probability densities are normalized such that their integrals equal one. To reduce the influence of extreme outliers on kernel density estimation, we trimmed the Shapley value to the 0.5–99.5 percentiles. The dashed vertical line denotes zero Shapley value.

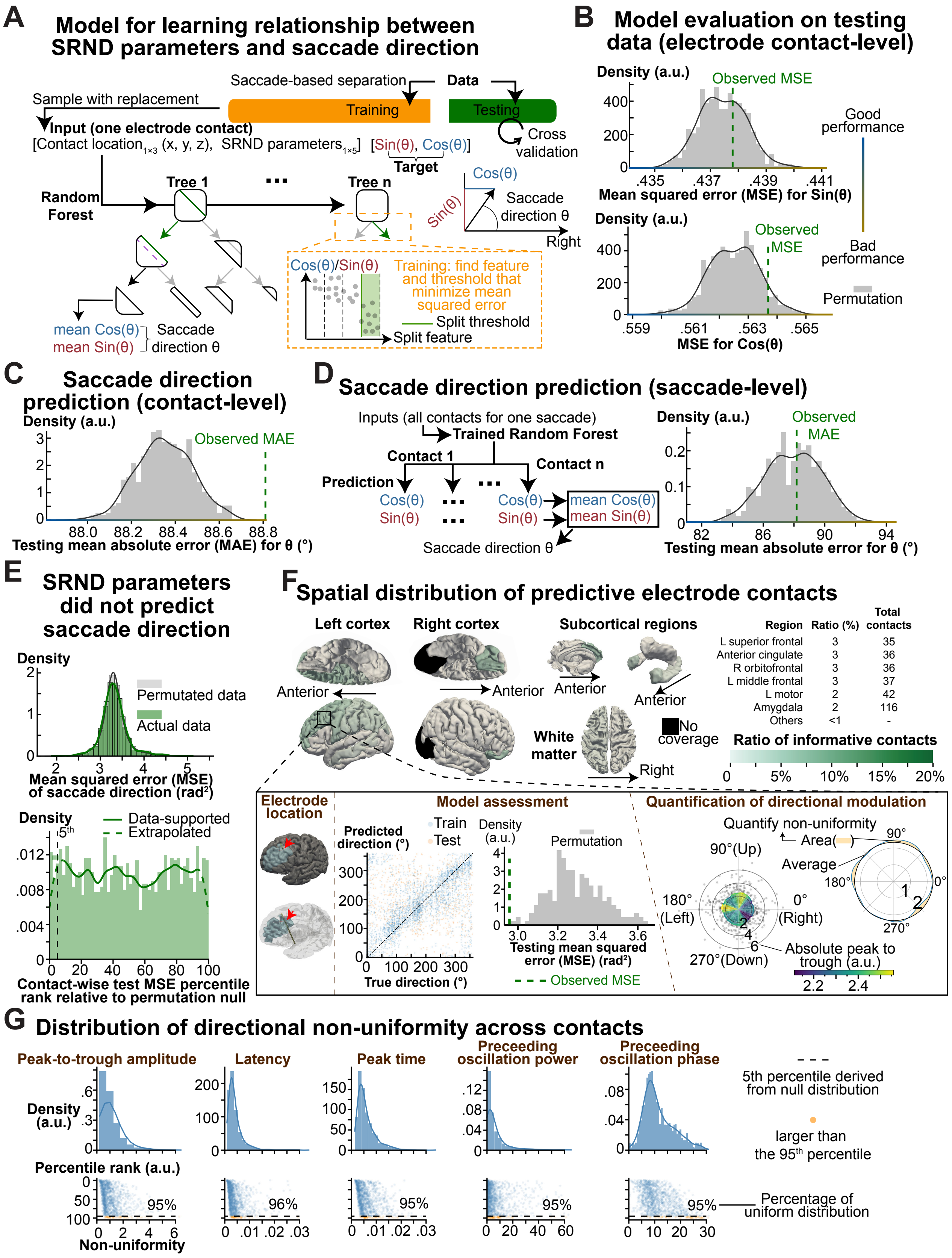
A
Model for learning relationship between SRND parameters and saccade direction
Saccade-based separation
Data
Sample with replacement
Training
Testing
Input (one electrode contact)
[Contact location1×3 (x, y, z), SRND parameters1×5] [Sin(θ), Cos(θ)]
Target
Cross validation
Random Forest
Tree 1
Tree n
Cos(θ)
Sin(θ)
Saccade direction θ
Right
Cos(θ)/Sin(θ)
Training: find feature and threshold that minimize mean squared error
Split threshold
Split feature
mean Cos(θ)
mean Sin(θ)
Saccade direction θ
B
Model evaluation on testing data (electrode contact-level)
Density (a.u.)
Observed MSE
Mean squared error (MSE) for Sin(θ)
Good performance
Bad performance
Permutation
Observed MSE
MSE for Cos(θ)
C
Saccade direction prediction (contact-level)
Density (a.u.)
Observed MAE
Testing mean absolute error (MAE) for θ (°)
D
Saccade direction prediction (saccade-level)
Inputs (all contacts for one saccade)
Trained Random Forest
Contact 1
Contact n
Prediction
Cos(θ)
Sin(θ)
mean Cos(θ)
mean Sin(θ)
Saccade direction θ
Density (a.u.)
Observed MAE
Testing mean absolute error for θ (°)
E
SRND parameters did not predict saccade direction
Density
Permutated data
Actual data
Mean squared error (MSE) of saccade direction (rad²)
Density
Data-supported
Extrapolated
5th
Contact-wise test MSE percentile rank relative to permutation null
F
Spatial distribution of predictive electrode contacts
Left cortex
Right cortex
Subcortical regions
Anterior
Anterior
Anterior
Anterior
White matter
No coverage
Right
Region | Ratio (%) | Total contacts
L superior frontal | 3 | 35
Anterior cingulate | 3 | 36
R orbitofrontal | 3 | 36
L middle frontal | 3 | 37
L motor | 2 | 42
Amygdala | 2 | 116
Others | <1 | -
Ratio of informative contacts
0 5% 10% 15% 20%
Electrode location
Model assessment
Predicted direction (°)
Train
Test
True direction (°)
Density (a.u.)
Permutation
Testing mean squared error (MSE) (rad²)
Observed MSE
Quantification of directional modulation
Quantify non-uniformity
Area
Average
90°(Up)
180° (Left)
0° (Right)
270°(Down)
Absolute peak to trough (a.u.)
G
Distribution of directional non-uniformity across contacts
Peak-to-trough amplitude
Latency
Peak time
Preceeding oscillation power
Preceeding oscillation phase
Density (a.u.)
Percentile rank (a.u.)
Non-uniformity
95%
96%
95%
95%
95%
5th percentile derived from null distribution
larger than the 95th percentile
Percentage of uniform distribution

**Fig. S15 (Figure 7). Most saccade-related evoked potentials (SREPs) did not encode saccade directions.** (**A**) Random forest regressor for learning the mapping between saccade-related neural dynamic (SRND) model parameters and saccade direction. We trained a random forest using 5-fold cross-validation. We split the training and testing data at the saccade level. The input to the random forest is a vector that includes the spatial locations of electrodes and the SRND parameters. To account for the circular nature of saccade direction, we defined the target as the vector $[Sin(\theta), Cos(\theta)]$, where $\theta$ denotes the saccade direction in angular space. This trigonometric transformation allows the use of Euclidean distance as a loss function and enables regression on a continuous 2D representation. During training, the random forest learned a set of features and split thresholds that minimize the mean squared error at each split node. To encourage spatial generalization while preserving spatial specificity, we added Gaussian-distributed spatial jitter to electrode coordinates, preventing the model from memorizing exact electrode contact locations. After training, each leaf node in a tree outputted $[Sin(\theta), Cos(\theta)]$, averaged across the training samples that reached that node. For SREPs recorded in an electrode contact during a saccade, the predicted saccade direction is the average of the output across all trees. We obtained the predicted saccade direction using the $atan2$, two-argument arctangent function. (**B**) Model evaluation on testing data. To assess whether SRND parameters contain information about saccade direction, we evaluated the model's prediction error on held-out testing data using mean squared error (MSE) between predicted and true [sin(θ), cos(θ)] values. Lower MSE indicates a relationship between SRND parameters and saccade direction, while higher MSE indicates little to no such relationship. The vertical green dashed line represents the observed MSE. The gray histogram shows the null distribution of MSE values obtained by training the model on data with permuted target directions. (**C**) Electrode contact-level prediction of saccade direction. We quantified prediction performance at the level of individual electrodes using the mean absolute error (MAE) between predicted and true saccade directions. (**D**) Saccade-level prediction of saccade direction. For each saccade, we averaged the predicted [sin(θ), cos(θ)] vectors across electrodes to obtain the saccade-level prediction of saccade direction. The plot on the right compares the observed MAE against a null distribution generated by permuting saccade directions during training. (**E**) SRND parameters in most electrode contacts did not contain information about saccade direction. The top plot shows the distribution of MSE between predicted and true saccade directions across electrode contacts, compared to a null distribution. For each electrode contact, we computed the observed MSE and generated a null distribution by permuting saccade directions during cross-validation 200 times. The bottom plot shows the distribution of MSE percentile rank relative to each electrode contact's null distribution. The vertical black dashed line marks the 5th percentile, and the green dashed line represents kernel density estimation outside the data support. (**F**) Ratio of saccade direction-informative electrode contacts across anatomical regions. We identified electrode contacts where SRND parameters were informative of saccade direction using permutation testing, by comparing the percentile rank of each contact's saccade-level test MSE within its permutation-derived null distribution against a Bonferroni-corrected significance threshold ($\alpha = 0.05$, corrected for the number of electrode contacts). The color of each anatomical region represents the proportion of saccade direction-informative electrode contacts among all contacts within that region. The inset illustrates an example informative electrode located in the left middle frontal gyrus. In the two 3D brain models on the left, the left middle frontal gyrus is highlighted in blue, and red arrows indicate the electrode location. The first middle plot in the inset illustrates the relationship between the predicted and true saccade directions for the training and testing data. The second middle plot shows the observed MSE for the testing data and the null distribution. The rightmost plots show the explanatory analysis of directional modulation for each SRND parameter. For example, we estimated the circular kernel density of the absolute peak-to-trough value as a function of saccade direction. We quantified directional modulation as non-uniformity, operationally measured as the integral of the absolute difference between the kernel density curve and the mean absolute peak-to-trough value across the angular domain. (**G**) Distribution of directional non-uniformity across electrode contacts. The top row shows the distribution of non-uniformity for SRND parameters. The bottom row shows the corresponding percentile rank of non-uniformity relative to each contact's null distribution. Each data point represents an electrode contact. The horizontal dashed line marks the 5th percentile of the null distribution. The annotated percentage indicates the proportion of electrode contacts whose non-uniformity is less than the 95th percentile of their relative null distribution.

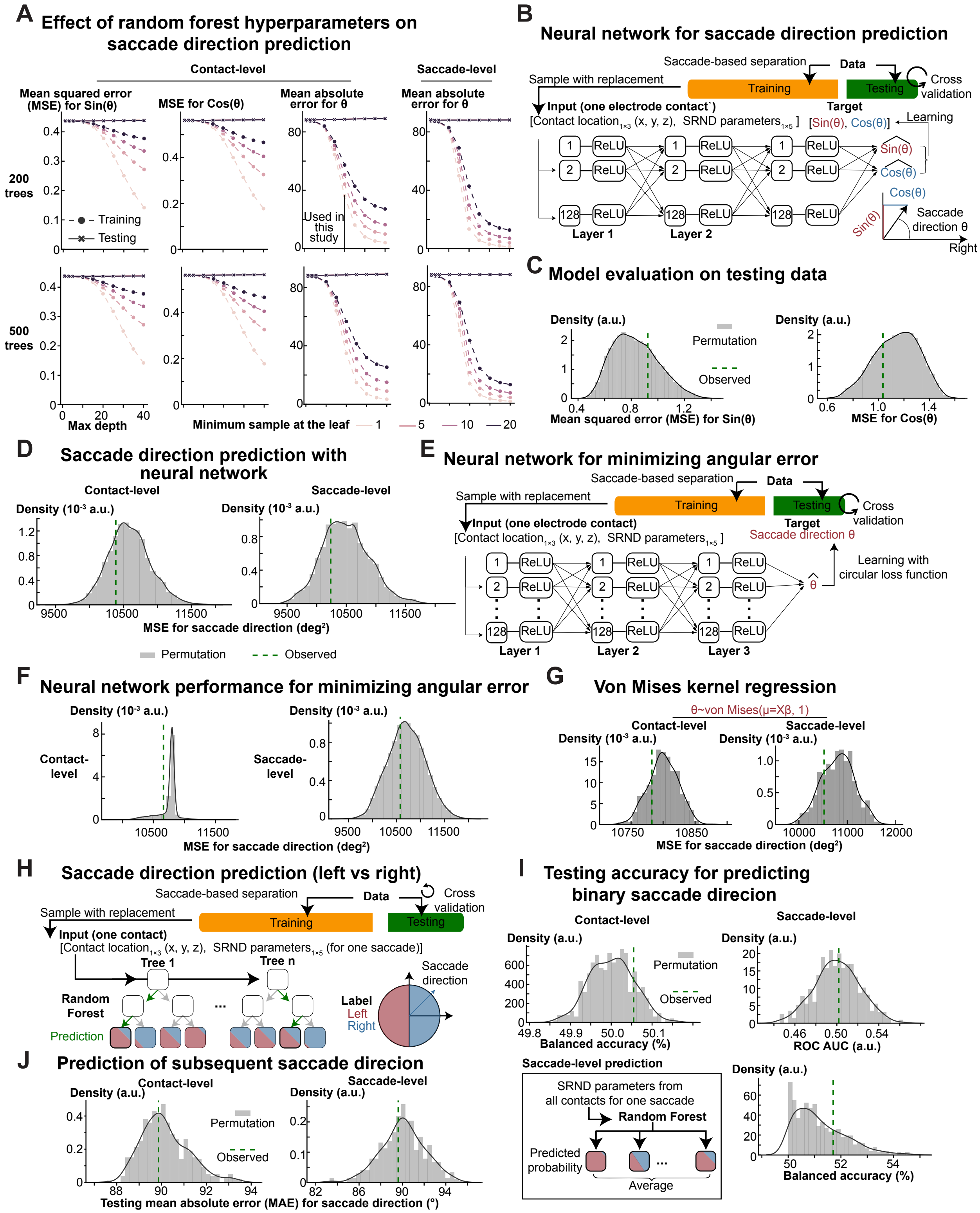


**Fig. S16 (Figure 7–figure supplement 1). Most SREPs did not encode saccade directions, nor subsequent saccade directions. (A)** Effect of hyperparameters on model performance during training and testing. We evaluated how different hyperparameter settings affected the performance of random forests for predicting saccade direction. The hyperparameter includes the number of trees, the maximum tree depth, and the minimum sample size at the leaf. In general, deeper trees with a small sample size can learn more complex patterns but risk overfitting. In each plot, color represents the minimum sample at the leaf, and the x-axis represents the maximum depth. From left to right, the y-axis represents electrode contact-level mean squared error (MSE) for $Sin(\theta)$, $Cos(\theta)$, saccade direction $\theta$, and saccade-level MSE for saccade direction $\theta$. The top row presents the model's performance using 200 decision trees, and the bottom row shows the model's performance with 500 decision trees. In this study, we used a default hyperparameter setting of 200 decision trees, a maximum depth of 20, and a minimum leaf size of 1. **(B)** Neural network architecture for predicting saccade direction. To rule out the possibility that the random forest model might not capture the relationship between saccade direction and SRND parameters, we implemented a neural network with three fully connected hidden layers. The input included electrode contact locations and EP characteristics. The output layer had two units representing sine and cosine of the saccade direction $Sin(\theta)$, $Cos(\theta)$. Each unit in the hidden layers was followed by a Rectified Linear Unit (ReLU) activation function to model non-linearity. **(C)** Evaluation of the neural network for predicting saccade direction. The two plots show the observed contact-level mean squared error (MSE) for $Sin(\theta)$ and $Cos(\theta)$, compared to null distributions of MSE obtained from neural networks trained on permuted data. **(D)** Electrode contact- and saccade-level MSE for saccade direction $\theta$. To calculate saccade-level MSE, we averaged the outputs [$Sin(\theta)$, $Cos(\theta)$] across electrode contacts, and computed the predicted saccade direction using $atan2$, the two-argument arctangent function. The vertical green dashed marks the observed MSE for saccade direction, and the grey histogram represents the null distribution of MSE obtained from neural networks trained with permuted data. **(E)** Neural network architecture for minimizing angular loss. As a complementary model to predicting the trigonometric function of the saccade angle (shown in panel B), we implemented a neural network to minimize angular loss. One unit in the output layer represents the saccade direction. **(F)** Evaluation of a neural network for minimizing angular loss. The vertical green dashed lines show the observed contact- and saccade-level MSEs between the predicted and actual saccade directions. The grey histogram represents the null distribution of MSEs from neural networks trained on permuted data. **(G)**. Saccade direction prediction using circular kernel (i.e., von Mises kernel) regression. To assess whether SRND parameters contribute linearly to saccade direction, we fitted a GLM with a von Mise link function. The plots show the observed contact- and saccade-level MSEs between predicted and actual saccade directions and their null distributions. **(H)** Random forest model for classifying binary saccade direction using SRND parameters. We binarized saccade direction as left vs. right, comparable to the prediction of successful visual encoding (**Figure 5B**). We trained the random forest classifier with inputs of SRND parameters. **(I)** Binary classification of saccade direction using a random forest classifier. The top-left plot shows the observed electrode contact-level balanced accuracy for testing data and its surrogate distribution. We generated the null distribution for testing balanced accuracy by training the random forest classifier on permuted saccade-direction labels. Similar to visual encoding prediction, we used the averaged predicted probability for rightward saccades across electrode contacts during a saccade as the saccade-level predicted probability. The right plots show the distribution of saccade-level area under the receiver operating characteristic (ROC) curve and the optimal balanced accuracy. We obtained the threshold for optimal balanced accuracy. We generated the null distribution of saccade-level area under the ROC curve and balanced accuracy using the same procedure but with permuted data. **(J)** Prediction of subsequent saccade direction using a random forest regressor. We identified subsequent saccades as the next saccade within 500ms. The random forest regressor structure is shown in **Fig. S15A**. The two plots show the observed contact- and saccade-level testing mean absolute error for saccade direction. Grey histograms represent the null distributions of the testing contact- and saccade-level mean absolute errors, generated from models trained on permuted data.

**Table S1 Data Summary**

| ID | Experiment duration (day) | Number of study session | Number of test session | Total number of valid visual encoding trials * | Total number of valid saccades |
|---|---|---|---|---|---|
| 1 | 4 | 4 | 4 | 184 | 1010 |
| 2 | 4 | 8 | 8 | 187 | 1112 |
| 3 | 2 | 4 | 4 | 91 | 431 |
| 4 | 4 | 8 | 8 | 196 | 1447 |
| 5 | 6 | 12 | 12 | 263 | 1561 |
| 6 | 2 | 4 | 4 | 69 | 392 |
| 7 | 2 | 4 | 4 | 80 | 1063 |
| 8 | 2 | 4 | 4 | 30 | 292 |

*The criteria for a valid visual encoding trial include 1. Based on the eye-tracking data, subjects maintained their gaze on the monitor for more than half the duration of each trial; 2. No brain stimulation was delivered during this trial.

**Table S2 Patient Characteristics**

| ID | Epilepsy diagnosis | Sex | Age | Handed ness | WAIS |
|---|---|---|---|---|---|
| 1 | Focal epilepsy with seizure onset in the left hippocampus | F | 36–40 | N/A | 90 |
| 2 | Multifocal mesial temporal lobe epilepsy | F | 36–40 | N/A | N/A |
| 3 | Multifocal epilepsy with seizure onset in the hippocampus | F | 41–45 | R | N/A |
| 4 | Focal left hippocampal epilepsy | M | 36–40 | R | 98 |
| 5 | Focal right hippocampal epilepsy | M | 36–40 | R | 76 |
| 6 | Focal epilepsy in left supplementary motor area | F | 36–40 | R | N/A |
| 7 | No spontaneous seizures recorded | F | 31–35 | R | 107 |
| 8 | Focal epilepsy with seizure onset in the right hippocampus and amygdala | F | 22–25 | R | 88 |

N/A: not available. In the Sex column, ‘F’ stands for ‘female’ and ‘M’ stands for ‘male’. In the Handedness column, ‘L’ stands for ’left’ and ‘R’ stands for ‘right’. WAIS: Wechsler Adult Intelligence Scale.

**Table S3 Visual encoding behavioral results**

| ID | Task ID | The number of: Visual encoding trial | The number of: Tested visual encoding trial | The number of: Remembered tested visual encoding trials | Correct recall rate (%) | The number of: Test trials | The number of: Novel test trials | The number of: Correct recognition of novel test trials | Correct recognition rate of novel test trials | Overall correct recognition rate |
|---|---|---|---|---|---|---|---|---|---|---|
| All trials | | | | | | | | | | |
| 1 | Encoding1 | 180 | 160 | 110 | 69% | 240 | 80 | 47 | 59% | 65% |
| 1 | Encoding2 | 180 | 160 | 155 | 97% | 240 | 80 | 5 | 6% | 67% |
| 2 | Encoding1 | 180 | 160 | 83 | 52% | 240 | 80 | 72 | 90% | 65% |
| 2 | Encoding2 | 180 | 160 | 126 | 79% | 240 | 80 | 54 | 68% | 75% |
| 3 | Encoding1 | 180 | 160 | 132 | 83% | 240 | 80 | 51 | 64% | 76% |
| 4 | Encoding1 | 180 | 160 | 121 | 76% | 240 | 80 | 68 | 85% | 79% |
| 4 | Encoding2 | 180 | 160 | 99 | 62% | 240 | 80 | 64 | 80% | 68% |
| 5 | Encoding1 | 180 | 160 | 131 | 82% | 240 | 80 | 59 | 74% | 79% |
| 5 | Encoding2 | 180 | 159 | 140 | 88% | 240 | 81 | 42 | 52% | 76% |
| 5 | Encoding3 | 180 | 160 | 148 | 93% | 240 | 80 | 38 | 48% | 78% |
| 6 | Encoding1 | 160 | 80 | 38 | 48% | 160 | 80 | 56 | 70% | 59% |
| 7 | Encoding1 | 160 | 80 | 71 | 89% | 160 | 80 | 65 | 81% | 85% |
| 8 | Encoding1 | 160 | 80 | 47 | 59% | 160 | 80 | 57 | 71% | 65% |
| Valid trials only* | | | | | | | | | | |
| 1 | Encoding1 | 91 | 73 | 50 | 68.5% | - | - | - | - | - |
| 1 | Encoding2 | 93 | 77 | 74 | 96.1% | - | - | - | - | - |
| 2 | Encoding1 | 95 | 77 | 36 | 46.8% | - | - | - | - | - |
| 2 | Encoding2 | 92 | 74 | 61 | 82.4% | - | - | - | - | - |
| 3 | Encoding1 | 91 | 77 | 64 | 83.1% | - | - | - | - | - |
| 4 | Encoding1 | 97 | 80 | 62 | 77.5% | - | - | - | - | - |

| | | | | | | | | | | |
|---|---|---|---|---|---|---|---|---|---|---|
| 4 | Encoding2 | 99 | 79 | 50 | 63.3% | - | - | - | - | - |
| 5 | Encoding1 | 98 | 79 | 62 | 78.5% | - | - | - | - | - |
| 5 | Encoding2 | 78 | 67 | 58 | 86.6% | - | - | - | - | - |
| 5 | Encoding3 | 87 | 75 | 69 | 92.0% | - | - | - | - | - |
| 6 | Encoding1 | 69 | 35 | 14 | 40.0% | - | - | - | - | - |
| 7 | Encoding1 | 80 | 40 | 35 | 87.5% | - | - | - | - | - |
| 8 | Encoding1 | 30 | 18 | 12 | 66.7% | - | - | - | - | - |

Correct recall rate represents the percentage of remembered visual encoding trials over the tested visual encoding trials. The overall correct recognition rate is calculated as the sum of true positives (old stimuli correctly recalled) and true negatives (new stimuli correctly rejected) divided by the total number of test trials. *The criteria for a valid visual encoding trial include 1. Based on the eye-tracking data, subjects maintained their gaze on the monitor for more than half the duration of each trial; 2. No brain stimulation was delivered during this trial.

**Table S4 Electrode coverage**

| ID | Task ID | The total number of implanted electrode contacts | The number of bad contacts* | The number of valid contacts | Average number of valid contacts for each subject |
|---|---|---|---|---|---|
| 1 | Encoding1 | 225 | 4 | 221 | 220.5 |
| | Encoding2 | 225 | 5 | 220 | |
| 2# | Encoding1 | 229 | 6 | 223 | 224.5 |
| | Encoding2 | 229 | 3 | 226 | |
| 3# | Encoding1 | 170 | 4 | 166 | 166 |
| 4# | Encoding1 | 150 | 4 | 146 | 145.5 |
| | Encoding2 | 150 | 5 | 145 | |
| 5# | Encoding1 | 214 | 3 | 211 | 210.3 |
| | Encoding2 | 214 | 4 | 210 | |
| | Encoding3 | 214 | 4 | 210 | |
| 6# | Encoding1 | 229 | 10 | 219 | 219 |
| 7# | Encoding1 | 190 | 2 | 188 | 188 |
| 8 | Encoding1 | 222 | 3 | 219 | 219 |
| Total | | | | | 1592.8 |

*Electrodes exhibiting epileptiform activity or artifactual signals (e.g., due to poor contact) were excluded prior to re-referencing. # For a subset of subjects, 2 Ad-yech Behnke Fried Depth electrodes, each with 8-contact micro recording, were implanted. Data from micro wires were excluded if they displayed epileptiform activity or artifactual signals. The number of micro wires is not included in this table.

**Movie S1.**
Figure 4–supplementary video.
